# Relative effectiveness of high-dose vs standard-dose influenza vaccines in preventing hospitalizations: a national retrospective cohort study in France, 2022/23 season

**DOI:** 10.1101/2024.10.11.24315085

**Authors:** Hélène Bricout, Marie-Cécile Levant, Pascal Crépey, Gaëtan Gavazzi, Jacques Gaillat, Marie Dufournet, Nada Assi, Benjamin Grenier, Fanny Raguideau, Fabienne Péretz, Camille Salamand, Anne Mosnier, Laurence Watier, Odile Launay, Matthew M. Loiaconol

## Abstract

**Background:** A French retrospective observational cohort study conducted during the 2021/22 influenza season found that the high-dose influenza vaccine (HD) was more effective than standard-dose vaccines (SDs) in preventing influenza-related hospitalizations in the elderly. The study continued during the 2022/23 season to obtain more accurate results and validate the findings during a different influenza season.

**Material and methods:** Data from community-dwelling 65+ adults vaccinated with HD or SD during the 2022/23 vaccination campaign were extracted from the National Health database. Hospitalizations were recorded from 14 days after vaccination until 30 June 2023. HD and SD recipients were matched using a propensity score with an exact constraint on age, sex, vaccination week, and geographical region. Associations between vaccines and hospitalizations (influenza or non-influenza related) were assessed by estimating incidence rate ratios and converting them to HD vs SD vaccine relative effectiveness (rVE).

**Results:** 675,412 HD recipients were matched to 2,701,648 SD recipients. The HD vs SD rVE for influenza-related hospitalizations was 27.39% [95%CI: 19.79;34.27]. It ranged from 22.65% [9.84;33.64] to 33.55% [21.19;43.98] across age groups, indicating that HD resulted in consistently better protection than SDs against influenza-related hospitalizations in all age groups, with the highest effect observed in 85+.

**Conclusion:** Our study is the first to publish rVE data comparing HD and SD in France, for the 2022/23 influenza season. Its findings reaffirm the benefit of HD vs SD in reducing influenza-related hospitalizations in a real-world setting. HD could help reduce the burden of severe respiratory infections in the elderly.

Registration number: Not applicable.

**Highlights:** *Take-home message:* In France, during the 2022/23 influenza season, the high-dose vaccine has reduced influenza-related hospitalizations by 27.4% compared to standard-dose vaccines among community-dwelling 65+ adults, with a clinical benefit observed across all age groups.

## Introduction

Across most countries, including France, influenza vaccination programs aim to reduce the number of severe cases, complications, and deaths [1]. Amongst these programs, and specifically within the French context, recommendations particularly target high-risk individuals such as adults aged 65 and over (65+). The heightened susceptibility to infections and severe outcomes in this population can be explained by immunosenescence, age-related anatomical and physiological alterations changes, a higher prevalence of comorbidities, and malnutrition [2].

While effective and essential from the public health perspective, current inactivated standard-dose influenza vaccines (i.e., influenza vaccines containing 15 µg of hemagglutinin per recommended strain) offer suboptimal protection in older adults, primarily due to immunosenescence and age-related increased inflammation [3]. To enhance immune response and decrease influenza-related morbidity and mortality, a high-dose influenza vaccine (HD), containing four times each of the antigens of the standard dose influenza vaccines (SDs), was developed. Initially trivalent and licensed in the USA in 2009, a quadrivalent formulation was developed a decade later and approved for use in the USA, Australia, Canada, and Europe [2]. The superior relative efficacy (rVE) of HD against severe clinical outcomes compared to SDs has been demonstrated in a pivotal randomized controlled study, confirmed by several randomized clinical trials, and consistently found in observational studies [4–7]. According to a recent meta-analysis [7], HD reduced hospitalizations due to pneumoniae or influenza by 23.5% compared to SDs in the 65+ adults.

In France, HD was used for the first time during the 2020/21 influenza season. In May 2020, the French Health Authority additionally recommended its use in 65+ under the same conditions of distribution, administration, and reimbursement as SD. In practice, before the start of each influenza season, high-risk individuals are provided with a voucher to get their influenza vaccine free of charge at a pharmacy. The vaccine is then administered by the pharmacist or another healthcare professional authorized to vaccinate, typically the family practitioner or a nurse. In line with the recommendations of the French Health Authority, any of the available influenza vaccines can be used for adults aged 65+; however, preferential use of HD for 65 is recommended by the French Geriatric Society (Société Française de Gériatrie et Gérontologie) [8, 9].

Using the French national administrative healthcare data collected from the start of the 2021/2022 influenza vaccination campaign to 30 June 2022, we previously conducted an observational study called DRIVEN [10]. This study aimed to estimate the rVE of HD vs SD against influenza-related hospitalizations in France in 65+. It confirmed that HD provided better protection than SD across more than 400,000 HD-and 1,600,000 SD-vaccinated individuals (relative vaccine effectiveness, rVE: 23.3% [95%CI: 8.4;35.8%]). The study was continued during the 2022/23 season to obtain more accurate results given the increased use of HD between the 2021/22 and 2022/23 seasons, and to confirm the findings during a different influenza season, considering potential epidemiological changes. We anticipated that the results from the 2022/23 season would confirm those of the 2021/22 season, improving robustness and precision owing to more individuals vaccinated with HD.

## Materials and methods

### Study design

The method was identical to that used for the 2021/22 influenza vaccination campaign (season 1), described in detail in the article by Bricout et al. [10]. Briefly, DRIVEN-season 2 was an observational retrospective cohort study, based on the National Health Data System (SNDS). It was designed to assess the rVE of HD vs SDs for hospitalization outcomes for the 2022/23 influenza season. Vaccine type (HD or SD) was identified using medication codes (Supplementary Table 1).

### Study population

Individuals aged 65 years or over (65+) on the day of vaccine dispensing with a record of influenza vaccine dispensed at a pharmacy during the official influenza vaccination campaign period (i.e., 18 October 2022/31 March 2023) were included in the study cohort. Individuals remained in the study cohort from the day of influenza vaccine dispensing (index date) up to 30 June 2023, or the day of admission to medico-social housing or a nursing home, or death.

### Data collection

The study used data from the SNDS, part of the National Health Insurance system. The SNDS encompasses anonymous, individual-level data for all healthcare claims for more than 99% of the population residing in France, regardless of their insurance scheme, i.e., close to 68 million people.

Baseline demographics, underlying diseases/medical history (medical conditions), and previous treatments and vaccinations were captured from 01 September 2017 to 31 March 2023. Hospitalizations related to influenza or other causes were collected from 14 days after the index date (i.e., presumed day of the start of the vaccine protection) to the end of follow-up. The index date was defined as the date of the claim for influenza vaccine reimbursement (= dispensing date); it was used as a proxy for influenza vaccination. By convention, the index date was referred to as the date of vaccination, and the individuals were referred to as vaccinated.

### Primary outcomes

Primary outcomes were hospitalizations for influenza and then non-influenza-specific causes (pneumonia, combined pneumonia/influenza, respiratory diseases, or cardiovascular or cardiorespiratory events). Hospitalizations were ascertained by the International Classification of Diseases 10th Revision (ICD-10) discharge diagnosis code. The ICD-10 discharge diagnosis code in the database could be noted as primary (PD), related (RD), or associated (AD). Hospitalizations were ascertained by their ICD-10 discharge diagnosis code (Supplementary Table 1).

### Matching

Since the study relied on retrospective database analysis with routine treatment allocation (no randomization), a treatment selection or indication bias could exist. To adjust for potential confounding effects, each HD-vaccinated individual was matched to four SD-vaccinated individuals using a propensity score with an exact constraint for sex, age group (65-75, >75-85, and >85 years), geographical region, and week of vaccine dispensing. The propensity score was computed using a logistic regression model including other covariables: i.e., medical variables such as previous visits to family practitioners or hospitalizations, vaccine histories such as previous influenza, pneumococcal, or COVID-19 vaccinations; social characteristics such as quintiles of the French social deprivation index (FDep); underlying diseases such as diabetes, obesity, malnutrition, or chronic obstructive pulmonary disease (COPD) [10].

### Statistical methods

To estimate the association between vaccination with HD or SD and hospitalization outcomes incidence rate ratios (IRRs) were calculated and then converted to rVE. Poisson models, negative binomial model, and their zero-inflated counterparts were used to estimate IRRs along with their corresponding 95% confidence interval (95%CI) [11]. The model with the lowest Akaike information criterion (AIC) was chosen [12]. Models included an offset for the log of the follow-up time, which allowed a rate model to be computed. The rVE was computed as ([1-IRR)] * 100), with its corresponding 95%CI by Taylor series variance approximation [13, 14].

IRRs and rVEs were calculated for hospitalizations for influenza, pneumonia, combined pneumonia/influenza, respiratory events, cardiorespiratory events, and cardiovascular events in the whole population and hospitalizations for influenza, pneumonia, and combined pneumonia/influenza per age group.

The main analysis was performed on the matched population using the PD code to ascertain hospitalization diagnosis. Hospitalizations with a discharge diagnosis code associated with COVID-19 were excluded from this main analysis (COVID-19 exclusion). Sensitivity analyses were performed with alternative outcome definitions: i.e., less specific ICD-10 discharge diagnosis code (i.e., PD, RD, or AD) or hospitalizations (i.e., COVID-19 included). To increase the specificity of the outcomes, an analysis was performed considering only hospitalizations that occurred within nine weeks encompassing the week of the peak incidence for the 2022/23 influenza season (from four weeks before to four weeks after). The peak incidence for the 2022/23 influenza season occurred during week 51; the nine-week period therefore ranged from 21 November 2022 to 22 January 2023.

Moreover, a falsification analysis was performed with negative control outcomes (i.e., hospitalizations for urinary tract infections, cataract surgery, or erysipelas), to assess any potential residual bias due to unmeasured confounding, and classical multivariable analysis on the whole population (no matching) to assess the stability of the findings.

Statistical analysis was performed using SAS Software, Version 9.4 (SAS Institute Inc., Cary, NC, USA). To account for the multiplicity of testing, the results of the main and sensitivity analyses were controlled using the Family-wise Error Rate (FWER) through the Bonferroni-Holm procedure. This method provided tight control over the probability of making one or more type I errors (false positives), and the corrected p-values were presented accordingly.

For the falsification analysis, the results were interpreted with an unadjusted level of type I error. The statistical significance threshold was set at 0.05.

## Results

### Study population and vaccine exposure

8,418,163 individuals vaccinated during the 2022/23 influenza vaccination campaign were identified in the database. Of these, 7,914,298 were 65+ adults living in the community. Among community-dwelling 65+, 976,211 (12.33%) were vaccinated with HD and 6,938,087 (87.67%) with SD. After matching (1:4), 675,412 individuals vaccinated with HD and 2,701,648 vaccinated with SD constituted the analysis population (Figure 1). The rate of matching was 71.70% in the HD cohort: 81.09% in the 65-75; 67.82% in the 75 −85; and 60.58% for the 85+. In the SD cohort, 36.15% of the 65 −75, 42.49% of the 75-85, and 48.29% of the 85+ were matched with HD individuals. The main characteristics of each cohort for matched individuals are presented in (Table 1).

**Figure 1.**
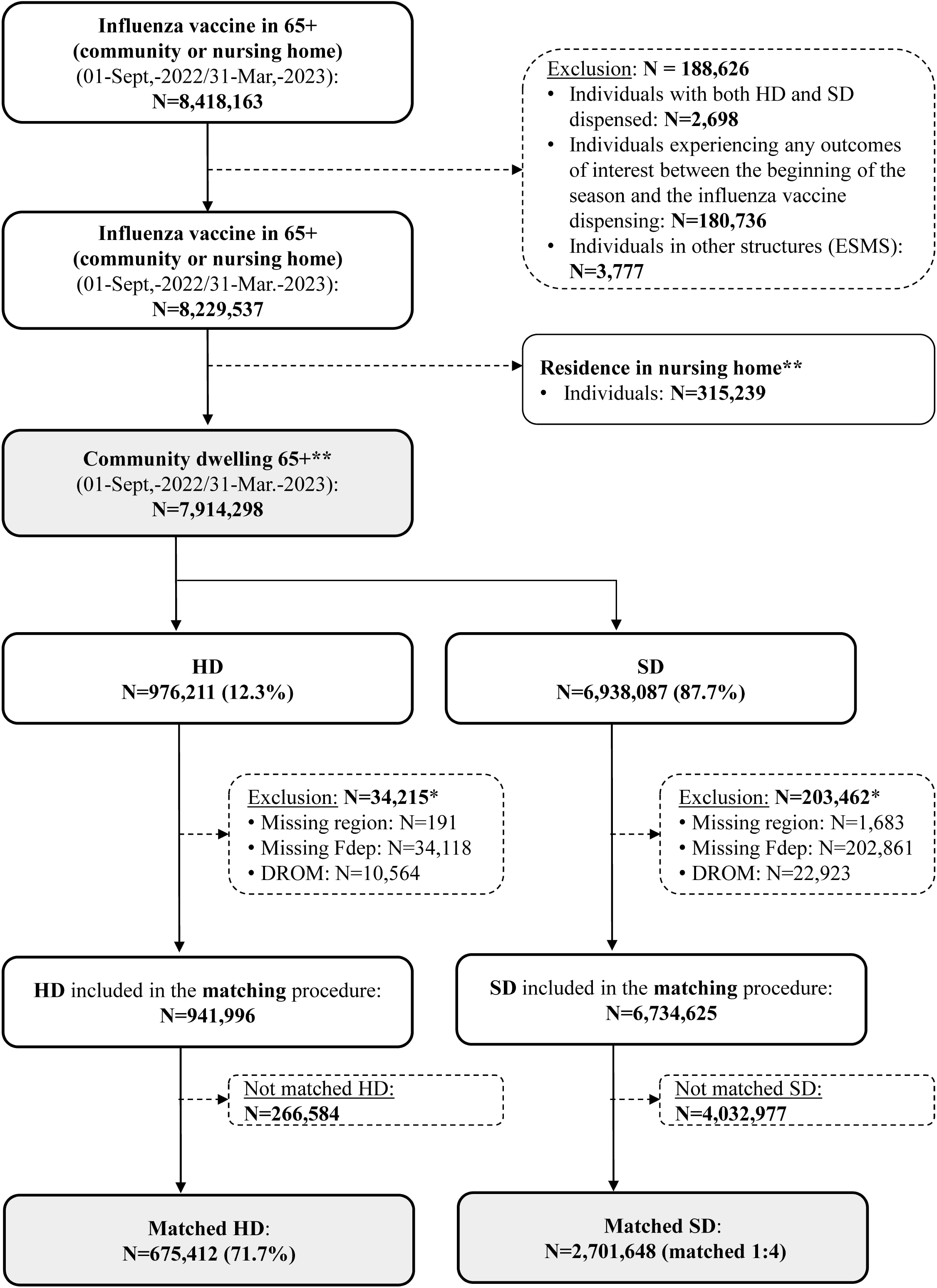
Study flow chart. * Details are non-exclusive: e.g., patients could have been excluded for a missing Fdep and because they resided in the DROM. ** on the date the vaccine is dispensed 65+: aged 65 and over; DROM: abbreviation for overseas departments and regions of France; ESMS: other social health-care institutions; Fdep: deprivation index; HD: high-dose influenza vaccine (or cohort); N: number of individuals; SD: standard-dose influenza vaccine (or cohort)

**Table 1.**
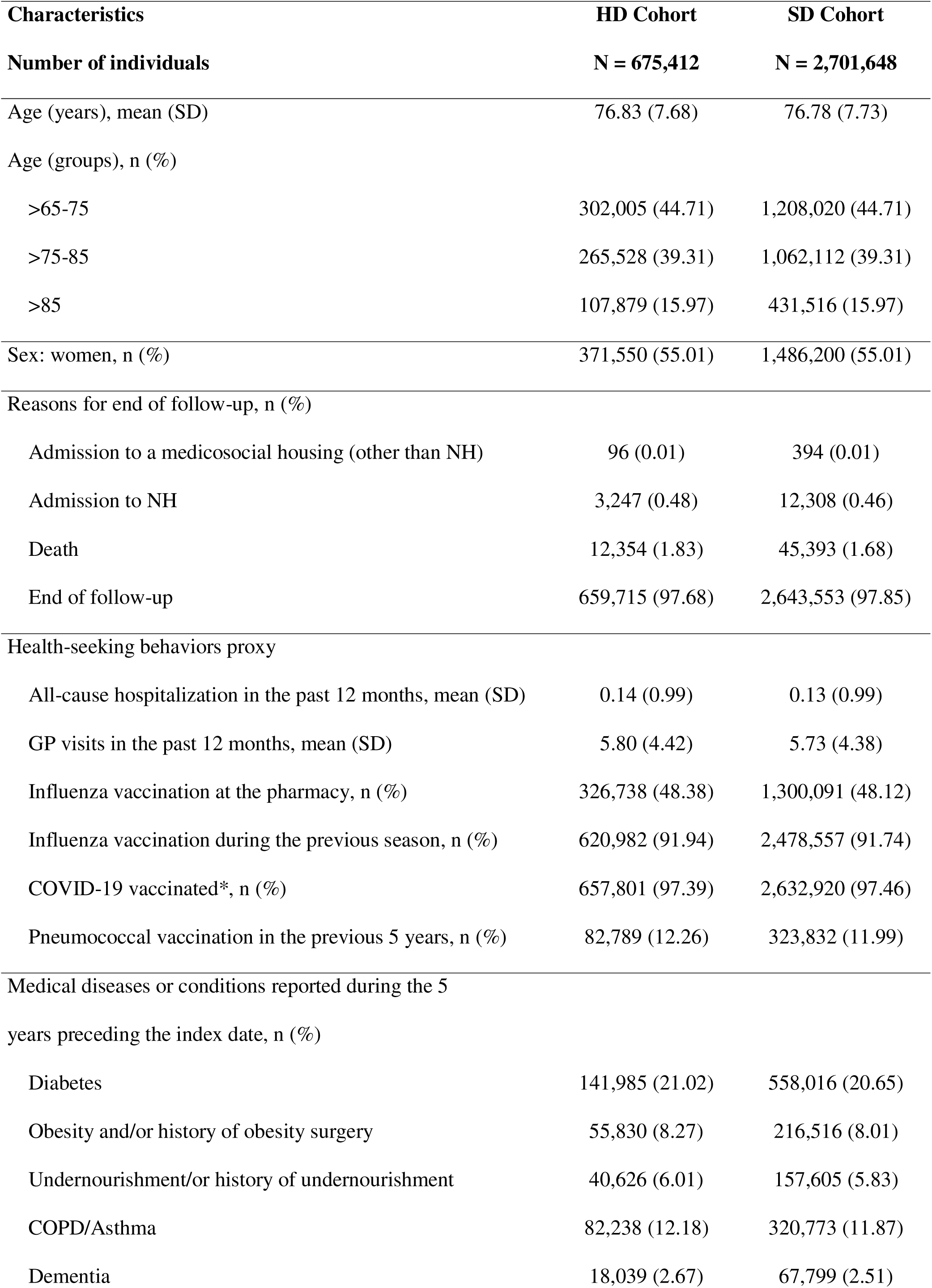
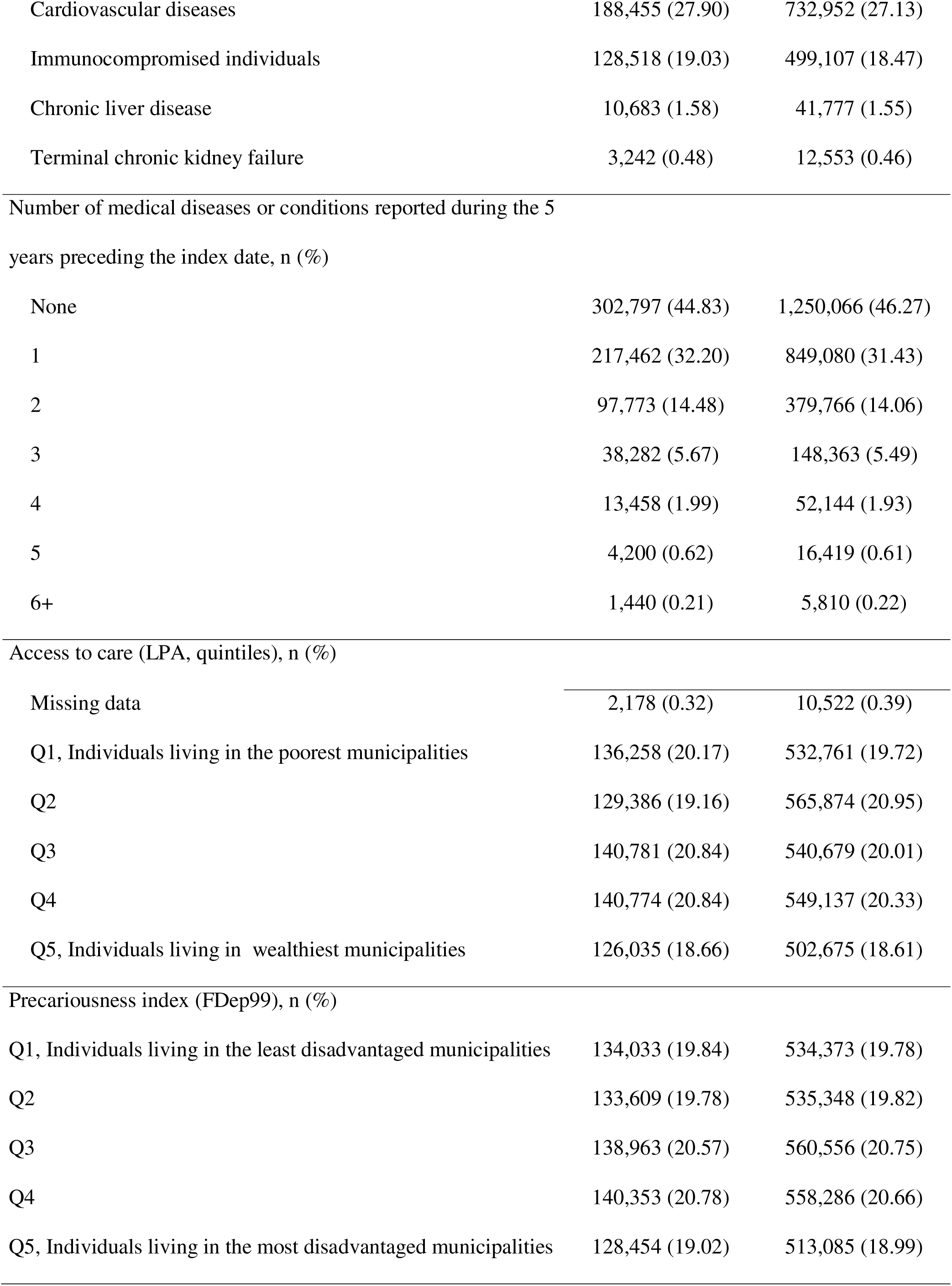
Main baseline characteristics of each cohort (matched individuals only, N=3,377,060) COPD: chronic obstructive pulmonary disease; FDe99: French ecological deprivation index; HD: high-dose influenza vaccine; LPA: local potential accessibility; NH: nursing home; SD: standard deviation; SD: standard dose influenza vaccine * ‘COVID-19 vaccinated’ is a variable identified as such within the database. It reflects the COVID-19 vaccination status of each patient at index date following current guidelines (it can refer to a single dose, 2, or 3, depending on the individual’s eligibility

| Characteristics | HD Cohort | SD Cohort |
| --- | --- | --- |
| **Number of individuals** | **N = 675,412** | **N = 2,701,648** |
| Age (years), mean (SD) | 76.83 (7.68) | 76.78 (7.73) |
| Age (groups), n (%) |  |  |
| >65-75 | 302,005 (44.71) | 1,208,020 (44.71) |
| >75-85 | 265,528 (39.31) | 1,062,112 (39.31) |
| >85 | 107,879 (15.97) | 431,516 (15.97) |
| Sex: women, n (%) | 371,550 (55.01) | 1,486,200 (55.01) |
| Reasons for end of follow-up, n (%) |  |  |
| Admission to a medicosocial housing (other than NH) | 96 (0.01) | 394 (0.01) |
| Admission to NH | 3,247 (0.48) | 12,308 (0.46) |
| Death | 12,354 (1.83) | 45,393 (1.68) |
| End of follow-up | 659,715 (97.68) | 2,643,553 (97.85) |
| Health-seeking behaviors proxy |  |  |
| All-cause hospitalization in the past 12 months, mean (SD) | 0.14 (0.99) | 0.13 (0.99) |
| GP visits in the past 12 months, mean (SD) | 5.80 (4.42) | 5.73 (4.38) |
| Influenza vaccination at the pharmacy, n (%) | 326,738 (48.38) | 1,300,091 (48.12) |
| Influenza vaccination during the previous season, n (%) | 620,982 (91.94) | 2,478,557 (91.74) |
| COVID-19 vaccinated*, n (%) | 657,801 (97.39) | 2,632,920 (97.46) |
| Pneumococcal vaccination in the previous 5 years, n (%) | 82,789 (12.26) | 323,832 (11.99) |
| Medical diseases or conditions reported during the 5<br>years preceding the index date, n (%) |  |  |
| Diabetes | 141,985 (21.02) | 558,016 (20.65) |
| Obesity and/or history of obesity surgery | 55,830 (8.27) | 216,516 (8.01) |
| Undernourishment/or history of undernourishment | 40,626 (6.01) | 157,605 (5.83) |
| COPD/Asthma | 82,238 (12.18) | 320,773 (11.87) |
| Dementia | 18,039 (2.67) | 67,799 (2.51) |
| Cardiovascular diseases | 188,455 (27.90) | 732,952 (27.13) |
| Immunocompromised individuals | 128,518 (19.03) | 499,107 (18.47) |
| Chronic liver disease | 10,683 (1.58) | 41,777 (1.55) |
| Terminal chronic kidney failure | 3,242 (0.48) | 12,553 (0.46) |
| None | 302,797 (44.83) | 1,250,066 (46.27) |
| 1 | 217,462 (32.20) | 849,080 (31.43) |
| 2 | 97,773 (14.48) | 379,766 (14.06) |
| 3 | 38,282 (5.67) | 148,363 (5.49) |
| 4 | 13,458 (1.99) | 52,144 (1.93) |
| 5 | 4,200 (0.62) | 16,419 (0.61) |
| 6+ | 1,440 (0.21) | 5,810 (0.22) |
| Access to care (LPA, quintiles), n (%) |  |  |
| Missing data | 2,178 (0.32) | 10,522 (0.39) |
| Q1, Individuals living in the poorest municipalities | 136,258 (20.17) | 532,761 (19.72) |
| Q2 | 129,386 (19.16) | 565,874 (20.95) |
| Q3 | 140,781 (20.84) | 540,679 (20.01) |
| Q4 | 140,774 (20.84) | 549,137 (20.33) |
| Q5, Individuals living in wealthiest municipalities | 126,035 (18.66) | 502,675 (18.61) |
| Precariousness index (FDep99), n (%) |  |  |
| Q1, Individuals living in the least disadvantaged municipalities | 134,033 (19.84) | 534,373 (19.78) |
| Q2 | 133,609 (19.78) | 535,348 (19.82) |
| Q3 | 138,963 (20.57) | 560,556 (20.75) |
| Q4 | 140,353 (20.78) | 558,286 (20.66) |
| Q5, Individuals living in the most disadvantaged municipalities | 128,454 (19.02) | 513,085 (18.99) |

Before matching (N=7,914,928), several baseline characteristics of HD- and SD-vaccinated individuals differed (Supplementary Table 2). For instance, individuals from the HD cohort were older (median: 77 vs 75 years); they more frequently presented with at least one specific long-term disease (*Affection Longue Durée,* ALD) (52.05% vs 47.69%); they have been more frequently vaccinated at least once against influenza during the previous five years (96.69% vs 94.03%); they had a poorer outcome at the end of the study (2.11% vs 1.46% died). In addition, most of the HD doses (80.79%) were dispensed within the first 4 weeks of the influenza vaccination campaign while most of the SD doses (87.76%) were dispensed within the first 7 weeks (Supplementary Figure 1). Individual characteristics also differed with age. The 85+ more frequently presented with at least one ALD (60.68% vs 51.59% in the 75-85 group and 42.01% in the 65-75 group); they have been more frequently vaccinated at least once against influenza during the previous five years (98.06% vs 97.64% and 90.73%, respectively); they had a poorer outcome at the end of the study (4.99% vs 1.42% and 0.64% died); they were more frequently provided with the doses of influenza vaccine within the first 4 weeks of the vaccination campaign (64.10% vs 58.35% and 54.49%) (Supplementary Table 3).

After matching (N=3,377,060), the two cohorts were overall well balanced (Supplementary Table 4). Standardized differences ranged between −0.1 and 0.1. However, there was a slight trend towards a marginally better health status in individuals vaccinated with SD than HD: the percentages of recipients having or having experienced medical conditions within the last five years were slightly higher in the HD than in the SD cohort as were the percentages of patients who died or were admitted to a nursing home during the study were greater.

Compared to unmatched individuals, matched individuals from the HD cohort were younger (median: 76 vs 80 years) and less frequently reported having specific long-term diseases (50.50% vs 55.97%). Additionally, they exhibited a lower prevalence of medical conditions in the previous five years (55.17% vs 61.74%) and a reduced risk of death during the study period (mortality rate, 0.48% vs 0.75%). Finally, they were not evenly distributed across France: they were more frequently located in the South of France (AURA, PACA, Occitanie, Nouvelle Aquitaine: 42.56% vs 22.20%) than unmatched individuals. In the SD cohort, matched individuals were older (median: 76 vs 74 years) and more frequently reported having specific long-term diseases (49.16% vs 46.74%) than unmatched individuals. Additionally, they exhibited a higher prevalence of medical conditions in the previous five years (53.73% vs 51.65%) and an increased risk of mortality during the study period (0.46% vs 0.32%). The discrepancies between matched and unmatched individuals were more pronounced in the HD than SD cohort (Supplementary Table 4).

### rVE estimates and IRR

The influenza-related hospitalization rates were 125.8 per 100,000 person-years and 173.4 per 100,000 person-years in the HD cohort and the SD cohort respectively, converting to an estimated rVE of 27.39% [19.79;34.27]. The estimated rVE ranged from 19.07% when the ICD-10 discharge diagnosis code was less specific (i.e., PD, RD, or AD) to 29.91% when the analysis was restricted around the seasonal peak (Table 2).

**Table 2.** Incidence rate ratios and relative vaccine effectiveness of HD compared with SD. CI: confidence interval; HD: high-dose influenza vaccine; IRR: incidence rate ratio; rVE: relative vaccine effectiveness; SD: standard-dose influenza vaccine

| **Vaccine group** | **Hospitalization rate**<br>**per 100,000 person-years (95% CI)** | **IRR**<br>**(95% CI)** | **rVE (%)**<br>**(95% CI)** | **P-value** |
| --- | --- | --- | --- | --- |
| **Main analysis** |  |  |  |  |
| HD | 125.76 (115.37-137.10) | 0.73 [0.66;0.80] | 27.39 [19.79;34.27] | <0.0001 |
| SD | 173.36 (167.11-179.85) | 1.00 |  |  |
| **Sensitivity analysis: PD, RD, AD** |  |  |  |  |
| HD | 210.82 (197.23-225.35) | 0.81 [0.75;0.88] | 19.07 [12.37;25.27] | <0.0001 |
| SD | 260.69 (252.99-268.61) | 1.00 |  |  |
| **Sensitivity analysis: with a COVID-19 code** |  |  |  |  |
| HD | 127.96 (117.47-139.38) | 0.73 [0.66;0.80] | 27.14 [19.57;33.99] | <0.0001 |
| SD | 175.80 (169.50-182.33) | 1.00 |  |  |
| **Sensitivity analysis: at the peak of the season** |  |  |  |  |
| HD | 91.40 (82.60-101.13) | 0.70 [0.62;0.79] | 29.91 [21.22;37.65] | <0.0001 |
| SD | 130.68 (125.26-136.33) | 1.00 |  |  |

Non-influenza-specific hospitalizations (i.e., due to pneumonia, combined pneumonia/influenza, respiratory diseases, cardiovascular events, or cardiorespiratory events) are shown in (Figure 2). In the primary analysis (Figure 2 A), no significant differences were observed for most non-influenza-specific endpoints albeit for cardiovascular hospitalizations where event rates were higher in the HD than the SD cohort. Similar trends were observed in the sensibility analysis (Figure 2 B et C). During the peak season, statistically significant differences were observed for hospitalizations for combined pneumonia/influenza and hospitalizations for respiratory diseases (Figure 2 D).

**Figure 2.**
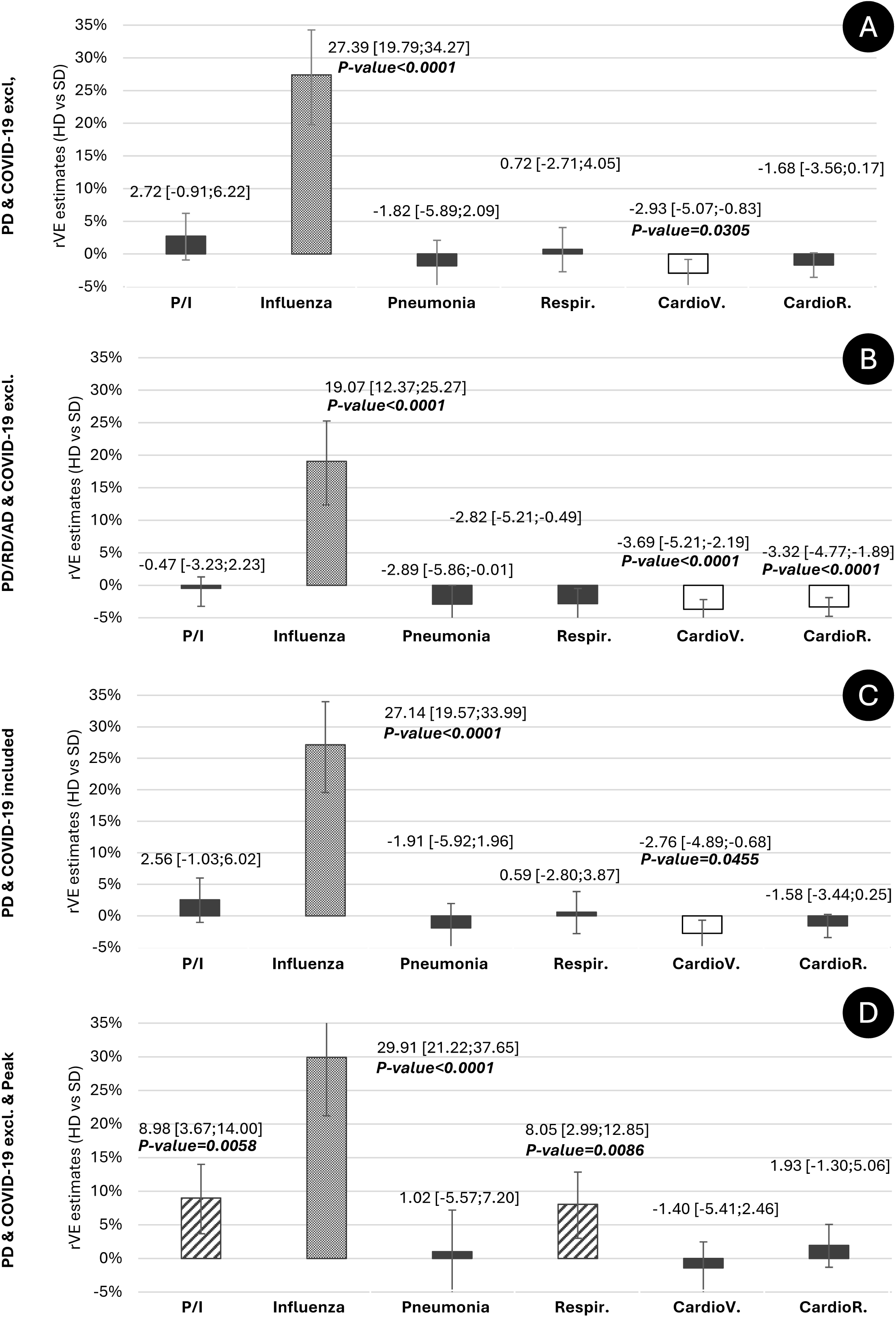
Relative vaccine effectiveness (rVE) of HD compared to SD during the 2022/23 influenza season. AD: associated diagnosis; CardioR: cardiorespiratory event; CardioV: cardiovascular event; excl.: excluded; HD: high-dose influenza vaccine; PD: primary diagnosis; P/I: pneumonia or influenza; RD: related diagnosis; Respir: respiratory event; SD: standard-dose influenza vaccine The ICD-10 discharge diagnosis code in the database could be noted as primary (PD), related (RD), or associated (AD). Values are Odds-ratios (OR) with 95% confidence intervals [95%CI]. Only p-values below 0.05 after Bonferroni-Holm correction are provided.

The estimated rVE for influenza-related hospitalizations varied from 22.65% to 33.55% across age groups, indicating that HD yielded better protection against influenza-related hospitalizations compared to SD, regardless of age (Table 3). The rVE was particularly high in the 85+ group. In this population, the influenza-related hospitalization rate was 279.90 per 100,000 person-years in the HD cohort and 419.76 per 100,000 person-years for the SD cohort, translating to an estimated rVE of 33.55%. Similar results were found at the peak of the season (85+, rVE=31.72%) or after the inclusion of hospitalizations with COVID-19 code (rVE=32.86%). When the ICD-10 discharge diagnosis code was less specific, the rVE was lower but remained above that observed for the whole population (rVE=25.03%) (Table 3).

**Table 3.**
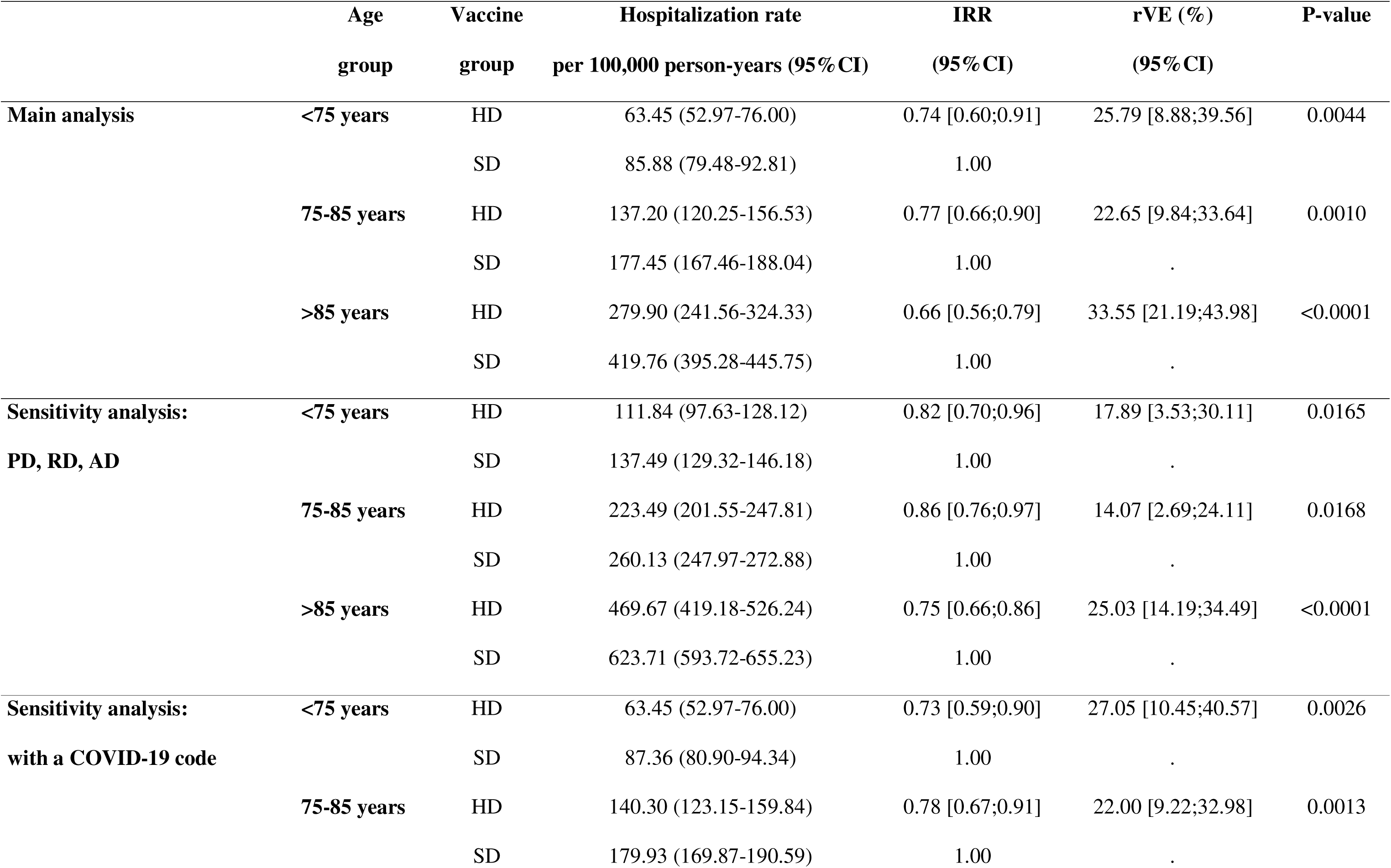
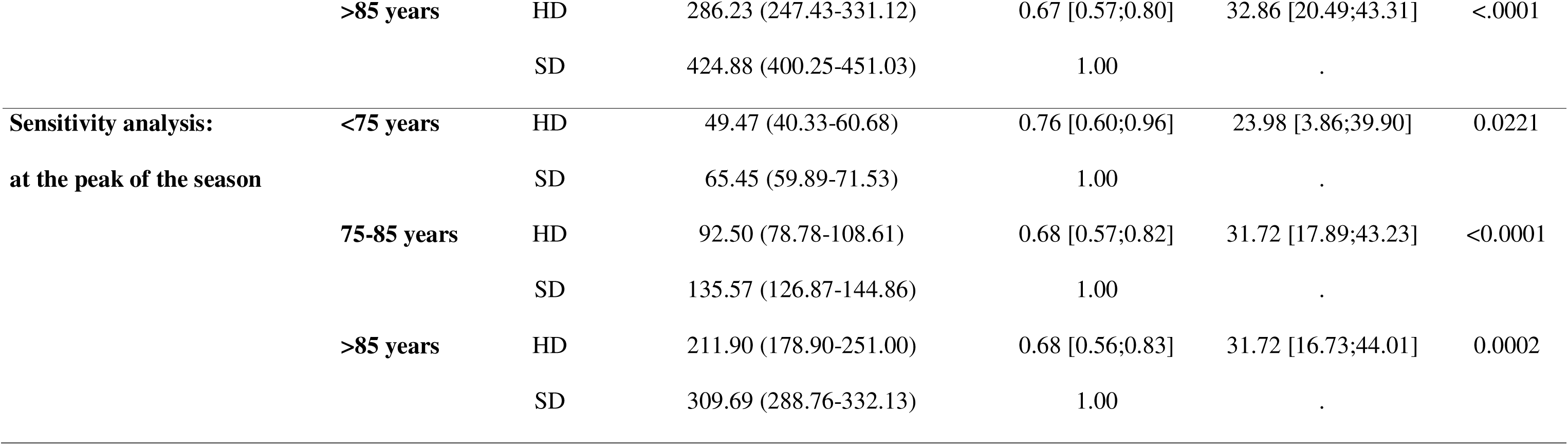
Incidence rate ratios and relative vaccine effectiveness of HD compared to SD according to the age group. CI: confidence interval; HD: high-dose influenza vaccine; IRR: incidence rate ratio; rVE: relative vaccine effectiveness; SD: standard-dose influenza vaccine

|  | Age | Vaccine | Hospitalization rate | IRR | rVE (%) | P-value |
| --- | --- | --- | --- | --- | --- | --- |
|  | group | group | per 100,000 person-years (95%CI) | (95%CI) | (95%CI) |  |
| **Main analysis** | **<75 years** | HD | 63.45 (52.97-76.00) | 0.74 [0.60;0.91] | 25.79 [8.88;39.56] | 0.0044 |
|  |  | SD | 85.88 (79.48-92.81) | 1.00 |  |  |
|  | **75-85 years** | HD | 137.20 (120.25-156.53) | 0.77 [0.66;0.90] | 22.65 [9.84;33.64] | 0.0010 |
|  |  | SD | 177.45 (167.46-188.04) | 1.00 | . |  |
|  | **>85 years** | HD | 279.90 (241.56-324.33) | 0.66 [0.56;0.79] | 33.55 [21.19;43.98] | <0.0001 |
|  |  | SD | 419.76 (395.28-445.75) | 1.00 | . |  |
| **Sensitivity analysis:**<br>**PD, RD, AD** | **<75 years** | HD | 111.84 (97.63-128.12) | 0.82 [0.70;0.96] | 17.89 [3.53;30.11] | 0.0165 |
|  |  | SD | 137.49 (129.32-146.18) | 1.00 | . |  |
|  | **75-85 years** | HD | 223.49 (201.55-247.81) | 0.86 [0.76;0.97] | 14.07 [2.69;24.11] | 0.0168 |
|  |  | SD | 260.13 (247.97-272.88) | 1.00 | . |  |
|  | **>85 years** | HD | 469.67 (419.18-526.24) | 0.75 [0.66;0.86] | 25.03 [14.19;34.49] | <0.0001 |
|  |  | SD | 623.71 (593.72-655.23) | 1.00 | . |  |
| **Sensitivity analysis:**<br>**with a COVID-19 code** | **<75 years** | HD | 63.45 (52.97-76.00) | 0.73 [0.59;0.90] | 27.05 [10.45;40.57] | 0.0026 |
|  |  | SD | 87.36 (80.90-94.34) | 1.00 | . |  |
|  | **75-85 years** | HD | 140.30 (123.15-159.84) | 0.78 [0.67;0.91] | 22.00 [9.22;32.98] | 0.0013 |
|  |  | SD | 179.93 (169.87-190.59) | 1.00 | . |  |
|  | >85 years | HD | 286.23 (247.43-331.12) | 0.67 [0.57;0.80] | 32.86 [20.49;43.31] | <.0001 |
|  |  | SD | 424.88 (400.25-451.03) | 1.00 | . |  |
| **Sensitivity analysis:**<br>**at the peak of the season** | <75 years | HD | 49.47 (40.33-60.68) | 0.76 [0.60;0.96] | 23.98 [3.86;39.90] | 0.0221 |
|  |  | SD | 65.45 (59.89-71.53) | 1.00 | . |  |
|  | 75-85 years | HD | 92.50 (78.78-108.61) | 0.68 [0.57;0.82] | 31.72 [17.89;43.23] | <0.0001 |
|  |  | SD | 135.57 (126.87-144.86) | 1.00 | . |  |
|  | >85 years | HD | 211.90 (178.90-251.00) | 0.68 [0.56;0.83] | 31.72 [16.73;44.01] | 0.0002 |
|  |  | SD | 309.69 (288.76-332.13) | 1.00 | . |  |

The IRR were 1.04 [1.00;1.07] against hospitalizations related to urinary tract infections (UTIs), 1.02 [1.00;1.03] against hospitalizations for cataract surgery, and 1.00 [0.93;1.08] against hospitalizations due to erysipelas, converting to rVEs of −3.52 [-7.00;-0.15] (p=0.0402), −1.71 [-3.22;-0.21] (p=0.0252), and −0.26 [-7.94;6.87] (p=0.9443), respectively, suggesting the existence of residual unmeasured confounding factors.

Considering multivariable analysis in the whole population (i.e., all 65+ vaccinated against influenza, N=7,676,621), the influenza-related hospitalization rate (DP only, COVID-19 excluded) was 151.44 [141.73;161.82] per 100,000 person-years in the HD full cohort and 154.67 [150.83;158.60] per 100,000 person-years for SD full cohort. Including the same covariables as in the matching procedure, the corresponding estimated adjusted rVE was very close to that obtained after matching: 27.42% [21.23;33.13] (p<0.001).

## Discussion

The results of this DRIVEN season 2 study reinforce those of season 1 [10], where HD was statistically significantly associated with fewer influenza-related hospitalizations but not with fewer non-specific influenza-related hospitalizations compared to SD in community-dwelling 65+ adults. Observational studies are inherently subject to bias, including residual bias due to unmeasured confounding. Therefore, their findings should not be regarded as conclusive without prior randomized controlled trials. Given that the benefits of HD against influenza and its severe complications compared to SD have been largely demonstrated in clinical trials and found consistently in other observational studies, we can have confidence in the findings related to influenza-related hospitalizations [5, 7].

Regarding non-influenza-specific hospitalizations, our findings differed from those reported in the literature [5, 7]. HD was not statistically significantly associated with a reduction in hospitalizations for pneumonia, combined pneumonia/influenza, or cardiorespiratory events. However, a benefit of HD over SDs in reducing hospitalizations for combined pneumonia/influenza and respiratory diseases was observed when the analysis focused on the peak of the influenza season. Residual bias due to unmeasured confounding may partially explain this discrepancy. The results of our falsification analysis supported this hypothesis. In assessing these less-specific endpoints, where relative effects could be smaller in magnitude, the ability to detect such effects could be obscured by such residual bias. This bias suggests that the benefits of HD vs SD on influenza-related hospitalizations observed in this study may be underestimated. The benefit of HD was robust, increasing the confidence in the findings. The benefits of HD in influenza-related hospitalization compared to SD were observed independently of the influenza diagnosis (primary, associated, related) and after the inclusion or exclusion of COVID-19 diagnosis. As expected, the highest VE was observed at the peak of the influenza season.

The rVE was consistent with the previous season and aligned with or surpassed estimates found in the literature [5, 10]. The difference between seasons 1 and 2 was partly explained by the number of individuals vaccinated with the HD (two times more individuals during the 2022/23 than the 2021/22 influenza vaccination campaign). The characteristics of the influenza epidemics in France also explained the difference between the two seasons. The 2021/22 influenza season which was due to the cocirculation of A(H3N2) and A (H1N1)pdm09 viruses was characterized by its late onset (March 2022), its short duration (9 weeks), and a low severity in 65+ whereas the 2022/23 influenza season, mainly related to A(H3N2) virus, was characterized by its early onset (November 2022), its long duration (19 weeks), and its high severity in the 65+ [15, 16]. In addition, due to the segmented RNA structure of influenza viruses and their high mutation rate, influenza vaccines require to be frequently updated to account for the latest antigenic and epidemiological changes. The composition of influenza vaccines is reassessed yearly based on information collected from national influenza reference centers and collaborating centers worldwide. Each year, the viral strains to be included in the vaccines are recommended by the World Health Organization (WHO) for each hemisphere. For the 2021/22 and 2022/23 seasons, the WHO recommended quadrivalent vaccines [17, 18]. As a consequence of the disappearance of the Yamagata lineage type B influenza, in 2020 and the recent recommendations from experts [19], trivalent vaccines are recommended for the 2024/25 influenza season [20].

Our results also showed that statistically significantly fewer influenza-related hospitalizations were consistently observed following HD than SD vaccination, with the rVE particularly high in the 85+ and maintained across all age groups. In light of these findings, one-third of this vulnerable population could have substantially benefited from HD vaccination and improved protection against influenza-related hospitalizations.

According to our study, the IRR for influenza-related hospitalizations in SD-vaccinated individuals was estimated at 173 for 100,000 person-years. Estimating that about 8 million individuals aged 65 years and over were vaccinated using the SD influenza vaccine during the 2022/23 influenza vaccination campaign, this would lead to approximately 12,000 influenza-related hospitalizations in this population. With an rVE of 27% for influenza-related hospitalizations, the replacement of the SD by the HD for the whole SD cohort would have avoided about 3,300 additional influenza-related hospitalizations over the few weeks of the season. According to a French study gathering data from eight epidemic seasons (i.e., 2010/11 to 2017/18) [21], the median length of hospital stay was 8 days for patients aged from 65 to 84 years and 10 days for those aged 85 years or more. The 3,300 influenza-related hospitalizations therefore represent about 29,000 days of hospitalization that could have been avoided by the use of HD instead of SD. It is to be noted that this estimate does not include the indirect costs and the consequences of hospitalization in older people or a better vaccine coverage rate. For the 2022/23 season, the vaccine coverage rate in 65+ was 54%, far below the French objective of 75% [22].

Our study had some limitations. Most of them were discussed in the article by Bricout et al. [10]. More specifically, the rate of matching in DRIVEN season 2 was lower than for season 1 (70.1% vs 99.2%), and 40% of 85+ HD-vaccinated were not matched with controls. Several reasons may explain these two observations, which are probably inter-related. Nearly twice as many HD doses were available on the French market in the 2022/23 season compared to the 2021/22 season. Therefore, as the matching ratio remained the same for both seasons (1:4), it was more difficult and sometimes impossible to find four controls within season 2. If a new study were to be conducted for the 2023/24 season or in countries where HD is more widely used, the matching ratio should be reconsidered. In addition, finding four controls fulfilling the matching criterion related to the index date (week of vaccination) was particularly difficult as the HD was mainly dispensed at the beginning of the season. Several hypotheses may explain the early dispensation of HD: (i) pharmacists knew that the vaccine was specifically designed for older adults and therefore prioritized it for the benefit of their older patients; (ii) patients had heard about this vaccine and were requesting it from their pharmacists; (iii) although in France all influenza vaccines are fully reimbursed once they are recommended (making the cost transparent to patients) pharmacists may have wanted to sell their stock of HD vaccines first since HD was more expensive for them to purchase; (iv) the older the individuals or the more comorbidities they had, the earlier they tended to get vaccinated. This latest hypothesis was partially supported by vaccine distribution data, which showed that 63.87% of the doses of influenza vaccine were dispensed within the first 4 weeks of the vaccination campaign in the 85+ compared to 58.15% in the 75-85 group, and 54.30% in the 65-75 group (Supplementary Table 3). Additionally, unmatched HD-vaccinated individuals were older and frailer than matched HD-vaccinated individuals. This indication bias possibly impacted the effectiveness of the HD and SD, but DRIVEN was not aimed at measuring the absolute effectiveness of each vaccine. On the contrary, this bias and the fact that the rVE was higher in the 85+ than in the 65-75 or 75-85 tended to indicate that the difference in effectiveness reported between the HD and SD was more perceptible in the oldest and most immunosenescent and that the indication bias possibly decreased the global rVE of the HD for the 2022/23 season.

Our study also had several strengths. DRIVEN season 2 was conducted across a large sample, including more than 2 million vaccinated individuals, capturing all HD provided free of charge in community pharmacies in France, giving a comprehensive overview of vaccine effectiveness. The large size of the study also allowed for subgroup analysis based on the age of vaccinated individuals. It is to be noted that the maintenance of the usual 5% significance level for tests and the large sample size require interpreting small statistically significant effects with caution. In addition, we conducted numerous sensitivity analyses and a falsification analysis to support the results of the main analysis. Finally, since the COVID-19 pandemic, the number of PCR tests in cases of suspected respiratory tract infection has increased, thereby improving the specificity of influenza coding at hospital discharge, and consecutively allowing for more accurate identification of influenza cases for the influenza hospitalization outcomes.

### Conclusion

This study provided the first published rVE data comparing HD vs SD for the season 2022-23 in France. It found that HD provided better protection against influenza-related hospitalizations compared to SD in older people in a real-world setting across all age groups (65-75 years, 75-85 years, and >85 years). Its results were consistent with those reported for the 2021/22 influenza season, but more robust given the larger number of HD-vaccinated individuals. These findings add to the six randomized controlled trials and 11 seasons of observational data showing the greater benefit of HD over SD. During the 2022/23 influenza season, in France, replacing SD with HD would have prevented about 29,000 hospital days. Finally, these data support the decision of health authorities who recommend its use in first intent or preferentially in older adults [23–25].

## Ethical approval and consent to participate

The authors confirm that all relevant ethical guidelines have been followed, and any necessary IRB and/or ethics committee approvals have been obtained.

## Consent to publication

All the authors consent to publication.

## Data and material availability

Additional information is available on justified request.

## Funding

The study was funded by Sanofi.

## Conflicts of interest

HB, MCL, MD, CS, MML are Sanofi employees and may hold shares in the company. NA, BG & FR are HEVA employees, which received funding from Sanofi to run the study.

PC reports to have participated in advisory committees organized by Sanofi and being a consultant for Sanofi.

JG reports to have participated in advisory committees organized by GSK, MSD, Pfizer, and Sanofi.

GG reports to have participated in advisory committees organized by Astellas, AstraZeneca, BioMerieux, MSD, Pfizer, Sanofi, Sanofi Pasteur, Sanofi Pasteur-MSD and Vifor, acted as consultant and speaker for these companies, and participated in congresses on invitation by Eisai, MSD, Novartis, Pfizer, Sanofi, and Vifor.

AM reports to have participated in advisory committees organized by Sanofi, Seqirus, and Viatris, to be a speaker for Sanofi, Seqirus, Viatris and Novavax, and to be a member of the scientific board of the SFM (formerly known as GEIG).

FP is the owner of Abelia Science, which received funding from Sanofi for medical writing. LW has received consulting fees from HEVA, Sanofi and Pfizer for works outside the submitted work.

OL reports to be a principal investigator in vaccine trials sponsored by Sanofi, MSD, Pfizer, GSK, Moderna. She received financial support for travel to medical congress and personal fees for participation in advisory boards for Sanofi, MSD, Pfizer, and GSK.

## Data Availability

Additional information is available on justified request.

## Acknowledgements

The authors would like to express their gratitude to Sacha Hiridjee at Heva for managing the data and Marion Fournier at Sanofi for her constant support in managing the study. They also extend their heartfelt thanks to the Caisse Nationale d’Assurance Maladie (CNAM) for providing the data that made this study possible.

MyPubli.online was used to collaborate and ChatGPT for translating some study documents and rephrasing some sentences.

## Authors’ contributions

The authors contributed to the conception, design, analysis, and review of this manuscript. The first two authors and the last author contributed more significantly to the writing and editing of the manuscript.

## List of abbreviations

AD: associated diagnosis
AIC: Akaike information criterion
ALD: long-term disease (Affection Longue Durée)
CI: confidence interval
COPD: chronic pulmonary obstructive disease
FDEp: French social deprivation index
FWER: Family-wise Error Rate
HD: high-dose influenza vaccine
ICD: International Classification of Diseases 10th Revision
IRR: incidence rate ratio
PD: primary diagnosis
RD: related diagnosis
rVE: relative vaccine effectiveness
SD: standard-dose influenza vaccine
SNDS: French Health Data System (Système National Des Données de Santé)
UTI: urinary tract infections
WHO: World Health Organization

## Additional Tables

**Supplementary Table 1. Medication codes, ICD-10 codes, and references used during the study**

**Supplementary Table 2. Characteristics of community-dwelling 65+ individuals vaccinated with HD or SD during the 2022/23 season (N=7,914,298)**

HD: high-dose influenza vaccine; SD: standard-dose influenza vaccine

**Supplementary Table 3. Characteristics of community-dwelling 65+ individuals vaccinated against influenza during the 2022/23 season, by age group (N=7,914,298)**

**Supplementary Table 4. Characteristics of matched and unmatched individuals vaccinated with HD or SD during the 2022/23 season (N=7,914,298)**

HD: high-dose influenza vaccine; SD: standard-dose influenza vaccine

## Additional Figures

**Supplementary Figure 1. HD and SD distribution over the influenza vaccine campaign (2022/23)**

HD: high-dose influenza vaccine; SD: standard-dose influenza vaccine

## Notes

### Author Declarations

The data supporting the study findings are part of the National Health Data System (SNDS, Systeme national des Donnees de Sante) and are available from the HDH (Health Data Hub https://www.health-data-hub.fr/). Restrictions apply to the availability of these data containing potentially identifying and sensitive patient information. Special permission to access these data for this study was granted by the ethics and scientific committee for health research, studies, and evaluations (CESREES, Comite Ethique et Scientifique pour les Recherches, les Etudes et les Evaluations dans le domaine de la Sante) (former CEREES) and the French data protection authority (Comite National de l'Informatique et des Libertes, CNIL). The study protocol obtained two consecutive authorizations from the French data protection authority CNIL (initial authorization: Decision No. DR-2022-049; substantial modifications authorization: Decision DR-2023-013). Informed consent was not required for the use of the anonymized secondary data, as mentioned in the Social Security Code, Article L161-28-1. All methods were performed in accordance with CNIL regulations and with REporting of studies Conducted using Observational Routinely-collected Data (RECORD) guidelines.

