## Supplemental Table 1 for "Relative effectiveness of high-dose vs standard-dose influenza vaccines in preventing hospitalizations: a national retrospective cohort study in France, 2022/23 season"

**Medication codes, ICD-10 codes, and references used during the study**

| Medication codes (UCD13 and CIP13) for SD and HD | | | |
| --- | --- | --- | --- |
| Vaccine | | **UCD13 code** | **CIP13 code** |
| Standard-dose | |  |  |
| - INFLUVAC TETRA SUSP INJ SER 0,5ML | | 3400894338703 | 3400930117712 |
| - VAXIGRIPTETRA SUSP INJ SER 0,5ML | | 3400894329657 | 3400928099877 |
|  | | 3400894329657 | 3400930067727 |
| High-Dose | |  |  |
| - EFLUELDA SUSP INJ SER VACCIN | | 3400890004602 | 3400930205372 |
|  |  |  | 3400930205389 |
|  |  |  | 3400930205426 |
|  |  |  | 3400930205402 |
|  |  |  | 3400930205396 |
|  |  |  | 3400930205419 |
| ICD-10 discharge codes for hospitalization | | | |
| Definition | **ICD-10 codes** | | |
| Influenza hospitalization | J09 to J11 | | |
| Pneumonia hospitalization | J12 to J18 | | |
| Respiratory hospitalization | J00 to J06, J09 to J18, J40-J41, J96 | | |
| Cardiovascular hospitalization | I16, I20 to I22, I24, I24, I26, I30, I40, I46 to I50, I63, I65-I66, G45-G46, J96 | | |
| Cardiorespiratory hospitalization | J00 to J06, J09 to J18, J40-J41, J96  I20 to I22, I24, I24, I26, I30, I40, I46 to I50, I63, I65-I66, G45-G46 | | |
| COVID-19 hospitalization | U071, U0710, U0711, U0714, U0715 | | |
| Urinary tract infection hospitalization | N410, N412-N413, N418, N419, N10, N110, N12, N136, N300, N309, T835 | | |
| Erysipelas hospitalization | A46 | | |
| Cataract hospitalization | BFGA427, BFGA004, BFPA002, BFGA368, BFGA008, BFGA002, BFGA003, BFGA010, BFGA006, GFGA009 | | |
| ICD-10 codes or references used to identify comorbidities | | | |
| Comorbidity | **Codes or references** | | |
| Diabetes | Rachas A, Gastaldi-Menager C, Denis P, Barthelemy P, Constantinou P, Drouin J, et al. The Economic Burden of Disease in France From the National Health Insurance Perspective: The Healthcare Expenditures and Conditions Mapping Used to Prepare the French Social Security Funding Act and the Public Health Act. Med Care. 2022;60(9):655-64. | | |
| Obesity and/or history of obesity surgery | HFCA001, HFCC003, HFFA001, HFFA011, HFFC004, HFFC018, HFGC900, HFKA001, HFKA002, HFKC001, HFLC900, HFLE002, HFMA009, HFMA010, HFMA011, HFMC006, HFMC007, HFMC008, HGCA009, HGCC027, E66 | | |
| Undernourishment/or history of undernourishment | E43, E44, E46 | | |
| COPD/Asthma | Rachas A, Gastaldi-Menager C, Denis P, Barthelemy P, Constantinou P, Drouin J, et al. The Economic Burden of Disease in France From the National Health Insurance Perspective: The Healthcare Expenditures and Conditions Mapping Used to Prepare the French Social Security Funding Act and the Public Health Act. Med Care. 2022;60(9):655-64. | | |
| Dementia | Rachas A, Gastaldi-Menager C, Denis P, Barthelemy P, Constantinou P, Drouin J, et al. The Economic Burden of Disease in France From the National Health Insurance Perspective: The Healthcare Expenditures and Conditions Mapping Used to Prepare the French Social Security Funding Act and the Public Health Act. Med Care. 2022;60(9):655-64. | | |
| Cardiovascular diseases | Rachas A, Gastaldi-Menager C, Denis P, Barthelemy P, Constantinou P, Drouin J, et al. The Economic Burden of Disease in France From the National Health Insurance Perspective: The Healthcare Expenditures and Conditions Mapping Used to Prepare the French Social Security Funding Act and the Public Health Act. Med Care. 2022;60(9):655-64. | | |
| Immunocompromised individuals | Rachas A, Gastaldi-Menager C, Denis P, Barthelemy P, Constantinou P, Drouin J, et al. The Economic Burden of Disease in France From the National Health Insurance Perspective: The Healthcare Expenditures and Conditions Mapping Used to Prepare the French Social Security Funding Act and the Public Health Act. Med Care. 2022;60(9):655-64.  And  Wyplosz B, Fernandes J, Goussiaume G, Moïsi J, Lortet-Tieulent J, Vainchtock A, et al. Adults at risk of pneumococcal disease in France. Infect Dis Now. 2021 Nov;51(8):661–6. | | |
| Chronic liver disease | Rachas A, Gastaldi-Menager C, Denis P, Barthelemy P, Constantinou P, Drouin J, et al. The Economic Burden of Disease in France From the National Health Insurance Perspective: The Healthcare Expenditures and Conditions Mapping Used to Prepare the French Social Security Funding Act and the Public Health Act. Med Care. 2022;60(9):655-64. | | |
| Terminal chronic kidney failure | Rachas A, Gastaldi-Menager C, Denis P, Barthelemy P, Constantinou P, Drouin J, et al. The Economic Burden of Disease in France From the National Health Insurance Perspective: The Healthcare Expenditures and Conditions Mapping Used to Prepare the French Social Security Funding Act and the Public Health Act. Med Care. 2022;60(9):655-64. | | |

HD: high-dose influenza vaccine; SD: standard-dose influenza vaccine
