## Supplemental Table 2 for "Relative effectiveness of high-dose vs standard-dose influenza vaccines in preventing hospitalizations: a national retrospective cohort study in France, 2022/23 season"

| **Variables** | **Modality** | **HD** | **SD** | **Total** |
| --- | --- | --- | --- | --- |
| **N** | **Total** | **976,211** | **6,938,087** | **7,914,298** |
| Age (years) at inclusion | N | 976,211 | 6,938,087 | 7,914,298 |
|  | Mean (±SD) | 77.75 (±7.82) | 75.87 (±7.57) | 76.10 (±7.62) |
|  | Min; Max | 65.00; 111.00 | 65.00; 111.00 | 65.00; 111.00 |
|  | Median | 77.00 | 75.00 | 75.00 |
|  | Q1; Q3 | 72.00; 84.00 | 70.00; 81.00 | 70.00; 81.00 |
| Age (years) at index date  (large categories) | <75 | 386,268 (39.57%) | 3,440,568 (49.59%) | 3,826,836 (48.35%) |
|  | 75-85 | 406,238 (41.61%) | 2,578,829 (37.17%) | 2,985,067 (37.72%) |
|  | >85 | 183,705 (18.82%) | 918,690 (13.24%) | 1,102,395 (13.93%) |
| Age (years) at index date (small categories) | <=70 | 199,474 (20.43%) | 1,977,330 (28.50%) | 2,176,804 (27.50%) |
|  | 71-75 | 237,021 (24.28%) | 1,828,952 (26.36%) | 2,065,973 (26.10%) |
|  | 76-80 | 195,329 (20.01%) | 1,307,808 (18.85%) | 1,503,137 (18.99%) |
|  | 81-85 | 160,682 (16.46%) | 905,307 (13.05%) | 1,065,989 (13.47%) |
|  | 86-90 | 118,702 (12.16%) | 613,338 (8.84%) | 732,040 (9.25%) |
|  | 91-95 | 52,247 (5.35%) | 250,023 (3.60%) | 302,270 (3.82%) |
|  | 96-100 | 11,587 (1.19%) | 50,662 (0.73%) | 62,249 (0.79%) |
|  | >100 | 1,169 (0.12%) | 4,667 (0.07%) | 5,836 (0.07%) |
| Gender | Male | 440,675 (45.14%) | 3,160,087 (45.55%) | 3,600,762 (45.50%) |
|  | Female | 535,536 (54.86%) | 3,778,000 (54.45%) | 4,313,536 (54.50%) |
| Region of  residence  at index date – (corrected) | Missing | 191 | 1,683 | 1,874 |
|  | Guadeloupe | 235 (0.02%) | 547 (0.01%) | 782 (0.01%) |
|  | Martinique | 47 (0.00%) | 73 (0.00%) | 120 (0.00%) |
|  | Guyane | 6 (0.00%) | 5 (0.00%) | 11 (0.00%) |
|  | La Réunion | 0 (0.00%) | 3 (0.00%) | 3 (0.00%) |
|  | Mayotte | 1 (0.00%) | 113 (0.00%) | 114 (0.00%) |
|  | DROM, other | 18 (0.00%) | 172 (0.00%) | 190 (0.00%) |
|  | Ile-de-France | 141,441 (14.49%) | 918,198 (13.24%) | 1,059,639 (13.39%) |
|  | Centre-Val de Loire | 51,178 (5.24%) | 303,460 (4.37%) | 354,638 (4.48%) |
|  | BFC | 36,554 (3.75%) | 335,480 (4.84%) | 372,034 (4.70%) |
|  | Normandie | 71,843 (7.36%) | 385,564 (5.56%) | 457,407 (5.78%) |
|  | Hauts-de-France | 99,006 (10.14%) | 581,420 (8.38%) | 680,426 (8.60%) |
|  | Grand Est | 108,550 (11.12%) | 556,342 (8.02%) | 664,892 (8.40%) |
|  | Pays de la Loire | 49,927 (5.12%) | 452,027 (6.52%) | 501,954 (6.34%) |
|  | Bretagne | 48,181 (4.94%) | 443,465 (6.39%) | 491,646 (6.21%) |
|  | Nouvelle Aquitaine | 103,829 (10.64%) | 788,193 (11.36%) | 892,022 (11.27%) |
|  | Occitanie | 83,175 (8.52%) | 710,217 (10.24%) | 793,392 (10.03%) |
|  | AURA | 109,488 (11.22%) | 813,839 (11.73%) | 923,327 (11.67%) |
|  | PACA | 59,677 (6.11%) | 588,769 (8.49%) | 648,446 (8.20%) |
|  | Corse | 2,607 (0.27%) | 36,552 (0.53%) | 39,159 (0.49%) |
|  | DROM, unspecified precision | 10,249 (1.05%) | 21,919 (0.32%) | 32,168 (0.41%) |
|  | COM, unspecified | 8 (0.00%) | 46 (0.00%) | 54 (0.00%) |
| Presence of C2S - Solidarity health insurance free of charge (formerly CMU-C) or with financial participation (formerly ACS) | No | 942,550 (96.55%) | 6,714,220 (96.77%) | 7,656,770 (96.75%) |
|  | Yes | 33,661 (3.45%) | 223,867 (3.23%) | 257,528 (3.25%) |
| Solidarity health insurance free of charge (formerly CMU-C) or with financial participation (formerly ACS) | No | 942,550 | 6,714,220 | 7,656,770 |
|  | Free | 9,420 (27.98%) | 62,156 (27.76%) | 71,576 (27.79%) |
|  | Unknown | 1,152 (3.42%) | 8,763 (3.91%) | 9,915 (3.85%) |
|  | Participatory | 23,089 (68.59%) | 152,948 (68.32%) | 176,037 (68.36%) |
| French social deprivation index (FDep) (quintile) | Missing | 34,118 | 202,861 | 236,979 |
|  | 1 | 187,698 (19.92%) | 1,278,287 (18.98%) | 1,465,985 (19.10%) |
|  | 2 | 183,565 (19.48%) | 1,322,423 (19.63%) | 1,505,988 (19.62%) |
|  | 3 | 192,478 (20.43%) | 1,388,236 (20.61%) | 1,580,714 (20.59%) |
|  | 4 | 190,465 (20.22%) | 1,425,704 (21.17%) | 1,616,169 (21.05%) |
|  | 5 | 187,887 (19.94%) | 1,320,576 (19.61%) | 1,508,463 (19.65%) |
| Local Potential Accessibility to General Practitioner (LPA) (corrected) (quintile) | Missing | 13,074 | 60,483 | 73,557 |
|  | 1 | 199,861 (20.75%) | 1,284,280 (18.67%) | 1,484,141 (18.93%) |
|  | 2 | 178,152 (18.50%) | 1,459,818 (21.23%) | 1,637,970 (20.89%) |
|  | 3 | 211,908 (22.00%) | 1,418,503 (20.62%) | 1,630,411 (20.79%) |
|  | 4 | 196,250 (20.38%) | 1,424,652 (20.71%) | 1,620,902 (20.67%) |
|  | 5 | 176,966 (18.37%) | 1,290,351 (18.76%) | 1,467,317 (18.71%) |
| Patient's social security regimen | CANSSM | 6,200 (0.64%) | 30,337 (0.44%) | 36,537 (0.46%) |
|  | CAVIMAC | 1,327 (0.14%) | 6,961 (0.10%) | 8,288 (0.10%) |
|  | CNMSS | 12,978 (1.33%) | 91,677 (1.32%) | 104,655 (1.32%) |
|  | CRPCEN | 1,677 (0.17%) | 13,533 (0.20%) | 15,210 (0.19%) |
|  | ENIM | 2,006 (0.21%) | 16,690 (0.24%) | 18,696 (0.24%) |
|  | MSA | 57,800 (5.92%) | 462,239 (6.66%) | 520,039 (6.57%) |
|  | Other | 73 (0.01%) | 598 (0.01%) | 671 (0.01%) |
|  | RATP | 1,528 (0.16%) | 11,234 (0.16%) | 12,762 (0.16%) |
|  | RG | 882,077 (90.36%) | 6,229,047 (89.78%) | 7,111,124 (89.85%) |
|  | RSI | 165 (0.02%) | 1,345 (0.02%) | 1,510 (0.02%) |
|  | SNCF | 10,380 (1.06%) | 74,426 (1.07%) | 84,806 (1.07%) |
| Patients with an LTD status | No | 468,102 (47.95%) | 3,629,650 (52.31%) | 4,097,752 (51.78%) |
|  | Yes | 508,109 (52.05%) | 3,308,437 (47.69%) | 3,816,546 (48.22%) |
| Nb of LTDs | N | 976,211 | 6,938,087 | 7,914,298 |
|  | Mean (±SD) | 0.73 (±0.85) | 0.65 (±0.81) | 0.66 (±0.81) |
|  | Min; Max | 0.00; 8.00 | 0.00; 8.00 | 0.00; 8.00 |
|  | Median | 1.00 | 0.00 | 0.00 |
|  | Q1; Q3 | 0.00; 1.00 | 0.00; 1.00 | 0.00; 1.00 |
| Nb of LTDs among patients having at least 1 LTD | Missing | 468,102 | 3,629,650 | 4,097,752 |
|  | 1 | 350,433 (68.97%) | 2,357,131 (71.25%) | 2,707,564 (70.94%) |
|  | 2 or more | 157,676 (31.03%) | 951,306 (28.75%) | 1,108,982 (29.06%) |
| Nb of all-cause hospitalizations* | N | 976,211 | 6,938,087 | 7,914,298 |
|  | Mean (±SD) | 0.15 (±1.10) | 0.11 (±0.89) | 0.12 (±0.92) |
|  | Min; Max | 0.00; 64.00 | 0.00; 70.00 | 0.00; 70.00 |
|  | Median | 0.00 | 0.00 | 0.00 |
|  | Q1; Q3 | 0.00; 0.00 | 0.00; 0.00 | 0.00; 0.00 |
| Nb of all-cause hospitalizations* - categorical | None | 900,731 (92.27%) | 6,509,890 (93.83%) | 7,410,621 (93.64%) |
|  | 1 | 55,555 (5.69%) | 322,890 (4.65%) | 378,445 (4.78%) |
|  | 2 | 11,408 (1.17%) | 61,116 (0.88%) | 72,524 (0.92%) |
|  | 3 | 3,300 (0.34%) | 17,691 (0.25%) | 20,991 (0.27%) |
|  | 4 | 1,559 (0.16%) | 7,805 (0.11%) | 9,364 (0.12%) |
|  | 5 | 865 (0.09%) | 4,421 (0.06%) | 5,286 (0.07%) |
|  | 6 | 507 (0.05%) | 2,549 (0.04%) | 3,056 (0.04%) |
|  | 7 | 280 (0.03%) | 1,403 (0.02%) | 1,683 (0.02%) |
|  | 8 | 211 (0.02%) | 1,090 (0.02%) | 1,301 (0.02%) |
|  | 9 | 140 (0.01%) | 664 (0.01%) | 804 (0.01%) |
|  | 10 or more | 1,655 (0.17%) | 8,568 (0.12%) | 10,223 (0.13%) |
| GP visits* | N | 976,211 | 6,938,087 | 7,914,298 |
|  | Mean (±SD) | 5.98 (±4.59) | 5.57 (±4.29) | 5.62 (±4.33) |
|  | Min; Max | 0.00; 197.00 | 0.00; 339.00 | 0.00; 339.00 |
|  | Median | 5.00 | 5.00 | 5.00 |
|  | Q1; Q3 | 3.00; 8.00 | 3.00; 7.00 | 3.00; 7.00 |
| GP visits* - categorical | None | 58,978 (6.04%) | 432,082 (6.23%) | 491,060 (6.20%) |
|  | 1 | 42,086 (4.31%) | 357,071 (5.15%) | 399,157 (5.04%) |
|  | 2 | 73,368 (7.52%) | 605,391 (8.73%) | 678,759 (8.58%) |
|  | 3 | 96,759 (9.91%) | 771,486 (11.12%) | 868,245 (10.97%) |
|  | 4 | 144,080 (14.76%) | 1,057,908 (15.25%) | 1,201,988 (15.19%) |
|  | 5 | 128,467 (13.16%) | 913,095 (13.16%) | 1,041,562 (13.16%) |
|  | 6 | 102,266 (10.48%) | 709,746 (10.23%) | 812,012 (10.26%) |
|  | 7 | 76,754 (7.86%) | 522,845 (7.54%) | 599,599 (7.58%) |
|  | 8 | 57,024 (5.84%) | 377,696 (5.44%) | 434,720 (5.49%) |
|  | 9 | 41,892 (4.29%) | 274,450 (3.96%) | 316,342 (4.00%) |
|  | 10 or more | 154,537 (15.83%) | 916,317 (13.21%) | 1,070,854 (13.53%) |
| Diabetes | No | 762,502 (78.11%) | 5,579,115 (80.41%) | 6,341,617 (80.13%) |
|  | Yes | 213,709 (21.89%) | 1,358,972 (19.59%) | 1,572,681 (19.87%) |
| Severe obesity | No | 889,864 (91.15%) | 6,399,958 (92.24%) | 7,289,822 (92.11%) |
|  | Yes | 86,347 (8.85%) | 538,129 (7.76%) | 624,476 (7.89%) |
| Severe malnutrition | No | 909,627 (93.18%) | 6,574,172 (94.75%) | 7,483,799 (94.56%) |
|  | Yes | 66,584 (6.82%) | 363,915 (5.25%) | 430,499 (5.44%) |
| COPD and  asthma | No | 853,604 (87.44%) | 6,150,428 (88.65%) | 7,004,032 (88.50%) |
|  | Yes | 122,607 (12.56%) | 787,659 (11.35%) | 910,266 (11.50%) |
| Dementia - neurological or degenerative disease | No | 944,223 (96.72%) | 6,784,399 (97.78%) | 7,728,622 (97.65%) |
|  | Yes | 31,988 (3.28%) | 153,688 (2.22%) | 185,676 (2.35%) |
| Cardiovascular disease | No | 690,918 (70.78%) | 5,140,810 (74.10%) | 5,831,728 (73.69%) |
|  | Yes | 285,293 (29.22%) | 1,797,277 (25.90%) | 2,082,570 (26.31%) |
| Myocardial infarction | No | 973,078 (99.68%) | 6,917,962 (99.71%) | 7,891,040 (99.71%) |
|  | Yes | 3,133 (0.32%) | 20,125 (0.29%) | 23,258 (0.29%) |
| Chronic coronary disease | No | 861,715 (88.27%) | 6,192,460 (89.25%) | 7,054,175 (89.13%) |
|  | Yes | 114,496 (11.73%) | 745,627 (10.75%) | 860,123 (10.87%) |
| Chronic heart failure | No | 941,188 (96.41%) | 6,741,863 (97.17%) | 7,683,051 (97.08%) |
|  | Yes | 35,023 (3.59%) | 196,224 (2.83%) | 231,247 (2.92%) |
| Occlusive arteriopathy of the lower limbs | No | 939,061 (96.19%) | 6,703,426 (96.62%) | 7,642,487 (96.57%) |
|  | Yes | 37,150 (3.81%) | 234,661 (3.38%) | 271,811 (3.43%) |
| Immuno-compromised subjects** | No | 786,443 (80.56%) | 5,662,422 (81.61%) | 6,448,865 (81.48%) |
|  | Yes | 189,768 (19.44%) | 1,275,665 (18.39%) | 1,465,433 (18.52%) |
| Cancer | No | 804,831 (82.44%) | 5,788,745 (83.43%) | 6,593,576 (83.31%) |
|  | Yes | 171,380 (17.56%) | 1,149,342 (16.57%) | 1,320,722 (16.69%) |
| Hematological tumors | No | 958,570 (98.19%) | 6,822,669 (98.34%) | 7,781,239 (98.32%) |
|  | Yes | 17,641 (1.81%) | 115,418 (1.66%) | 133,059 (1.68%) |
| Solid tumors | No | 818,835 (83.88%) | 5,880,523 (84.76%) | 6,699,358 (84.65%) |
|  | Yes | 157,376 (16.12%) | 1,057,564 (15.24%) | 1,214,940 (15.35%) |
| Solid organ transplant | No | 974,060 (99.78%) | 6,924,832 (99.81%) | 7,898,892 (99.81%) |
|  | Yes | 2,151 (0.22%) | 13,255 (0.19%) | 15,406 (0.19%) |
| Stem cells transplant | No | 975,564 (99.93%) | 6,933,325 (99.93%) | 7,908,889 (99.93%) |
|  | Yes | 647 (0.07%) | 4,762 (0.07%) | 5,409 (0.07%) |
| HIV patients | No | 974,249 (99.80%) | 6,924,713 (99.81%) | 7,898,962 (99.81%) |
|  | Yes | 1,962 (0.20%) | 13,374 (0.19%) | 15,336 (0.19%) |
| Patients with chronic autoimmune or inflammatory diseases*** | No | 957,601 (98.09%) | 6,810,664 (98.16%) | 7,768,265 (98.15%) |
|  | Yes | 18,610 (1.91%) | 127,423 (1.84%) | 146,033 (1.85%) |
| Chronic liver disease - diseases of the liver or pancreas | No | 960,504 (98.39%) | 6,832,351 (98.48%) | 7,792,855 (98.47%) |
|  | Yes | 15,707 (1.61%) | 105,736 (1.52%) | 121,443 (1.53%) |
| Severe renal disease | No | 971,268 (99.49%) | 6,908,027 (99.57%) | 7,879,295 (99.56%) |
|  | Yes | 4,943 (0.51%) | 30,060 (0.43%) | 35,003 (0.44%) |
| Nb of comorbidities of interest**** | N | 976,211 | 6,938,087 | 7,914,298 |
|  | Mean (±SD) | 1.04 (±1.08) | 0.92 (±1.03) | 0.94 (±1.04) |
|  | Min; Max | 0.00; 8.00 | 0.00; 8.00 | 0.00; 8.00 |
|  | Median | 1.00 | 1.00 | 1.00 |
|  | Q1; Q3 | 0.00; 2.00 | 0.00; 1.00 | 0.00; 1.00 |
| Nb of comorbidities of interest****- categorical | None | 369,356 (37.84%) | 2,965,797 (42.75%) | 3,335,153 (42.14%) |
|  | 1 | 333,687 (34.18%) | 2,305,777 (33.23%) | 2,639,464 (33.35%) |
|  | 2 or more | 273,168 (27.98%) | 1,666,513 (24.02%) | 1,939,681 (24.51%) |
| Nb of comorbidities of interest (detailed) | N | 976,211 | 6,938,087 | 7,914,298 |
|  | Mean (±SD) | 0.95 (±1.10) | 0.85 (±1.04) | 0.86 (±1.05) |
|  | Min; Max | 0.00; 10.00 | 0.00; 11.00 | 0.00; 11.00 |
|  | Median | 1.00 | 1.00 | 1.00 |
|  | Q1; Q3 | 0.00; 1.00 | 0.00; 1.00 | 0.00; 1.00 |
| Nb of comorbidities of interest (detailed) - categorical | None | 419,473 (42.97%) | 3,299,310 (47.55%) | 3,718,783 (46.99%) |
|  | 1 | 317,685 (32.54%) | 2,176,356 (31.37%) | 2,494,041 (31.51%) |
|  | 2 | 148,512 (15.21%) | 936,563 (13.50%) | 1,085,075 (13.71%) |
|  | 3 | 59,448 (6.09%) | 353,638 (5.10%) | 413,086 (5.22%) |
|  | 4 | 21,650 (2.22%) | 121,668 (1.75%) | 143,318 (1.81%) |
|  | 5 | 6,934 (0.71%) | 37,312 (0.54%) | 44,246 (0.56%) |
|  | 6 or more | 2,509 (0.26%) | 13,240 (0.19%) | 15,749 (0.20%) |
| Charlson comorbidity index (unadjusted for age) at index date | N | 976,211 | 6,938,087 | 7,914,298 |
|  | Mean (±SD) | 0.53 (±0.79) | 0.48 (±0.75) | 0.48 (±0.75) |
|  | Min; Max | 0.00; 10.00 | 0.00; 9.00 | 0.00; 10.00 |
|  | Median | 0.00 | 0.00 | 0.00 |
|  | Q1; Q3 | 0.00; 1.00 | 0.00; 1.00 | 0.00; 1.00 |
| Follow-up duration (in days) | N | 976,211 | 6,938,087 | 7,914,298 |
|  | Mean (±SD) | 238.66 (±26.87) | 229.02 (±25.77) | 230.21 (±26.10) |
|  | Min; Max | 1.00; 301.00 | 1.00; 302.00 | 1.00; 302.00 |
|  | Median | 247.00 | 233.00 | 234.00 |
|  | Q1; Q3 | 234.00; 254.00 | 218.00; 248.00 | 219.00; 249.00 |
| Reason for the end of follow up | Admission into an ESMS (EHPAD excluded) | 176 (0.02%) | 879 (0.01%) | 1,055 (0.01%) |
|  | Admission into an EHPAD | 5,378 (0.55%) | 25,668 (0.37%) | 31,046 (0.39%) |
|  | Death | 20,605 (2.11%) | 101,369 (1.46%) | 121,974 (1.54%) |
|  | End of follow-up | 950,052 (97.32%) | 6,810,171 (98.16%) | 7,760,223 (98.05%) |
| Death during follow-up | No | 955,616 (97.89%) | 6,836,766 (98.54%) | 7,792,382 (98.46%) |
|  | Yes | 20,595 (2.11%) | 101,321 (1.46%) | 121,916 (1.54%) |
| In-hospital death during study period | No | 960,862 (98.43%) | 6,859,204 (98.86%) | 7,820,066 (98.81%) |
|  | Yes | 15,349 (1.57%) | 78,883 (1.14%) | 94,232 (1.19%) |
| Field in which the death occurred (MCO, HAD, SSR or PSY) | HAD | 1,531 (0.16%) | 7,167 (0.10%) | 8,698 (0.11%) |
|  | MCO | 12,620 (1.29%) | 65,826 (0.95%) | 78,446 (0.99%) |
|  | No death | 960,862 (98.43%) | 6,859,204 (98.86%) | 7,820,066 (98.81%) |
|  | PSY | 4 (0.00%) | 58 (0.00%) | 62 (0.00%) |
|  | SSR | 1,194 (0.12%) | 5,832 (0.08%) | 7,026 (0.09%) |
| Last DP/DR of RUM is a P/I code | No | 976,163 (100.00%) | 6,937,868 (100.00%) | 7,914,031 (100.00%) |
|  | Yes | 48 (0.00%) | 219 (0.00%) | 267 (0.00%) |
| Last DP/DR of RUM is a cardiovascular code | No | 976,211 (100.00%) | 6,938,087 (100.00%) | 7,914,298 (100.00%) |
| Last DP/DR of RUM is a respiratory code | No | 976,211 (100.00%) | 6,938,084 (100.00%) | 7,914,295 (100.00%) |
|  | Yes | 0 (0.00%) | 3 (0.00%) | 3 (0.00%) |
| Last DP of RUM associated with a hospital death | I5009 | 281 (0.03%) | 1,277 (0.02%) | 1,558 (0.02%) |
|  | I5019 | 159 (0.02%) | 802 (0.01%) | 961 (0.01%) |
|  | J181 | 119 (0.01%) | 627 (0.01%) | 746 (0.01%) |
|  | J189 | 145 (0.01%) | 747 (0.01%) | 892 (0.01%) |
|  | J690 | 324 (0.03%) | 1,437 (0.02%) | 1,761 (0.02%) |
|  | J9600 | 230 (0.02%) | 973 (0.01%) | 1,203 (0.02%) |
|  | Other | 972,479 (99.62%) | 6,918,329 (99.72%) | 7,890,808 (99.70%) |
|  | R53+0 | 169 (0.02%) | 886 (0.01%) | 1,055 (0.01%) |
|  | R572 | 209 (0.02%) | 1,033 (0.01%) | 1,242 (0.02%) |
|  | U0710 | 303 (0.03%) | 1,577 (0.02%) | 1,880 (0.02%) |
|  | Z515 | 1,793 (0.18%) | 10,399 (0.15%) | 12,192 (0.15%) |
| Last DR of RUM associated with a hospital death | C189 | 28 (0.00%) | 103 (0.00%) | 131 (0.00%) |
|  | C20 | 30 (0.00%) | 188 (0.00%) | 218 (0.00%) |
|  | C220 | 51 (0.01%) | 305 (0.00%) | 356 (0.00%) |
|  | C221 | 19 (0.00%) | 136 (0.00%) | 155 (0.00%) |
|  | C250 | 37 (0.00%) | 313 (0.00%) | 350 (0.00%) |
|  | C259 | 28 (0.00%) | 188 (0.00%) | 216 (0.00%) |
|  | C341 | 51 (0.01%) | 370 (0.01%) | 421 (0.01%) |
|  | C343 | 21 (0.00%) | 183 (0.00%) | 204 (0.00%) |
|  | C349 | 101 (0.01%) | 641 (0.01%) | 742 (0.01%) |
|  | C509 | 55 (0.01%) | 313 (0.00%) | 368 (0.00%) |
|  | C56 | 41 (0.00%) | 227 (0.00%) | 268 (0.00%) |
|  | C61 | 85 (0.01%) | 439 (0.01%) | 524 (0.01%) |
|  | C64 | 25 (0.00%) | 182 (0.00%) | 207 (0.00%) |
|  | C679 | 41 (0.00%) | 247 (0.00%) | 288 (0.00%) |
|  | C920 | 24 (0.00%) | 154 (0.00%) | 178 (0.00%) |
|  | N185 | 23 (0.00%) | 165 (0.00%) | 188 (0.00%) |
|  | Other | 975,551 (99.93%) | 6,933,933 (99.94%) | 7,909,484 (99.94%) |
| Type of vaccine at index date | HD-QIV | 976,211 (100.00%) | 0 (0.00%) | 976,211 (12.33%) |
|  | SD-QIV | 0 (0.00%) | 6,938,087 (100.00%) | 6,938,087 (87.67%) |
| ~~Vaccine brand at index date~~ | ~~Efluelda~~ | ~~976,211 (100.00%)~~ | ~~0 (0.00%)~~ | ~~976,211 (12.33%)~~ |
|  | ~~Fluarix Tetra~~ | ~~0 (0.00%)~~ | ~~358,614 (5.17%)~~ | ~~358,614 (4.53%)~~ |
|  | ~~Influvac Tetra~~ | ~~0 (0.00%)~~ | ~~3,628,035 (52.29%)~~ | ~~3,628,035 (45.84%)~~ |
|  | ~~Vaxigrip Tetra~~ | ~~0 (0.00%)~~ | ~~2,951,438 (42.54%)~~ | ~~2,951,438 (37.29%)~~ |
| Vaccine administration by pharmacist | No | 506,363 (51.87%) | 3,283,652 (47.33%) | 3,790,015 (47.89%) |
|  | Yes | 469,848 (48.13%) | 3,654,435 (52.67%) | 4,124,283 (52.11%) |
| Week of vaccination | Week 1: 35 | 1 (0.00%) | 8 (0.00%) | 9 (0.00%) |
|  | Week 2: 36 | 0 (0.00%) | 40 (0.00%) | 40 (0.00%) |
|  | Week 3: 37 | 5 (0.00%) | 147 (0.00%) | 152 (0.00%) |
|  | Week 4: 38 | 31 (0.00%) | 231 (0.00%) | 262 (0.00%) |
|  | Week 5: 39 | 83 (0.01%) | 1,114 (0.02%) | 1,197 (0.02%) |
|  | Week 6: 40 | 732 (0.07%) | 3,759 (0.05%) | 4,491 (0.06%) |
|  | Week 7: 41 | 1,583 (0.16%) | 7,472 (0.11%) | 9,055 (0.11%) |
|  | Week 8: 42 | 346,309 (35.47%) | 1,291,737 (18.62%) | 1,638,046 (20.70%) |
|  | Week 9: 43 | 197,568 (20.24%) | 846,779 (12.20%) | 1,044,347 (13.20%) |
|  | Week 10: 44 | 131,128 (13.43%) | 728,355 (10.50%) | 859,483 (10.86%) |
|  | Week 11: 45 | 113,718 (11.65%) | 862,644 (12.43%) | 976,362 (12.34%) |
|  | Week 12: 46 | 80,912 (8.29%) | 895,344 (12.90%) | 976,256 (12.34%) |
|  | Week 13: 47 | 44,844 (4.59%) | 765,589 (11.03%) | 810,433 (10.24%) |
|  | Week 14: 48 | 27,446 (2.81%) | 699,336 (10.08%) | 726,782 (9.18%) |
|  | Week 15: 49 | 12,388 (1.27%) | 394,248 (5.68%) | 406,636 (5.14%) |
|  | Week 16: 50 | 6,333 (0.65%) | 192,756 (2.78%) | 199,089 (2.52%) |
|  | Week 17: 51 | 3,913 (0.40%) | 87,867 (1.27%) | 91,780 (1.16%) |
|  | Week 18: 52 | 3,076 (0.32%) | 61,059 (0.88%) | 64,135 (0.81%) |
|  | Week 19: 1 | 2,579 (0.26%) | 46,039 (0.66%) | 48,618 (0.61%) |
|  | Week 20: 2 | 1,399 (0.14%) | 23,328 (0.34%) | 24,727 (0.31%) |
|  | Week 21: 3 | 829 (0.08%) | 12,734 (0.18%) | 13,563 (0.17%) |
|  | Week 22: 4 | 655 (0.07%) | 9,342 (0.13%) | 9,997 (0.13%) |
|  | Week 23: 5 | 367 (0.04%) | 4,723 (0.07%) | 5,090 (0.06%) |
|  | Week 24: 6 | 110 (0.01%) | 1,297 (0.02%) | 1,407 (0.02%) |
|  | Week 25: 7 | 67 (0.01%) | 779 (0.01%) | 846 (0.01%) |
|  | Week 26: 8 | 49 (0.01%) | 535 (0.01%) | 584 (0.01%) |
|  | Week 27: 9 | 31 (0.00%) | 447 (0.01%) | 478 (0.01%) |
|  | Week 28: 10 | 7 (0.00%) | 120 (0.00%) | 127 (0.00%) |
|  | Week 29: 11 | 14 (0.00%) | 88 (0.00%) | 102 (0.00%) |
|  | Week 30: 12 | 8 (0.00%) | 94 (0.00%) | 102 (0.00%) |
|  | Week 31: 13 | 26 (0.00%) | 76 (0.00%) | 102 (0.00%) |
| ~~Second brand of influenza vaccine received in the season~~ | ~~Missing~~ | ~~976,211~~ | ~~6,937,989~~ | ~~7,914,200~~ |
|  | ~~Influvac Tetra~~ | ~~0 (.%)~~ | ~~10 (10.20%)~~ | ~~10 (10.20%)~~ |
|  | ~~VaxigripTetra~~ | ~~0 (.%)~~ | ~~88 (89.80%)~~ | ~~88 (89.80%)~~ |
| Nb of seasons with flu vaccination ***** | N | 976,211 | 6,938,087 | 7,914,298 |
|  | Mean (±SD) | 3.34 (±1.09) | 3.10 (±1.25) | 3.13 (±1.23) |
|  | Min; Max | 0.00; 5.00 | 0.00; 5.00 | 0.00; 5.00 |
|  | Median | 4.00 | 4.00 | 4.00 |
|  | Q1; Q3 | 3.00; 4.00 | 2.00; 4.00 | 2.00; 4.00 |
| Nb of seasons with flu vaccination ***** - categorical | 0 | 32,320 (3.31%) | 414,187 (5.97%) | 446,507 (5.64%) |
|  | 1 | 52,155 (5.34%) | 530,945 (7.65%) | 583,100 (7.37%) |
|  | 2 | 116,902 (11.98%) | 1,047,328 (15.10%) | 1,164,230 (14.71%) |
|  | 3 | 120,713 (12.37%) | 929,597 (13.40%) | 1,050,310 (13.27%) |
|  | 4 | 654,120 (67.01%) | 4,016,027 (57.88%) | 4,670,147 (59.01%) |
|  | 5 | 1 (0.00%) | 3 (0.00%) | 4 (0.00%) |
| Type of flu vaccination in the previous season | HD | 157,889 (16.17%) | 220,883 (3.18%) | 378,772 (4.79%) |
|  | Not vaccinated | 69,705 (7.14%) | 770,251 (11.10%) | 839,956 (10.61%) |
|  | SD | 748,289 (76.65%) | 5,946,394 (85.71%) | 6,694,683 (84.59%) |
|  | both | 328 (0.03%) | 559 (0.01%) | 887 (0.01%) |
| Nb of flu vaccination ***** (dispensation) | N | 976,211 | 6,938,087 | 7,914,298 |
|  | Mean (±SD) | 3.35 (±1.10) | 3.11 (±1.26) | 3.14 (±1.24) |
|  | Min; Max | 0.00; 11.00 | 0.00; 16.00 | 0.00; 16.00 |
|  | Median | 4.00 | 4.00 | 4.00 |
|  | Q1; Q3 | 3.00; 4.00 | 2.00; 4.00 | 2.00; 4.00 |
| Nb of flu vaccinations ***** (dispensation) - categorical | 0 | 32,322 (3.31%) | 414,199 (5.97%) | 446,521 (5.64%) |
|  | 1 | 51,974 (5.32%) | 529,166 (7.63%) | 581,140 (7.34%) |
|  | 2 | 116,236 (11.91%) | 1,041,462 (15.01%) | 1,157,698 (14.63%) |
|  | 3 | 120,229 (12.32%) | 927,274 (13.36%) | 1,047,503 (13.24%) |
|  | 4 | 647,588 (66.34%) | 3,979,326 (57.35%) | 4,626,914 (58.46%) |
|  | 5 | 7,638 (0.78%) | 45,150 (0.65%) | 52,788 (0.67%) |
|  | 6 or more | 224 (0.02%) | 1,510 (0.02%) | 1,734 (0.02%) |
| Vaccination status for COVID-19 at index date | On-going | 1,433 (0.15%) | 9,537 (0.14%) | 10,970 (0.14%) |
|  | Primo-vaccination | 948,366 (97.15%) | 6,744,844 (97.21%) | 7,693,210 (97.21%) |
|  | Unvaccinated | 26,412 (2.71%) | 183,706 (2.65%) | 210,118 (2.65%) |
| Pneumococcal vaccination status ***** | No | 853,368 (87.42%) | 6,149,947 (88.64%) | 7,003,315 (88.49%) |
|  | Yes | 122,843 (12.58%) | 788,140 (11.36%) | 910,983 (11.51%) |
| UTI-related hospitalizations | No | 968,045 (99.16%) | 6,893,159 (99.35%) | 7,861,204 (99.33%) |
|  | Yes | 8,166 (0.84%) | 44,928 (0.65%) | 53,094 (0.67%) |
| Nb of UTI-related hospitalizations | N | 976,211 | 6,938,087 | 7,914,298 |
|  | Mean (±SD) | 0.01 (±0.11) | 0.01 (±0.10) | 0.01 (±0.10) |
|  | Min; Max | 0.00; 15.00 | 0.00; 16.00 | 0.00; 16.00 |
|  | Median | 0.00 | 0.00 | 0.00 |
|  | Q1; Q3 | 0.00; 0.00 | 0.00; 0.00 | 0.00; 0.00 |
| Nb of UTI-related hospitalizations in patients with at least 1 UTI hospitalization | Missing | 968,045 | 6,893,159 | 7,861,204 |
|  | N | 8,166 | 44,928 | 53,094 |
|  | Mean (±SD) | 1.12 (±0.42) | 1.12 (±0.41) | 1.12 (±0.41) |
|  | Min; Max | 1.00; 15.00 | 1.00; 16.00 | 1.00; 16.00 |
|  | Median | 1.00 | 1.00 | 1.00 |
|  | Q1; Q3 | 1.00; 1.00 | 1.00; 1.00 | 1.00; 1.00 |
| Erysipelas-related hospitalizations | No | 974,463 (99.82%) | 6,928,557 (99.86%) | 7,903,020 (99.86%) |
|  | Yes | 1,748 (0.18%) | 9,530 (0.14%) | 11,278 (0.14%) |
| Nb of Erysipelas-related hospitalizations | N | 976,211 | 6,938,087 | 7,914,298 |
|  | Mean (±SD) | 0.00 (±0.05) | 0.00 (±0.05) | 0.00 (±0.05) |
|  | Min; Max | 0.00; 5.00 | 0.00; 9.00 | 0.00; 9.00 |
|  | Median | 0.00 | 0.00 | 0.00 |
|  | Q1; Q3 | 0.00; 0.00 | 0.00; 0.00 | 0.00; 0.00 |
| Nb of Erysipelas-related hospitalizations in patients with at least one Erysipelas hospitalization | Missing | 974,463 | 6,928,557 | 7,903,020 |
|  | N | 1,748 | 9,530 | 11,278 |
|  | Mean (±SD) | 1.13 (±0.40) | 1.14 (±0.42) | 1.14 (±0.42) |
|  | Min; Max | 1.00; 5.00 | 1.00; 9.00 | 1.00; 9.00 |
|  | Median | 1.00 | 1.00 | 1.00 |
|  | Q1; Q3 | 1.00; 1.00 | 1.00; 1.00 | 1.00; 1.00 |
| Cataract-related hospitalizations | No | 943,764 (96.68%) | 6,721,745 (96.88%) | 7,665,509 (96.86%) |
|  | Yes | 32,447 (3.32%) | 216,342 (3.12%) | 248,789 (3.14%) |
| Nb of Cataract-related hospitalizations | N | 976,211 | 6,938,087 | 7,914,298 |
|  | Mean (±SD) | 0.05 (±0.29) | 0.05 (±0.28) | 0.05 (±0.28) |
|  | Min; Max | 0.00; 3.00 | 0.00; 5.00 | 0.00; 5.00 |
|  | Median | 0.00 | 0.00 | 0.00 |
|  | Q1; Q3 | 0.00; 0.00 | 0.00; 0.00 | 0.00; 0.00 |
| Nb of Cataract-related hospitalizations in patients with at least one Cataract hospitalization | Missing | 943,764 | 6,721,745 | 7,665,509 |
|  | N | 32,447 | 216,342 | 248,789 |
|  | Mean (±SD) | 1.52 (±0.50) | 1.51 (±0.50) | 1.51 (±0.50) |
|  | Min; Max | 1.00; 3.00 | 1.00; 5.00 | 1.00; 5.00 |
|  | Median | 2.00 | 2.00 | 2.00 |
|  | Q1; Q3 | 1.00; 2.00 | 1.00; 2.00 | 1.00; 2.00 |
| Presence of flu hospitalizations | No | 974,850 (99.86%) | 6,928,984 (99.87%) | 7,903,834 (99.87%) |
|  | Yes | 1,361 (0.14%) | 9,103 (0.13%) | 10,464 (0.13%) |
| Nb of flu hospitalizations | N | 976,211 | 6,938,087 | 7,914,298 |
|  | Mean (±SD) | 0.00 (±0.04) | 0.00 (±0.04) | 0.00 (±0.04) |
|  | Min; Max | 0.00; 4.00 | 0.00; 3.00 | 0.00; 4.00 |
|  | Median | 0.00 | 0.00 | 0.00 |
|  | Q1; Q3 | 0.00; 0.00 | 0.00; 0.00 | 0.00; 0.00 |
| Nb of flu hospitalizations in patients with at least 1 flu hospitalization | Missing | 974,850 | 6,928,984 | 7,903,834 |
|  | N | 1,361 | 9,103 | 10,464 |
|  | Mean (±SD) | 1.11 (±0.34) | 1.08 (±0.29) | 1.09 (±0.30) |
|  | Min; Max | 1.00; 4.00 | 1.00; 3.00 | 1.00; 4.00 |
|  | Median | 1.00 | 1.00 | 1.00 |
|  | Q1; Q3 | 1.00; 1.00 | 1.00; 1.00 | 1.00; 1.00 |
| Presence of pneumonia hospitalizations | No | 962,721 (98.62%) | 6,866,174 (98.96%) | 7,828,895 (98.92%) |
|  | Yes | 13,490 (1.38%) | 71,913 (1.04%) | 85,403 (1.08%) |
| Nb of pneumonia hospitalizations | N | 976,211 | 6,938,087 | 7,914,298 |
|  | Mean (±SD) | 0.02 (±0.14) | 0.01 (±0.12) | 0.01 (±0.12) |
|  | Min; Max | 0.00; 6.00 | 0.00; 9.00 | 0.00; 9.00 |
|  | Median | 0.00 | 0.00 | 0.00 |
|  | Q1; Q3 | 0.00; 0.00 | 0.00; 0.00 | 0.00; 0.00 |
| Nb of pneumonia hospitalizations in patients with at least 1 pneumonia hospitalization | Missing | 962,721 | 6,866,174 | 7,828,895 |
|  | N | 13,490 | 71,913 | 85,403 |
|  | Mean (±SD) | 1.15 (±0.42) | 1.14 (±0.40) | 1.14 (±0.40) |
|  | Min; Max | 1.00; 6.00 | 1.00; 9.00 | 1.00; 9.00 |
|  | Median | 1.00 | 1.00 | 1.00 |
|  | Q1; Q3 | 1.00; 1.00 | 1.00; 1.00 | 1.00; 1.00 |
| Presence of P/I hospitalizations | No | 961,709 (98.51%) | 6,859,209 (98.86%) | 7,820,918 (98.82%) |
|  | Yes | 14,502 (1.49%) | 78,878 (1.14%) | 93,380 (1.18%) |
| Nb of P/I hospitalizations | N | 976,211 | 6,938,087 | 7,914,298 |
|  | Mean (±SD) | 0.02 (±0.15) | 0.01 (±0.13) | 0.01 (±0.13) |
|  | Min; Max | 0.00; 6.00 | 0.00; 9.00 | 0.00; 9.00 |
|  | Median | 0.00 | 0.00 | 0.00 |
|  | Q1; Q3 | 0.00; 0.00 | 0.00; 0.00 | 0.00; 0.00 |
| Nb of P/I hospitalizations in patients with at least 1 P/I hospitalization | Missing | 961,709 | 6,859,209 | 7,820,918 |
|  | N | 14,502 | 78,878 | 93,380 |
|  | Mean (±SD) | 1.17 (±0.45) | 1.16 (±0.44) | 1.16 (±0.44) |
|  | Min; Max | 1.00; 6.00 | 1.00; 9.00 | 1.00; 9.00 |
|  | Median | 1.00 | 1.00 | 1.00 |
|  | Q1; Q3 | 1.00; 1.00 | 1.00; 1.00 | 1.00; 1.00 |
| Presence of respiratory disease hospitalizations | No | 955,247 (97.85%) | 6,824,358 (98.36%) | 7,779,605 (98.30%) |
|  | Yes | 20,964 (2.15%) | 113,729 (1.64%) | 134,693 (1.70%) |
| Nb of respiratory disease hospitalizations | N | 976,211 | 6,938,087 | 7,914,298 |
|  | Mean (±SD) | 0.03 (±0.19) | 0.02 (±0.16) | 0.02 (±0.17) |
|  | Min; Max | 0.00; 8.00 | 0.00; 9.00 | 0.00; 9.00 |
|  | Median | 0.00 | 0.00 | 0.00 |
|  | Q1; Q3 | 0.00; 0.00 | 0.00; 0.00 | 0.00; 0.00 |
| Nb of respiratory hospitalizations in patients with at least 1 respiratory hospitalization | Missing | 955,247 | 6,824,358 | 7,779,605 |
|  | N | 20,964 | 113,729 | 134,693 |
|  | Mean (±SD) | 1.20 (±0.52) | 1.19 (±0.51) | 1.19 (±0.51) |
|  | Min; Max | 1.00; 8.00 | 1.00; 9.00 | 1.00; 9.00 |
|  | Median | 1.00 | 1.00 | 1.00 |
|  | Q1; Q3 | 1.00; 1.00 | 1.00; 1.00 | 1.00; 1.00 |
| Presence of cardiovascular disease hospitalizations | No | 926,245 (94.88%) | 6,658,999 (95.98%) | 7,585,244 (95.84%) |
|  | Yes | 49,966 (5.12%) | 279,088 (4.02%) | 329,054 (4.16%) |
| Nb of cardiovascular disease hospitalizations | N | 976,211 | 6,938,087 | 7,914,298 |
|  | Mean (±SD) | 0.07 (±0.33) | 0.05 (±0.29) | 0.05 (±0.30) |
|  | Min; Max | 0.00; 12.00 | 0.00; 14.00 | 0.00; 14.00 |
|  | Median | 0.00 | 0.00 | 0.00 |
|  | Q1; Q3 | 0.00; 0.00 | 0.00; 0.00 | 0.00; 0.00 |
| Nb of cardiovascular hospitalizations in patients with at least 1 cardiovascular hospitalization | Missing | 926,245 | 6,658,999 | 7,585,244 |
|  | N | 49,966 | 279,088 | 329,054 |
|  | Mean (±SD) | 1.33 (±0.70) | 1.31 (±0.68) | 1.31 (±0.68) |
|  | Min; Max | 1.00; 12.00 | 1.00; 14.00 | 1.00; 14.00 |
|  | Median | 1.00 | 1.00 | 1.00 |
|  | Q1; Q3 | 1.00; 1.00 | 1.00; 1.00 | 1.00; 1.00 |
| Presence of cardiorespiratory disease hospitalizations | No | 921,330 (94.38%) | 6,630,072 (95.56%) | 7,551,402 (95.41%) |
|  | Yes | 54,881 (5.62%) | 308,015 (4.44%) | 362,896 (4.59%) |
| Nb of cardio-respiratory disease hospitalizations | N | 976,211 | 6,938,087 | 7,914,298 |
|  | Mean (±SD) | 0.09 (±0.46) | 0.07 (±0.40) | 0.07 (±0.41) |
|  | Min; Max | 0.00; 18.00 | 0.00; 18.00 | 0.00; 18.00 |
|  | Median | 0.00 | 0.00 | 0.00 |
|  | Q1; Q3 | 0.00; 0.00 | 0.00; 0.00 | 0.00; 0.00 |
| Nb of cardio-respiratory hospitalizations in patients with at least 1 cardio-respiratory hospitalization | Missing | 921,330 | 6,630,072 | 7,551,402 |
|  | N | 54,881 | 308,015 | 362,896 |
|  | Mean (±SD) | 1.67 (±1.07) | 1.63 (±1.03) | 1.63 (±1.03) |
|  | Min; Max | 1.00; 18.00 | 1.00; 18.00 | 1.00; 18.00 |
|  | Median | 1.00 | 1.00 | 1.00 |
|  | Q1; Q3 | 1.00; 2.00 | 1.00; 2.00 | 1.00; 2.00 |
| LTD ICD-10 and name | Other | 235,980 (31.85%) | 1,508,995 (32.08%) | 1,744,975 (32.05%) |
|  | E11 | 155,804 (21.03%) | 1,004,539 (21.36%) | 1,160,343 (21.31%) |
|  | I25 | 65,942 (8.90%) | 430,608 (9.16%) | 496,550 (9.12%) |
|  | I48 | 54,243 (7.32%) | 331,111 (7.04%) | 385,354 (7.08%) |
|  | C61 | 32,959 (4.45%) | 217,179 (4.62%) | 250,138 (4.59%) |
|  | C50 | 28,837 (3.89%) | 197,192 (4.19%) | 226,029 (4.15%) |
|  | I64 | 18,480 (2.49%) | 113,947 (2.42%) | 132,427 (2.43%) |
|  | I70 | 14,192 (1.92%) | 94,641 (2.01%) | 108,833 (2.00%) |
|  | I21 | 13,414 (1.81%) | 93,507 (1.99%) | 106,921 (1.96%) |
|  | I50 | 15,273 (2.06%) | 87,908 (1.87%) | 103,181 (1.90%) |
|  | N18 | 12,905 (1.74%) | 75,035 (1.60%) | 87,940 (1.62%) |
|  | I702 | 11,602 (1.57%) | 71,543 (1.52%) | 83,145 (1.53%) |
|  | F00 (G30.-) | 13,508 (1.82%) | 66,346 (1.41%) | 79,854 (1.47%) |
|  | C18 | 10,686 (1.44%) | 67,356 (1.43%) | 78,042 (1.43%) |
|  | I49 | 10,631 (1.43%) | 63,078 (1.34%) | 73,709 (1.35%) |
|  | G20 | 10,294 (1.39%) | 62,693 (1.33%) | 72,987 (1.34%) |
|  | E10 | 9,979 (1.35%) | 62,539 (1.33%) | 72,518 (1.33%) |
|  | C67 | 8,504 (1.15%) | 55,164 (1.17%) | 63,668 (1.17%) |
|  | I35 | 8,721 (1.18%) | 53,769 (1.14%) | 62,490 (1.15%) |
|  | I10 | 8,901 (1.20%) | 46,212 (0.98%) | 55,113 (1.01%) |

AURA: Auvergne-Rhône-Alpes (French region); BFC: Bourgogne-Franche-Comté (French region); DP: Primary diagnosis; DR: related diagnosis; DROM: French oversea department and region; EHPAD: nursing home (*Etablissement d'Hébergement pour Personnes Agées Dépendantes*); ESMS: medico-social housing establishment; Flu: influenza; HAD: homecare setting (*hospitalisation à domicile*); HD-QIV: high dose QIV; ICD-10: International Classification of Diseases, 10th Revision (ICD-10); LTD: long-term disease; Max: maximum; MCO: Medicine, Surgery, Obstetrics (*medicine, chirurgie, obstétrique*); Min: minimum; Nb: number; PACA: Provence-Alpes-Côte d'Azur (French region); P/I: pneumonia or influenza; PSY: psychiatry; Q1-Q3: first and third quartiles; QIV: quadrivalent influenza vaccine; RUM: summary of patient’s medical data in one unit (*résumé d’unité médicale*); SD: standard deviation; SD-QIV: standard dose QIV; SSR: after care and rehabilitation (*soins de suite et de réadaptation*); UTI: urinary tract infection

* in the past 12 months (proxy for health status); ** including cancer patients, organ transplant, HIV patients, and chronic autoimmune or inflammatory diseases treated with immune-suppressive or biologic drugs; *** treated by immunosuppressive or biologic drugs; **** major groups of comorbidities; ***** in the 5 preceding seasons
