## Supplemental Table 3 for "Relative effectiveness of high-dose vs standard-dose influenza vaccines in preventing hospitalizations: a national retrospective cohort study in France, 2022/23 season"

| **Variables** | **Modality** | **<75 years** | **75-85 years** | **>85 years** | **Total** |
| --- | --- | --- | --- | --- | --- |
| **N** | **Total** | **3,826,836** | **2,985,067** | **1,102,395** | **7,914,298** |
| Age at inclusion | N | 3,826,836 | 2,985,067 | 1,102,395 | 7,914,298 |
|  | Mean (±SD) | 69.74 (±2.83) | 79.25 (±3.19) | 89.67 (±3.24) | 76.10 (±7.62) |
|  | Min; Max | 65.00; 74.00 | 75.00; 85.00 | 86.00; 111.00 | 65.00; 111.00 |
|  | Median | 70.00 | 79.00 | 89.00 | 75.00 |
|  | Q1; Q3 | 67.00; 72.00 | 76.00; 82.00 | 87.00; 92.00 | 70.00; 81.00 |
| Age at index date (large categories) | <75 years | 3,826,836 (100.00%) | 0 (0.00%) | 0 (0.00%) | 3,826,836 (48.35%) |
|  | 75-85 years | 0 (0.00%) | 2,985,067 (100.00%) | 0 (0.00%) | 2,985,067 (37.72%) |
|  | >85 years | 0 (0.00%) | 0 (0.00%) | 1,102,395 (100.00%) | 1,102,395 (13.93%) |
| Age at index date (small categories) | <=70 years | 2,176,804 (56.88%) | 0 (0.00%) | 0 (0.00%) | 2,176,804 (27.50%) |
|  | **71-75 years** | **1,650,032 (43.12%)** | **415,941 (13.93%)** | **0 (0.00%)** | **2,065,973 (26.10%)** |
|  | 76-80 years | 0 (0.00%) | 1,503,137 (50.36%) | 0 (0.00%) | 1,503,137 (18.99%) |
|  | 81-85 years | 0 (0.00%) | 1,065,989 (35.71%) | 0 (0.00%) | 1,065,989 (13.47%) |
|  | 86-90 years | 0 (0.00%) | 0 (0.00%) | 732,040 (66.40%) | 732,040 (9.25%) |
|  | 91-95 years | 0 (0.00%) | 0 (0.00%) | 302,270 (27.42%) | 302,270 (3.82%) |
|  | 96-100 years | 0 (0.00%) | 0 (0.00%) | 62,249 (5.65%) | 62,249 (0.79%) |
|  | >100 years | 0 (0.00%) | 0 (0.00%) | 5,836 (0.53%) | 5,836 (0.07%) |
| Gender | Men | 1,840,082 (48.08%) | 1,361,160 (45.60%) | 399,520 (36.24%) | 3,600,762 (45.50%) |
|  | Women | 1,986,754 (51.92%) | 1,623,907 (54.40%) | 702,875 (63.76%) | 4,313,536 (54.50%) |
| Region of residence at index date - Corrected | Missing | 1,092 | 665 | 117 | 1,874 |
|  | Guadeloupe | 214 (0.01%) | 375 (0.01%) | 193 (0.02%) | 782 (0.01%) |
|  | Martinique | 50 (0.00%) | 56 (0.00%) | 14 (0.00%) | 120 (0.00%) |
|  | Guyane | 6 (0.00%) | 5 (0.00%) | 0 (0.00%) | 11 (0.00%) |
|  | La Réunion | 3 (0.00%) | 0 (0.00%) | 0 (0.00%) | 3 (0.00%) |
|  | Mayotte | 86 (0.00%) | 26 (0.00%) | 2 (0.00%) | 114 (0.00%) |
|  | Autres DROM | 107 (0.00%) | 66 (0.00%) | 17 (0.00%) | 190 (0.00%) |
|  | Ile-de-France | 513,749 (13.43%) | 401,261 (13.45%) | 144,629 (13.12%) | 1,059,639 (13.39%) |
|  | Centre-Val de Loire | 170,703 (4.46%) | 133,161 (4.46%) | 50,774 (4.61%) | 354,638 (4.48%) |
|  | Bourgogne-Franche-Comté | 177,658 (4.64%) | 141,931 (4.76%) | 52,445 (4.76%) | 372,034 (4.70%) |
|  | Normandie | 230,761 (6.03%) | 165,531 (5.55%) | 61,115 (5.54%) | 457,407 (5.78%) |
|  | Hauts-de-France | 361,732 (9.46%) | 237,476 (7.96%) | 81,218 (7.37%) | 680,426 (8.60%) |
|  | Grand Est | 333,666 (8.72%) | 244,695 (8.20%) | 86,531 (7.85%) | 664,892 (8.40%) |
|  | Pays de la Loire | 248,522 (6.50%) | 185,680 (6.22%) | 67,752 (6.15%) | 501,954 (6.34%) |
|  | Bretagne | 244,149 (6.38%) | 181,477 (6.08%) | 66,020 (5.99%) | 491,646 (6.21%) |
|  | Nouvelle Aquitaine | 424,733 (11.10%) | 332,653 (11.15%) | 134,636 (12.21%) | 892,022 (11.27%) |
|  | Occitanie | 366,907 (9.59%) | 307,251 (10.30%) | 119,234 (10.82%) | 793,392 (10.03%) |
|  | Auvergne-Rhône-Alpes | 433,420 (11.33%) | 361,232 (12.10%) | 128,675 (11.67%) | 923,327 (11.67%) |
|  | Provence-Alpes-Côte d'Azur | 286,501 (7.49%) | 263,674 (8.84%) | 98,271 (8.92%) | 648,446 (8.20%) |
|  | Corse | 17,060 (0.45%) | 15,741 (0.53%) | 6,358 (0.58%) | 39,159 (0.49%) |
|  | DROM Sans precision | 15,685 (0.41%) | 12,092 (0.41%) | 4,391 (0.40%) | 32,168 (0.41%) |
|  | COM Sans precision | 32 (0.00%) | 19 (0.00%) | 3 (0.00%) | 54 (0.00%) |
| Presence of C2S - Solidarity health insurance free of charge (formerly CMU-C) or with financial participation (formerly ACS) | No | 3,669,585 (95.89%) | 2,910,884 (97.51%) | 1,076,301 (97.63%) | 7,656,770 (96.75%) |
|  | Yes | 157,251 (4.11%) | 74,183 (2.49%) | 26,094 (2.37%) | 257,528 (3.25%) |
| Solidarity health insurance free of charge (formerly CMU-C) or with financial participation (formerly ACS) | No | 3,669,585 | 2,910,884 | 1,076,301 | 7,656,770 |
|  | Free | 48,500 (30.84%) | 17,261 (23.27%) | 5,815 (22.28%) | 71,576 (27.79%) |
|  | Unknown if Free or Participatory | 4,435 (2.82%) | 2,873 (3.87%) | 2,607 (9.99%) | 9,915 (3.85%) |
|  | Participatory | 104,316 (66.34%) | 54,049 (72.86%) | 17,672 (67.72%) | 176,037 (68.36%) |
| French social deprivation index (FDep) (quintile) | Missing | 112,758 | 93,591 | 30,63 | 236,979 |
|  | 1st quintile | 704,882 (18.98%) | 564,684 (19.53%) | 196,419 (18.33%) | 1,465,985 (19.10%) |
|  | 2nd quintile | 744,325 (20.04%) | 565,683 (19.56%) | 195,980 (18.29%) | 1,505,988 (19.62%) |
|  | 3rd quintile | 763,950 (20.57%) | 595,131 (20.58%) | 221,633 (20.68%) | 1,580,714 (20.59%) |
|  | 4th quintile | 778,770 (20.97%) | 604,681 (20.91%) | 232,718 (21.71%) | 1,616,169 (21.05%) |
|  | 5th quintile | 722,151 (19.44%) | 561,297 (19.41%) | 225,015 (20.99%) | 1,508,463 (19.65%) |
| Local Potential Accessibility to General Practitioner (LPA) - Corrected | Missing | 34,061 | 28,609 | 10,887 | 73,557 |
|  | 1st quintile | 726,804 (19.16%) | 554,859 (18.77%) | 202,478 (18.55%) | 1,484,141 (18.93%) |
|  | 2nd quintile | 787,222 (20.76%) | 616,957 (20.87%) | 233,791 (21.42%) | 1,637,970 (20.89%) |
|  | 3rd quintile | 792,377 (20.89%) | 614,990 (20.80%) | 223,044 (20.43%) | 1,630,411 (20.79%) |
|  | 4th quintile | 786,964 (20.75%) | 611,119 (20.67%) | 222,819 (20.41%) | 1,620,902 (20.67%) |
|  | 5th quintile | 699,408 (18.44%) | 558,533 (18.89%) | 209,376 (19.18%) | 1,467,317 (18.71%) |
| Patient's social security regimen | CANSSM | 8,329 (0.22%) | 14,889 (0.50%) | 13,319 (1.21%) | 36,537 (0.46%) |
|  | CAVIMAC | 2,022 (0.05%) | 3,674 (0.12%) | 2,592 (0.24%) | 8,288 (0.10%) |
|  | CNMSS | 45,733 (1.20%) | 40,767 (1.37%) | 18,155 (1.65%) | 104,655 (1.32%) |
|  | CRPCEN | 6,966 (0.18%) | 6,016 (0.20%) | 2,228 (0.20%) | 15,210 (0.19%) |
|  | ENIM | 6,290 (0.16%) | 8,518 (0.29%) | 3,888 (0.35%) | 18,696 (0.24%) |
|  | MSA | 194,343 (5.08%) | 195,682 (6.56%) | 130,014 (11.79%) | 520,039 (6.57%) |
|  | Other | 384 (0.01%) | 270 (0.01%) | 17 (0.00%) | 671 (0.01%) |
|  | RATP | 5,357 (0.14%) | 5,259 (0.18%) | 2,146 (0.19%) | 12,762 (0.16%) |
|  | RG | 3,519,741 (91.98%) | 2,675,731 (89.64%) | 915,652 (83.06%) | 7,111,124 (89.85%) |
|  | RSI | 1,070 (0.03%) | 364 (0.01%) | 76 (0.01%) | 1,510 (0.02%) |
|  | SNCF | 36,601 (0.96%) | 33,897 (1.14%) | 14,308 (1.30%) | 84,806 (1.07%) |
| Patients with an LTD (long-term disease) status | No | 2,219,181 (57.99%) | 1,445,117 (48.41%) | 433,454 (39.32%) | 4,097,752 (51.78%) |
|  | Yes | 1,607,655 (42.01%) | 1,539,950 (51.59%) | 668,941 (60.68%) | 3,816,546 (48.22%) |
| Number of LTDs (long term disease) | N | 3,826,836 | 2,985,067 | 1,102,395 | 7,914,298 |
|  | Mean (±SD) | 0.55 (±0.75) | 0.72 (±0.84) | 0.88 (±0.89) | 0.66 (±0.81) |
|  | Min; Max | 0.00; 7.00 | 0.00; 8.00 | 0.00; 8.00 | 0.00; 8.00 |
|  | Median | 0.00 | 1.00 | 1.00 | 0.00 |
|  | Q1; Q3 | 0.00; 1.00 | 0.00; 1.00 | 0.00; 1.00 | 0.00; 1.00 |
| Number of LTDs among patients having at least one LTD | Missing | 2,219,181 | 1,445,117 | 433,454 | 4,097,752 |
|  | One LTD | 1,202,534 (74.80%) | 1,065,972 (69.22%) | 439,058 (65.63%) | 2,707,564 (70.94%) |
|  | Two or more LTDs | 405,121 (25.20%) | 473,978 (30.78%) | 229,883 (34.37%) | 1,108,982 (29.06%) |
| Number of all-cause hospitalization in the past 12 months (proxy for health status) | N | 3,826,836 | 2,985,067 | 1,102,395 | 7,914,298 |
|  | Mean (±SD) | 0.11 (±0.87) | 0.13 (±0.99) | 0.11 (±0.88) | 0.12 (±0.92) |
|  | Min; Max | 0.00; 70.00 | 0.00; 64.00 | 0.00; 53.00 | 0.00; 70.00 |
|  | Median | 0.00 | 0.00 | 0.00 | 0.00 |
|  | Q1; Q3 | 0.00; 0.00 | 0.00; 0.00 | 0.00; 0.00 | 0.00; 0.00 |
| Number of all-cause hospitalization in the past 12 months (proxy for health status) - categorical | None | 3,604,341 (94.19%) | 2,775,378 (92.98%) | 1,030,902 (93.51%) | 7,410,621 (93.64%) |
|  | 1 | 168,419 (4.40%) | 155,106 (5.20%) | 54,920 (4.98%) | 378,445 (4.78%) |
|  | 2 | 29,552 (0.77%) | 32,105 (1.08%) | 10,867 (0.99%) | 72,524 (0.92%) |
|  | 3 | 9,658 (0.25%) | 8,763 (0.29%) | 2,570 (0.23%) | 20,991 (0.27%) |
|  | 4 | 4,498 (0.12%) | 3,978 (0.13%) | 888 (0.08%) | 9,364 (0.12%) |
|  | 5 | 2,550 (0.07%) | 2,294 (0.08%) | 442 (0.04%) | 5,286 (0.07%) |
|  | 6 | 1,480 (0.04%) | 1,328 (0.04%) | 248 (0.02%) | 3,056 (0.04%) |
|  | 7 | 848 (0.02%) | 707 (0.02%) | 128 (0.01%) | 1,683 (0.02%) |
|  | 8 | 645 (0.02%) | 563 (0.02%) | 93 (0.01%) | 1,301 (0.02%) |
|  | 9 | 406 (0.01%) | 334 (0.01%) | 64 (0.01%) | 804 (0.01%) |
|  | 10 or more | 4,439 (0.12%) | 4,511 (0.15%) | 1,273 (0.12%) | 10,223 (0.13%) |
| General practitioner (GP) visits in the past 12 months (proxy for health status) | N | 3,826,836 | 2,985,067 | 1,102,395 | 7,914,298 |
|  | Mean (±SD) | 5.01 (±3.95) | 5.90 (±4.30) | 6.99 (±5.21) | 5.62 (±4.33) |
|  | Min; Max | 0.00; 216.00 | 0.00; 316.00 | 0.00; 339.00 | 0.00; 339.00 |
|  | Median | 4.00 | 5.00 | 6.00 | 5.00 |
|  | Q1; Q3 | 3.00; 7.00 | 3.00; 8.00 | 4.00; 9.00 | 3.00; 7.00 |
| General practitioner (GP) visits in the past 12 months (proxy for health status) - categorical | None | 294,421 (7.69%) | 149,230 (5.00%) | 47,409 (4.30%) | 491,060 (6.20%) |
|  | 1 | 252,417 (6.60%) | 118,390 (3.97%) | 28,350 (2.57%) | 399,157 (5.04%) |
|  | 2 | 401,892 (10.50%) | 223,452 (7.49%) | 53,415 (4.85%) | 678,759 (8.58%) |
|  | 3 | 472,208 (12.34%) | 308,600 (10.34%) | 87,437 (7.93%) | 868,245 (10.97%) |
|  | 4 | 588,522 (15.38%) | 462,313 (15.49%) | 151,153 (13.71%) | 1,201,988 (15.19%) |
|  | 5 | 487,913 (12.75%) | 410,294 (13.74%) | 143,355 (13.00%) | 1,041,562 (13.16%) |
|  | 6 | 367,036 (9.59%) | 323,589 (10.84%) | 121,387 (11.01%) | 812,012 (10.26%) |
|  | 7 | 261,429 (6.83%) | 242,133 (8.11%) | 96,037 (8.71%) | 599,599 (7.58%) |
|  | 8 | 182,846 (4.78%) | 178,420 (5.98%) | 73,454 (6.66%) | 434,720 (5.49%) |
|  | 9 | 129,758 (3.39%) | 129,884 (4.35%) | 56,700 (5.14%) | 316,342 (4.00%) |
|  | 10 or more | 388,394 (10.15%) | 438,762 (14.70%) | 243,698 (22.11%) | 1,070,854 (13.53%) |
| Diabetes | No | 3,082,272 (80.54%) | 2,353,043 (78.83%) | 906,302 (82.21%) | 6,341,617 (80.13%) |
|  | Yes | 744,564 (19.46%) | 632,024 (21.17%) | 196,093 (17.79%) | 1,572,681 (19.87%) |
| Severe obesity | No | 3,510,735 (91.74%) | 2,740,388 (91.80%) | 1,038,699 (94.22%) | 7,289,822 (92.11%) |
|  | Yes | 316,101 (8.26%) | 244,679 (8.20%) | 63,696 (5.78%) | 624,476 (7.89%) |
| Severe malnutrition | No | 3,730,816 (97.49%) | 2,810,791 (94.16%) | 942,192 (85.47%) | 7,483,799 (94.56%) |
|  | Yes | 96,020 (2.51%) | 174,276 (5.84%) | 160,203 (14.53%) | 430,499 (5.44%) |
| COPD and asthma | No | 3,403,396 (88.93%) | 2,634,333 (88.25%) | 966,303 (87.65%) | 7,004,032 (88.50%) |
|  | Yes | 423,440 (11.07%) | 350,734 (11.75%) | 136,092 (12.35%) | 910,266 (11.50%) |
| Dementia - neurological or degenerative disease | No | 3,806,898 (99.48%) | 2,902,920 (97.25%) | 1,018,804 (92.42%) | 7,728,622 (97.65%) |
|  | Yes | 19,938 (0.52%) | 82,147 (2.75%) | 83,591 (7.58%) | 185,676 (2.35%) |
| Cardiovascular disease | No | 3,098,184 (80.96%) | 2,100,067 (70.35%) | 633,477 (57.46%) | 5,831,728 (73.69%) |
|  | Yes | 728,652 (19.04%) | 885,000 (29.65%) | 468,918 (42.54%) | 2,082,570 (26.31%) |
| Myocardial infarction | No | 3,817,698 (99.76%) | 2,975,227 (99.67%) | 1,098,115 (99.61%) | 7,891,040 (99.71%) |
|  | Yes | 9,138 (0.24%) | 9,840 (0.33%) | 4,280 (0.39%) | 23,258 (0.29%) |
| Chronic coronary disease | No | 3,482,010 (90.99%) | 2,621,600 (87.82%) | 950,565 (86.23%) | 7,054,175 (89.13%) |
|  | Yes | 344,826 (9.01%) | 363,467 (12.18%) | 151,830 (13.77%) | 860,123 (10.87%) |
| Chronic heart failure | No | 3,770,253 (98.52%) | 2,893,043 (96.92%) | 1,019,755 (92.50%) | 7,683,051 (97.08%) |
|  | Yes | 56,583 (1.48%) | 92,024 (3.08%) | 82,640 (7.50%) | 231,247 (2.92%) |
| Occlusive arteriopathy of the lower limbs (AOMI) | No | 3,718,809 (97.18%) | 2,874,231 (96.29%) | 1,049,447 (95.20%) | 7,642,487 (96.57%) |
|  | Yes | 108,027 (2.82%) | 110,836 (3.71%) | 52,948 (4.80%) | 271,811 (3.43%) |
| Immunocompromised subjects including cancer patients, organ transplant, HIV patients, and chronic autoimmune or inflammatory diseases treated with immunosuppressive or biologic drugs | No | 3,190,210 (83.36%) | 2,360,202 (79.07%) | 898,453 (81.50%) | 6,448,865 (81.48%) |
|  | Yes | 636,626 (16.64%) | 624,865 (20.93%) | 203,942 (18.50%) | 1,465,433 (18.52%) |
| Cancer | No | 3,266,975 (85.37%) | 2,413,408 (80.85%) | 913,193 (82.84%) | 6,593,576 (83.31%) |
|  | Yes | 559,861 (14.63%) | 571,659 (19.15%) | 189,202 (17.16%) | 1,320,722 (16.69%) |
| Hematological tumours | No | 3,771,766 (98.56%) | 2,927,459 (98.07%) | 1,082,014 (98.15%) | 7,781,239 (98.32%) |
|  | Yes | 55,070 (1.44%) | 57,608 (1.93%) | 20,381 (1.85%) | 133,059 (1.68%) |
| Solid tumours | No | 3,311,678 (86.54%) | 2,457,967 (82.34%) | 929,713 (84.34%) | 6,699,358 (84.65%) |
|  | Yes | 515,158 (13.46%) | 527,100 (17.66%) | 172,682 (15.66%) | 1,214,940 (15.35%) |
| Solid organ transplant | No | 3,815,987 (99.72%) | 2,980,784 (99.86%) | 1,102,121 (99.98%) | 7,898,892 (99.81%) |
|  | Yes | 10,849 (0.28%) | 4,283 (0.14%) | 274 (0.02%) | 15,406 (0.19%) |
| Stem cells transplant | No | 3,821,943 (99.87%) | 2,984,571 (99.98%) | 1,102,375 (100.00%) | 7,908,889 (99.93%) |
|  | Yes | 4,893 (0.13%) | 496 (0.02%) | 20 (0.00%) | 5,409 (0.07%) |
| HIV patients | No | 3,815,814 (99.71%) | 2,981,289 (99.87%) | 1,101,859 (99.95%) | 7,898,962 (99.81%) |
|  | Yes | 11,022 (0.29%) | 3,778 (0.13%) | 536 (0.05%) | 15,336 (0.19%) |
| Patients affected by chronic autoimmune or inflammatory diseases treated by immunosuppressive or biologic drugs | No | 3,756,220 (98.15%) | 2,926,713 (98.05%) | 1,085,332 (98.45%) | 7,768,265 (98.15%) |
|  | Yes | 70,616 (1.85%) | 58,354 (1.95%) | 17,063 (1.55%) | 146,033 (1.85%) |
| Chronic liver disease - diseases of the liver or pancreas | No | 3,762,982 (98.33%) | 2,940,615 (98.51%) | 1,089,258 (98.81%) | 7,792,855 (98.47%) |
|  | Yes | 63,854 (1.67%) | 44,452 (1.49%) | 13,137 (1.19%) | 121,443 (1.53%) |
| Severe renal disease | No | 3,809,423 (99.54%) | 2,971,091 (99.53%) | 1,098,781 (99.67%) | 7,879,295 (99.56%) |
|  | Yes | 17,413 (0.46%) | 13,976 (0.47%) | 3,614 (0.33%) | 35,003 (0.44%) |
| Number of comorbidities of interest (major groups of comorbidities) | N | 3,826,836 | 2,985,067 | 1,102,395 | 7,914,298 |
|  | Mean (±SD) | 0.80 (±0.99) | 1.02 (±1.07) | 1.21 (±1.08) | 0.94 (±1.04) |
|  | Min; Max | 0.00; 8.00 | 0.00; 8.00 | 0.00; 8.00 | 0.00; 8.00 |
|  | Median | 1.00 | 1.00 | 1.00 | 1.00 |
|  | Q1; Q3 | 0.00; 1.00 | 0.00; 2.00 | 0.00; 2.00 | 0.00; 1.00 |
| Number of comorbidities of interest (major groups of comorbidities) - categorical | None | 1,873,536 (48.96%) | 1,139,262 (38.17%) | 322,355 (29.24%) | 3,335,153 (42.14%) |
|  | 1 | 1,201,846 (31.41%) | 1,031,344 (34.55%) | 406,274 (36.85%) | 2,639,464 (33.35%) |
|  | 2 or more | 751,454 (19.64%) | 814,461 (27.28%) | 373,766 (33.90%) | 1,939,681 (24.51%) |
| Number of comorbidities of interest (detailed) | N | 3,826,836 | 2,985,067 | 1,102,395 | 7,914,298 |
|  | Mean (±SD) | 0.75 (±1.00) | 0.93 (±1.08) | 1.05 (±1.10) | 0.86 (±1.05) |
|  | Min; Max | 0.00; 11.00 | 0.00; 11.00 | 0.00; 9.00 | 0.00; 11.00 |
|  | Median | 0.00 | 1.00 | 1.00 | 1.00 |
|  | Q1; Q3 | 0.00; 1.00 | 0.00; 1.00 | 0.00; 2.00 | 0.00; 1.00 |
| Number of comorbidities of interest (detailed) - categorical | None | 1,998,234 (52.22%) | 1,304,514 (43.70%) | 416,035 (37.74%) | 3,718,783 (46.99%) |
|  | 1 | 1,142,781 (29.86%) | 974,439 (32.64%) | 376,821 (34.18%) | 2,494,041 (31.51%) |
|  | 2 | 444,541 (11.62%) | 445,252 (14.92%) | 195,282 (17.71%) | 1,085,075 (13.71%) |
|  | 3 | 161,932 (4.23%) | 173,214 (5.80%) | 77,940 (7.07%) | 413,086 (5.22%) |
|  | 4 | 55,468 (1.45%) | 61,370 (2.06%) | 26,480 (2.40%) | 143,318 (1.81%) |
|  | 5 | 17,268 (0.45%) | 19,418 (0.65%) | 7,560 (0.69%) | 44,246 (0.56%) |
|  | 6 or more | 6,612 (0.17%) | 6,860 (0.23%) | 2,277 (0.21%) | 15,749 (0.20%) |
| Charlson comorbidity index (unadjusted for age) at index date | N | 3,826,836 | 2,985,067 | 1,102,395 | 7,914,298 |
|  | Mean (±SD) | 0.45 (±0.71) | 0.51 (±0.78) | 0.53 (±0.82) | 0.48 (±0.75) |
|  | Min; Max | 0.00; 10.00 | 0.00; 9.00 | 0.00; 8.00 | 0.00; 10.00 |
|  | Median | 0.00 | 0.00 | 0.00 | 0.00 |
|  | Q1; Q3 | 0.00; 1.00 | 0.00; 1.00 | 0.00; 1.00 | 0.00; 1.00 |
| Follow-up duration (in days) | N | 3,826,836 | 2,985,067 | 1,102,395 | 7,914,298 |
|  | Mean (±SD) | 230.66 (±21.92) | 231.02 (±24.96) | 226.46 (±38.99) | 230.21 (±26.10) |
|  | Min; Max | 1.00; 302.00 | 1.00; 302.00 | 1.00; 292.00 | 1.00; 302.00 |
|  | Median | 234.00 | 235.00 | 235.00 | 234.00 |
|  | Q1; Q3 | 218.00; 249.00 | 220.00; 249.00 | 220.00; 249.00 | 219.00; 249.00 |
| Reason for the end of follow up | Admission into a medico-social housing establishment ESMS (other than EHPAD) | 156 (0.00%) | 362 (0.01%) | 537 (0.05%) | 1,055 (0.01%) |
|  | Admission into an EHPAD nursing home | 1,833 (0.05%) | 9,031 (0.30%) | 20,182 (1.83%) | 31,046 (0.39%) |
|  | Death | 24,530 (0.64%) | 42,399 (1.42%) | 55,045 (4.99%) | 121,974 (1.54%) |
|  | End of follow-up | 3,800,317 (99.31%) | 2,933,275 (98.26%) | 1,026,631 (93.13%) | 7,760,223 (98.05%) |
| Death during follow-up | No | 3,802,312 (99.36%) | 2,942,686 (98.58%) | 1,047,384 (95.01%) | 7,792,382 (98.46%) |
|  | Yes | 24,524 (0.64%) | 42,381 (1.42%) | 55,011 (4.99%) | 121,916 (1.54%) |
| In-hospital death during study period | No | 3,807,072 (99.48%) | 2,950,340 (98.84%) | 1,062,654 (96.40%) | 7,820,066 (98.81%) |
|  | Yes | 19,764 (0.52%) | 34,727 (1.16%) | 39,741 (3.60%) | 94,232 (1.19%) |
| Field in which the death occured (MCO, HAD, SSR or PSY) | HAD | 1,798 (0.05%) | 3,014 (0.10%) | 3,886 (0.35%) | 8,698 (0.11%) |
|  | MCO | 17,078 (0.45%) | 29,365 (0.98%) | 32,003 (2.90%) | 78,446 (0.99%) |
|  | No death | 3,807,072 (99.48%) | 2,950,340 (98.84%) | 1,062,654 (96.40%) | 7,820,066 (98.81%) |
|  | RIP | 24 (0.00%) | 24 (0.00%) | 14 (0.00%) | 62 (0.00%) |
|  | SSR | 864 (0.02%) | 2,324 (0.08%) | 3,838 (0.35%) | 7,026 (0.09%) |
| Last DP/DR of RUM is a P/I code | No | 3,826,782 (100.00%) | 2,984,983 (100.00%) | 1,102,266 (99.99%) | 7,914,031 (100.00%) |
|  | Yes | 54 (0.00%) | 84 (0.00%) | 129 (0.01%) | 267 (0.00%) |
| Last DP/DR of RUM is a cardiovascular code | No | 3,826,836 (100.00%) | 2,985,067 (100.00%) | 1,102,395 (100.00%) | 7,914,298 (100.00%) |
| Last DP/DR of RUM is a respiratory code | No | 3,826,835 (100.00%) | 2,985,066 (100.00%) | 1,102,394 (100.00%) | 7,914,295 (100.00%) |
|  | Yes | 1 (0.00%) | 1 (0.00%) | 1 (0.00%) | 3 (0.00%) |
| Last DP of RUM associated with a hospital death | I5009 | 70 (0.00%) | 386 (0.01%) | 1,102 (0.10%) | 1,558 (0.02%) |
|  | I5019 | 45 (0.00%) | 267 (0.01%) | 649 (0.06%) | 961 (0.01%) |
|  | J181 | 94 (0.00%) | 231 (0.01%) | 421 (0.04%) | 746 (0.01%) |
|  | J189 | 99 (0.00%) | 314 (0.01%) | 479 (0.04%) | 892 (0.01%) |
|  | J690 | 213 (0.01%) | 619 (0.02%) | 929 (0.08%) | 1,761 (0.02%) |
|  | J9600 | 292 (0.01%) | 477 (0.02%) | 434 (0.04%) | 1,203 (0.02%) |
|  | Other | 3,821,614 (99.86%) | 2,976,345 (99.71%) | 1,092,849 (99.13%) | 7,890,808 (99.70%) |
|  | R53+0 | 243 (0.01%) | 358 (0.01%) | 454 (0.04%) | 1,055 (0.01%) |
|  | R572 | 364 (0.01%) | 516 (0.02%) | 362 (0.03%) | 1,242 (0.02%) |
|  | U0710 | 228 (0.01%) | 618 (0.02%) | 1,034 (0.09%) | 1,880 (0.02%) |
|  | Z515 | 3,574 (0.09%) | 4,936 (0.17%) | 3,682 (0.33%) | 12,192 (0.15%) |
| Last DR of RUM associated with a hospital death | C189 | 38 (0.00%) | 40 (0.00%) | 53 (0.00%) | 131 (0.00%) |
|  | C20 | 77 (0.00%) | 101 (0.00%) | 40 (0.00%) | 218 (0.00%) |
|  | C220 | 150 (0.00%) | 163 (0.01%) | 43 (0.00%) | 356 (0.00%) |
|  | C221 | 60 (0.00%) | 66 (0.00%) | 29 (0.00%) | 155 (0.00%) |
|  | C250 | 118 (0.00%) | 144 (0.00%) | 88 (0.01%) | 350 (0.00%) |
|  | C259 | 85 (0.00%) | 97 (0.00%) | 34 (0.00%) | 216 (0.00%) |
|  | C341 | 208 (0.01%) | 160 (0.01%) | 53 (0.00%) | 421 (0.01%) |
|  | C343 | 97 (0.00%) | 83 (0.00%) | 24 (0.00%) | 204 (0.00%) |
|  | C349 | 360 (0.01%) | 274 (0.01%) | 108 (0.01%) | 742 (0.01%) |
|  | C509 | 114 (0.00%) | 159 (0.01%) | 95 (0.01%) | 368 (0.00%) |
|  | C56 | 89 (0.00%) | 137 (0.00%) | 42 (0.00%) | 268 (0.00%) |
|  | C61 | 114 (0.00%) | 254 (0.01%) | 156 (0.01%) | 524 (0.01%) |
|  | C64 | 77 (0.00%) | 85 (0.00%) | 45 (0.00%) | 207 (0.00%) |
|  | C679 | 82 (0.00%) | 128 (0.00%) | 78 (0.01%) | 288 (0.00%) |
|  | C920 | 34 (0.00%) | 106 (0.00%) | 38 (0.00%) | 178 (0.00%) |
|  | N185 | 21 (0.00%) | 82 (0.00%) | 85 (0.01%) | 188 (0.00%) |
|  | Other | 3,825,112 (99.95%) | 2,982,988 (99.93%) | 1,101,384 (99.91%) | 7,909,484 (99.94%) |
| Type of vaccine at index date | High-dose QIV | 386,268 (10.09%) | 406,238 (13.61%) | 183,705 (16.66%) | 976,211 (12.33%) |
|  | Standard-dose QIV | 3,440,568 (89.91%) | 2,578,829 (86.39%) | 918,690 (83.34%) | 6,938,087 (87.67%) |
| Vaccine brand at index date | Efluelda | 386,268 (10.09%) | 406,238 (13.61%) | 183,705 (16.66%) | 976,211 (12.33%) |
|  | Fluarix Tetra | 177,741 (4.64%) | 132,910 (4.45%) | 47,963 (4.35%) | 358,614 (4.53%) |
|  | Influvac Tetra | 1,779,849 (46.51%) | 1,360,704 (45.58%) | 487,482 (44.22%) | 3,628,035 (45.84%) |
|  | Vaxigriptetra | 1,482,978 (38.75%) | 1,085,215 (36.35%) | 383,245 (34.76%) | 2,951,438 (37.29%) |
| Vaccine administration (Pharmacist vs. Other) | other | 1,565,344 (40.90%) | 1,486,142 (49.79%) | 738,529 (66.99%) | 3,790,015 (47.89%) |
|  | pharmacist | 2,261,492 (59.10%) | 1,498,925 (50.21%) | 363,866 (33.01%) | 4,124,283 (52.11%) |
| Week of vaccination | Week 1: 35 | 7 (0.00%) | 2 (0.00%) | 0 (0.00%) | 9 (0.00%) |
|  | Week 2: 36 | 27 (0.00%) | 13 (0.00%) | 0 (0.00%) | 40 (0.00%) |
|  | Week 3: 37 | 101 (0.00%) | 42 (0.00%) | 9 (0.00%) | 152 (0.00%) |
|  | Week 4: 38 | 145 (0.00%) | 86 (0.00%) | 31 (0.00%) | 262 (0.00%) |
|  | Week 5: 39 | 579 (0.02%) | 448 (0.02%) | 170 (0.02%) | 1,197 (0.02%) |
|  | Week 6: 40 | 1,964 (0.05%) | 1,774 (0.06%) | 753 (0.07%) | 4,491 (0.06%) |
|  | Week 7: 41 | 3,997 (0.10%) | 3,498 (0.12%) | 1,560 (0.14%) | 9,055 (0.11%) |
|  | Week 8: 42 | 779,550 (20.37%) | 616,350 (20.65%) | 242,146 (21.97%) | 1,638,046 (20.70%) |
|  | Week 9: 43 | 468,085 (12.23%) | 403,507 (13.52%) | 172,755 (15.67%) | 1,044,347 (13.20%) |
|  | Week 10: 44 | 385,545 (10.07%) | 335,230 (11.23%) | 138,708 (12.58%) | 859,483 (10.86%) |
|  | Week 11: 45 | 445,136 (11.63%) | 380,707 (12.75%) | 150,519 (13.65%) | 976,362 (12.34%) |
|  | Week 12: 46 | 455,470 (11.90%) | 380,558 (12.75%) | 140,228 (12.72%) | 976,256 (12.34%) |
|  | Week 13: 47 | 405,582 (10.60%) | 307,452 (10.30%) | 97,399 (8.84%) | 810,433 (10.24%) |
|  | Week 14: 48 | 391,855 (10.24%) | 261,006 (8.74%) | 73,921 (6.71%) | 726,782 (9.18%) |
|  | Week 15: 49 | 229,768 (6.00%) | 138,337 (4.63%) | 38,531 (3.50%) | 406,636 (5.14%) |
|  | Week 16: 50 | 115,563 (3.02%) | 65,456 (2.19%) | 18,070 (1.64%) | 199,089 (2.52%) |
|  | Week 17: 51 | 52,173 (1.36%) | 30,616 (1.03%) | 8,991 (0.82%) | 91,780 (1.16%) |
|  | Week 18: 52 | 36,590 (0.96%) | 21,403 (0.72%) | 6,142 (0.56%) | 64,135 (0.81%) |
|  | Week 19: 1 | 25,411 (0.66%) | 17,823 (0.60%) | 5,384 (0.49%) | 48,618 (0.61%) |
|  | Week 20: 2 | 12,623 (0.33%) | 9,058 (0.30%) | 3,046 (0.28%) | 24,727 (0.31%) |
|  | Week 21: 3 | 6,968 (0.18%) | 4,914 (0.16%) | 1,681 (0.15%) | 13,563 (0.17%) |
|  | Week 22: 4 | 5,104 (0.13%) | 3,627 (0.12%) | 1,266 (0.11%) | 9,997 (0.13%) |
|  | Week 23: 5 | 2,680 (0.07%) | 1,820 (0.06%) | 590 (0.05%) | 5,090 (0.06%) |
|  | Week 24: 6 | 721 (0.02%) | 517 (0.02%) | 169 (0.02%) | 1,407 (0.02%) |
|  | Week 25: 7 | 449 (0.01%) | 291 (0.01%) | 106 (0.01%) | 846 (0.01%) |
|  | Week 26: 8 | 301 (0.01%) | 211 (0.01%) | 72 (0.01%) | 584 (0.01%) |
|  | Week 27: 9 | 212 (0.01%) | 172 (0.01%) | 94 (0.01%) | 478 (0.01%) |
|  | Week 28: 10 | 76 (0.00%) | 43 (0.00%) | 8 (0.00%) | 127 (0.00%) |
|  | Week 29: 11 | 52 (0.00%) | 40 (0.00%) | 10 (0.00%) | 102 (0.00%) |
|  | Week 30: 12 | 56 (0.00%) | 29 (0.00%) | 17 (0.00%) | 102 (0.00%) |
|  | Week 31: 13 | 46 (0.00%) | 37 (0.00%) | 19 (0.00%) | 102 (0.00%) |
| Second brand of influenza vaccine received in the season | Missing | 3,826,790 | 2,985,035 | 1,102,375 | 7,914,200 |
|  | Influvac Tetra | 5 (10.87%) | 3 (9.38%) | 2 (10.00%) | 10 (10.20%) |
|  | Vaxigriptetra | 41 (89.13%) | 29 (90.63%) | 18 (90.00%) | 88 (89.80%) |
| Number of seasons with influenza vaccination in the 5 preceding seasons | N | 3,826,836 | 2,985,067 | 1,102,395 | 7,914,298 |
|  | Mean (±SD) | 2.72 (±1.36) | 3.46 (±0.99) | 3.62 (±0.86) | 3.13 (±1.23) |
|  | Min; Max | 0.00; 5.00 | 0.00; 5.00 | 0.00; 4.00 | 0.00; 5.00 |
|  | Median | 3.00 | 4.00 | 4.00 | 4.00 |
|  | Q1; Q3 | 2.00; 4.00 | 3.00; 4.00 | 4.00; 4.00 | 2.00; 4.00 |
| Number of seasons with influenza vaccination in the 5 preceding seasons - categorical | 0 | 354,668 (9.27%) | 70,475 (2.36%) | 21,364 (1.94%) | 446,507 (5.64%) |
|  | 1 | 435,939 (11.39%) | 117,947 (3.95%) | 29,214 (2.65%) | 583,100 (7.37%) |
|  | 2 | 778,481 (20.34%) | 320,240 (10.73%) | 65,509 (5.94%) | 1,164,230 (14.71%) |
|  | 3 | 600,417 (15.69%) | 334,699 (11.21%) | 115,194 (10.45%) | 1,050,310 (13.27%) |
|  | 4 | 1,657,328 (43.31%) | 2,141,705 (71.75%) | 871,114 (79.02%) | 4,670,147 (59.01%) |
|  | 5 | 3 (0.00%) | 1 (0.00%) | 0 (0.00%) | 4 (0.00%) |
| Type of influenza vaccination in the previous season | HD | 145,385 (3.80%) | 162,278 (5.44%) | 71,109 (6.45%) | 378,772 (4.79%) |
|  | Not vaccinated | 561,309 (14.67%) | 206,282 (6.91%) | 72,365 (6.56%) | 839,956 (10.61%) |
|  | SD | 3,119,861 (81.53%) | 2,616,124 (87.64%) | 958,698 (86.97%) | 6,694,683 (84.59%) |
|  | both | 281 (0.01%) | 383 (0.01%) | 223 (0.02%) | 887 (0.01%) |
| Number of influenza vaccination during the five previous seasons (dispensation) | N | 3,826,836 | 2,985,067 | 1,102,395 | 7,914,298 |
|  | Mean (±SD) | 2.73 (±1.37) | 3.47 (±1.00) | 3.63 (±0.87) | 3.14 (±1.24) |
|  | Min; Max | 0.00; 16.00 | 0.00; 14.00 | 0.00; 11.00 | 0.00; 16.00 |
|  | Median | 3.00 | 4.00 | 4.00 | 4.00 |
|  | Q1; Q3 | 2.00; 4.00 | 3.00; 4.00 | 4.00; 4.00 | 2.00; 4.00 |
| Number of influenza vaccinations during the five previous seasons (dispensation) - categorical | 0 | 354,678 (9.27%) | 70,478 (2.36%) | 21,365 (1.94%) | 446,521 (5.64%) |
|  | 1 | 434,554 (11.36%) | 117,476 (3.94%) | 29,110 (2.64%) | 581,140 (7.34%) |
|  | 2 | 774,300 (20.23%) | 318,291 (10.66%) | 65,107 (5.91%) | 1,157,698 (14.63%) |
|  | 3 | 599,500 (15.67%) | 333,635 (11.18%) | 114,368 (10.37%) | 1,047,503 (13.24%) |
|  | 4 | 1,643,129 (42.94%) | 2,120,868 (71.05%) | 862,917 (78.28%) | 4,626,914 (58.46%) |
|  | 5 | 19,733 (0.52%) | 23,749 (0.80%) | 9,306 (0.84%) | 52,788 (0.67%) |
|  | 6 or more | 942 (0.02%) | 570 (0.02%) | 222 (0.02%) | 1,734 (0.02%) |
| Vaccination status for COVID-19 at index date | On-going | 3,735 (0.10%) | 4,124 (0.14%) | 3,111 (0.28%) | 10,970 (0.14%) |
|  | Primo-vaccination | 3,743,605 (97.83%) | 2,897,866 (97.08%) | 1,051,739 (95.40%) | 7,693,210 (97.21%) |
|  | Unvaccinated | 79,496 (2.08%) | 83,077 (2.78%) | 47,545 (4.31%) | 210,118 (2.65%) |
| Pneumococcal vaccination status (presence during the 5 years preceeding index date) | No | 3,401,847 (88.89%) | 2,616,114 (87.64%) | 985,354 (89.38%) | 7,003,315 (88.49%) |
|  | Yes | 424,989 (11.11%) | 368,953 (12.36%) | 117,041 (10.62%) | 910,983 (11.51%) |
| Urinary tract infection-related hospitalizations | No | 3,813,829 (99.66%) | 2,963,648 (99.28%) | 1,083,727 (98.31%) | 7,861,204 (99.33%) |
|  | Yes | 13,007 (0.34%) | 21,419 (0.72%) | 18,668 (1.69%) | 53,094 (0.67%) |
| Number of Urinary tract infection-related hospitalizations | N | 3,826,836 | 2,985,067 | 1,102,395 | 7,914,298 |
|  | Mean (±SD) | 0.00 (±0.07) | 0.01 (±0.10) | 0.02 (±0.15) | 0.01 (±0.10) |
|  | Min; Max | 0.00; 9.00 | 0.00; 16.00 | 0.00; 8.00 | 0.00; 16.00 |
|  | Median | 0.00 | 0.00 | 0.00 | 0.00 |
|  | Q1; Q3 | 0.00; 0.00 | 0.00; 0.00 | 0.00; 0.00 | 0.00; 0.00 |
| **Number of Urinary tract infection-related hospitalizations in patients with at least one UTI hospitalization** | **Missing** | **3,813,829** | **2,963,648** | **1,083,727** | **7,861,204** |
|  | N | 13,007 | 21,419 | 18,668 | 53,094 |
|  | Mean (±SD) | 1.13 (±0.43) | 1.13 (±0.43) | 1.11 (±0.35) | 1.12 (±0.41) |
|  | Min; Max | 1.00; 9.00 | 1.00; 16.00 | 1.00; 8.00 | 1.00; 16.00 |
|  | Median | 1.00 | 1.00 | 1.00 | 1.00 |
|  | Q1; Q3 | 1.00; 1.00 | 1.00; 1.00 | 1.00; 1.00 | 1.00; 1.00 |
| Erysipelas-related hospitalizations | No | 3,824,105 (99.93%) | 2,980,864 (99.86%) | 1,098,051 (99.61%) | 7,903,020 (99.86%) |
|  | Yes | 2,731 (0.07%) | 4,203 (0.14%) | 4,344 (0.39%) | 11,278 (0.14%) |
| Number of Erysipelas-related hospitalizations | N | 3,826,836 | 2,985,067 | 1,102,395 | 7,914,298 |
|  | Mean (±SD) | 0.00 (±0.03) | 0.00 (±0.05) | 0.00 (±0.07) | 0.00 (±0.05) |
|  | Min; Max | 0.00; 9.00 | 0.00; 8.00 | 0.00; 5.00 | 0.00; 9.00 |
|  | Median | 0.00 | 0.00 | 0.00 | 0.00 |
|  | Q1; Q3 | 0.00; 0.00 | 0.00; 0.00 | 0.00; 0.00 | 0.00; 0.00 |
| **Number of Erysipelas-related hospitalizations in patients with at least one Erysipelas hospitalization** | **Missing** | **3,824,105** | **2,980,864** | **1,098,051** | **7,903,020** |
|  | N | 2,731 | 4,203 | 4,344 | 11,278 |
|  | Mean (±SD) | 1.15 (±0.46) | 1.15 (±0.43) | 1.13 (±0.39) | 1.14 (±0.42) |
|  | Min; Max | 1.00; 9.00 | 1.00; 8.00 | 1.00; 5.00 | 1.00; 9.00 |
|  | Median | 1.00 | 1.00 | 1.00 | 1.00 |
|  | Q1; Q3 | 1.00; 1.00 | 1.00; 1.00 | 1.00; 1.00 | 1.00; 1.00 |
| Cataract-related hospitalizations | No | 3,723,282 (97.29%) | 2,860,824 (95.84%) | 1,081,403 (98.10%) | 7,665,509 (96.86%) |
|  | Yes | 103,554 (2.71%) | 124,243 (4.16%) | 20,992 (1.90%) | 248,789 (3.14%) |
| Number of Cataract-related hospitalizations | N | 3,826,836 | 2,985,067 | 1,102,395 | 7,914,298 |
|  | Mean (±SD) | 0.04 (±0.26) | 0.06 (±0.32) | 0.03 (±0.22) | 0.05 (±0.28) |
|  | Min; Max | 0.00; 5.00 | 0.00; 3.00 | 0.00; 3.00 | 0.00; 5.00 |
|  | Median | 0.00 | 0.00 | 0.00 | 0.00 |
|  | Q1; Q3 | 0.00; 0.00 | 0.00; 0.00 | 0.00; 0.00 | 0.00; 0.00 |
| **Number of Cataract-related hospitalizations in patients with at least one Cataract hospitalization** | **Missing** | **3,723,282** | **2,860,824** | **1,081,403** | **7,665,509** |
|  | N | 103,554 | 124,243 | 20,992 | 248,789 |
|  | Mean (±SD) | 1.50 (±0.50) | 1.53 (±0.50) | 1.50 (±0.50) | 1.51 (±0.50) |
|  | Min; Max | 1.00; 5.00 | 1.00; 3.00 | 1.00; 3.00 | 1.00; 5.00 |
|  | Median | 1.00 | 2.00 | 1.50 | 2.00 |
|  | Q1; Q3 | 1.00; 2.00 | 1.00; 2.00 | 1.00; 2.00 | 1.00; 2.00 |
| Presence of influenza hospitalizations | No | 3,824,233 (99.93%) | 2,980,888 (99.86%) | 1,098,713 (99.67%) | 7,903,834 (99.87%) |
|  | Yes | 2,603 (0.07%) | 4,179 (0.14%) | 3,682 (0.33%) | 10,464 (0.13%) |
| Number of influenza hospitalizations | N | 3,826,836 | 2,985,067 | 1,102,395 | 7,914,298 |
|  | Mean (±SD) | 0.00 (±0.03) | 0.00 (±0.04) | 0.00 (±0.07) | 0.00 (±0.04) |
|  | Min; Max | 0.00; 3.00 | 0.00; 3.00 | 0.00; 4.00 | 0.00; 4.00 |
|  | Median | 0.00 | 0.00 | 0.00 | 0.00 |
|  | Q1; Q3 | 0.00; 0.00 | 0.00; 0.00 | 0.00; 0.00 | 0.00; 0.00 |
| **Number of influenza hospitalizations in patients with at least one influenza hospitalization** | **Missing** | **3,824,233** | **2,980,888** | **1,098,713** | **7,903,834** |
|  | N | 2,603 | 4,179 | 3,682 | 10,464 |
|  | Mean (±SD) | 1.09 (±0.30) | 1.09 (±0.30) | 1.09 (±0.30) | 1.09 (±0.30) |
|  | Min; Max | 1.00; 3.00 | 1.00; 3.00 | 1.00; 4.00 | 1.00; 4.00 |
|  | Median | 1.00 | 1.00 | 1.00 | 1.00 |
|  | Q1; Q3 | 1.00; 1.00 | 1.00; 1.00 | 1.00; 1.00 | 1.00; 1.00 |
| Presence of pneumonia hospitalizations | No | 3,806,412 (99.47%) | 2,952,019 (98.89%) | 1,070,464 (97.10%) | 7,828,895 (98.92%) |
|  | Yes | 20,424 (0.53%) | 33,048 (1.11%) | 31,931 (2.90%) | 85,403 (1.08%) |
| Number of pneumonia hospitalizations | N | 3,826,836 | 2,985,067 | 1,102,395 | 7,914,298 |
|  | Mean (±SD) | 0.01 (±0.09) | 0.01 (±0.13) | 0.03 (±0.20) | 0.01 (±0.12) |
|  | Min; Max | 0.00; 7.00 | 0.00; 9.00 | 0.00; 6.00 | 0.00; 9.00 |
|  | Median | 0.00 | 0.00 | 0.00 | 0.00 |
|  | Q1; Q3 | 0.00; 0.00 | 0.00; 0.00 | 0.00; 0.00 | 0.00; 0.00 |
| **Number of pneumonia hospitalizations in patients with at least one pneumonia hospitalization** | **Missing** | **3,806,412** | **2,952,019** | **1,070,464** | **7,828,895** |
|  | N | 20,424 | 33,048 | 31,931 | 85,403 |
|  | Mean (±SD) | 1.14 (±0.41) | 1.14 (±0.41) | 1.13 (±0.39) | 1.14 (±0.40) |
|  | Min; Max | 1.00; 7.00 | 1.00; 9.00 | 1.00; 6.00 | 1.00; 9.00 |
|  | Median | 1.00 | 1.00 | 1.00 | 1.00 |
|  | Q1; Q3 | 1.00; 1.00 | 1.00; 1.00 | 1.00; 1.00 | 1.00; 1.00 |
| Presence of P/I hospitalizations | No | 3,804,454 (99.42%) | 2,948,827 (98.79%) | 1,067,637 (96.85%) | 7,820,918 (98.82%) |
|  | Yes | 22,382 (0.58%) | 36,240 (1.21%) | 34,758 (3.15%) | 93,380 (1.18%) |
| Number of P/I hospitalizations | N | 3,826,836 | 2,985,067 | 1,102,395 | 7,914,298 |
|  | Mean (±SD) | 0.01 (±0.10) | 0.01 (±0.14) | 0.04 (±0.22) | 0.01 (±0.13) |
|  | Min; Max | 0.00; 7.00 | 0.00; 9.00 | 0.00; 6.00 | 0.00; 9.00 |
|  | Median | 0.00 | 0.00 | 0.00 | 0.00 |
|  | Q1; Q3 | 0.00; 0.00 | 0.00; 0.00 | 0.00; 0.00 | 0.00; 0.00 |
| **Number of P/I hospitalizations in patients with at least one influenza or one pneumonia hospitalization** | **Missing** | **3,804,454** | **2,948,827** | **1,067,637** | **7,820,918** |
|  | N | 22,382 | 36,24 | 34,758 | 93,38 |
|  | Mean (±SD) | 1.17 (±0.46) | 1.17 (±0.45) | 1.16 (±0.42) | 1.16 (±0.44) |
|  | Min; Max | 1.00; 7.00 | 1.00; 9.00 | 1.00; 6.00 | 1.00; 9.00 |
|  | Median | 1.00 | 1.00 | 1.00 | 1.00 |
|  | Q1; Q3 | 1.00; 1.00 | 1.00; 1.00 | 1.00; 1.00 | 1.00; 1.00 |
| Presence of respiratory disease hospitalizations | No | 3,790,865 (99.06%) | 2,932,369 (98.23%) | 1,056,371 (95.83%) | 7,779,605 (98.30%) |
|  | Yes | 35,971 (0.94%) | 52,698 (1.77%) | 46,024 (4.17%) | 134,693 (1.70%) |
| Number of respiratory disease hospitalizations | N | 3,826,836 | 2,985,067 | 1,102,395 | 7,914,298 |
|  | Mean (±SD) | 0.01 (±0.13) | 0.02 (±0.17) | 0.05 (±0.25) | 0.02 (±0.17) |
|  | Min; Max | 0.00; 9.00 | 0.00; 9.00 | 0.00; 7.00 | 0.00; 9.00 |
|  | Median | 0.00 | 0.00 | 0.00 | 0.00 |
|  | Q1; Q3 | 0.00; 0.00 | 0.00; 0.00 | 0.00; 0.00 | 0.00; 0.00 |
| **Number of respiratory hospitalizations in patients with at least one respiratory hospitalization** | **Missing** | **3,790,865** | **2,932,369** | **1,056,371** | **7,779,605** |
|  | N | 35,971 | 52,698 | 46,024 | 134,693 |
|  | Mean (±SD) | 1.21 (±0.55) | 1.20 (±0.53) | 1.17 (±0.45) | 1.19 (±0.51) |
|  | Min; Max | 1.00; 9.00 | 1.00; 9.00 | 1.00; 7.00 | 1.00; 9.00 |
|  | Median | 1.00 | 1.00 | 1.00 | 1.00 |
|  | Q1; Q3 | 1.00; 1.00 | 1.00; 1.00 | 1.00; 1.00 | 1.00; 1.00 |
| Presence of cardiovascular disease hospitalizations | No | 3,739,446 (97.72%) | 2,848,838 (95.44%) | 996,960 (90.44%) | 7,585,244 (95.84%) |
|  | Yes | 87,390 (2.28%) | 136,229 (4.56%) | 105,435 (9.56%) | 329,054 (4.16%) |
| Number of cardiovascular disease hospitalizations | N | 3,826,836 | 2,985,067 | 1,102,395 | 7,914,298 |
|  | Mean (±SD) | 0.03 (±0.22) | 0.06 (±0.31) | 0.13 (±0.44) | 0.05 (±0.30) |
|  | Min; Max | 0.00; 12.00 | 0.00; 12.00 | 0.00; 14.00 | 0.00; 14.00 |
|  | Median | 0.00 | 0.00 | 0.00 | 0.00 |
|  | Q1; Q3 | 0.00; 0.00 | 0.00; 0.00 | 0.00; 0.00 | 0.00; 0.00 |
| **Number of cardiovascular hospitalizations in patients with at least one cardiovascular hospitalization** | **Missing** | **3,739,446** | **2,848,838** | **996,96** | **7,585,244** |
|  | N | 87,39 | 136,229 | 105,435 | 329,054 |
|  | Mean (±SD) | 1.29 (±0.66) | 1.32 (±0.71) | 1.32 (±0.67) | 1.31 (±0.68) |
|  | Min; Max | 1.00; 12.00 | 1.00; 12.00 | 1.00; 14.00 | 1.00; 14.00 |
|  | Median | 1.00 | 1.00 | 1.00 | 1.00 |
|  | Q1; Q3 | 1.00; 1.00 | 1.00; 1.00 | 1.00; 1.00 | 1.00; 1.00 |
| Presence of cardio-respiratory disease hospitalizations | No | 3,729,572 (97.46%) | 2,835,404 (94.99%) | 986,426 (89.48%) | 7,551,402 (95.41%) |
|  | Yes | 97,264 (2.54%) | 149,663 (5.01%) | 115,969 (10.52%) | 362,896 (4.59%) |
| Number of cardio-respiratory disease hospitalizations | N | 3,826,836 | 2,985,067 | 1,102,395 | 7,914,298 |
|  | Mean (±SD) | 0.04 (±0.30) | 0.08 (±0.43) | 0.17 (±0.60) | 0.07 (±0.41) |
|  | Min; Max | 0.00; 18.00 | 0.00; 18.00 | 0.00; 18.00 | 0.00; 18.00 |
|  | Median | 0.00 | 0.00 | 0.00 | 0.00 |
|  | Q1; Q3 | 0.00; 0.00 | 0.00; 0.00 | 0.00; 0.00 | 0.00; 0.00 |
| **Number of cardio-respiratory hospitalizations in patients with at least one cardio-respiratory hospitalization** | **Missing** | **3,729,572** | **2,835,404** | **986,426** | **7,551,402** |
|  | N | 97,264 | 149,663 | 115,969 | 362,896 |
|  | Mean (±SD) | 1.61 (±1.04) | 1.63 (±1.05) | 1.66 (±1.00) | 1.63 (±1.03) |
|  | Min; Max | 1.00; 18.00 | 1.00; 18.00 | 1.00; 18.00 | 1.00; 18.00 |
|  | Median | 1.00 | 1.00 | 1.00 | 1.00 |
|  | Q1; Q3 | 1.00; 2.00 | 1.00; 2.00 | 1.00; 2.00 | 1.00; 2.00 |
| Long Term Disease ICD-10 and name | Other | 749,023 (34.08%) | 688,580 (30.79%) | 307,372 (30.42%) | 1,744,975 (32.05%) |
|  | E11 - Diabete sucre de type 2 | 572,620 (26.06%) | 455,650 (20.38%) | 132,073 (13.07%) | 1,160,343 (21.31%) |
|  | I25 - Cardiopathie ischemique chronique | 193,630 (8.81%) | 209,942 (9.39%) | 92,978 (9.20%) | 496,550 (9.12%) |
|  | I48 - Fibrillation et flutter auriculaires | 99,689 (4.54%) | 176,331 (7.89%) | 109,334 (10.82%) | 385,354 (7.08%) |
|  | C61 - Tumeur maligne de la prostate | 96,809 (4.41%) | 115,581 (5.17%) | 37,748 (3.74%) | 250,138 (4.59%) |
|  | C50 - Tumeur maligne du sein | 98,100 (4.46%) | 92,763 (4.15%) | 35,166 (3.48%) | 226,029 (4.15%) |
|  | I64 - Accident vasculaire cerebral, non precise comme etant hemorragique ou par infarctus | 40,948 (1.86%) | 56,422 (2.52%) | 35,057 (3.47%) | 132,427 (2.43%) |
|  | I70 - Atherosclerose | 42,453 (1.93%) | 44,351 (1.98%) | 22,029 (2.18%) | 108,833 (2.00%) |
|  | I21 - Infarctus aigu du myocarde | 50,806 (2.31%) | 40,367 (1.81%) | 15,748 (1.56%) | 106,921 (1.96%) |
|  | I50 - Insuffisance cardiaque | 25,539 (1.16%) | 39,573 (1.77%) | 38,069 (3.77%) | 103,181 (1.90%) |
|  | N18 - Maladie renale chronique | 26,417 (1.20%) | 38,219 (1.71%) | 23,304 (2.31%) | 87,940 (1.62%) |
|  | I702 - Atherosclerose des arteres distales | 34,683 (1.58%) | 33,238 (1.49%) | 15,224 (1.51%) | 83,145 (1.53%) |
|  | F00 - Demence de la maladie d'Alzheimer (G30.-) | 8,404 (0.38%) | 36,774 (1.64%) | 34,676 (3.43%) | 79,854 (1.47%) |
|  | C18 - Tumeur maligne du colon | 26,553 (1.21%) | 33,311 (1.49%) | 18,178 (1.80%) | 78,042 (1.43%) |
|  | I49 - Autres arythmies cardiaques | 18,501 (0.84%) | 33,061 (1.48%) | 22,147 (2.19%) | 73,709 (1.35%) |
|  | G20 - Maladie de Parkinson | 24,084 (1.10%) | 36,514 (1.63%) | 12,389 (1.23%) | 72,987 (1.34%) |
|  | E10 - Diabete sucre de type 1 | 36,698 (1.67%) | 26,972 (1.21%) | 8,848 (0.88%) | 72,518 (1.33%) |
|  | C67 - Tumeur maligne de la vessie | 22,451 (1.02%) | 28,989 (1.30%) | 12,228 (1.21%) | 63,668 (1.17%) |
|  | I35 - Atteintes non rhumatismales de la valvule aortique | 17,660 (0.80%) | 27,202 (1.22%) | 17,628 (1.74%) | 62,490 (1.15%) |
|  | I10 - Hypertension essentielle (primitive) | 12,495 (0.57%) | 22,238 (0.99%) | 20,380 (2.02%) | 55,113 (1.01%) |
