## Supplemental Table 4 for "Relative effectiveness of high-dose vs standard-dose influenza vaccines in preventing hospitalizations: a national retrospective cohort study in France, 2022/23 season"

| **Variables** | **Modality** | **HD Cohort** | | **SD Cohort** | |
| --- | --- | --- | --- | --- | --- |
|  |  | **Matched** | **Unmatched** | **Matched** | **Unmatched** |
| **N** | **Total** | **675,412** | **266,584** | **2,701,648** | **4,032,977** |
| Age at inclusion | N | 675,412 | 266,584 | 2,701,648 | 4,032,977 |
|  | Mean (±SD) | 76.83 (±7.68) | 80.11 (±7.72) | 76.78 (±7.73) | 75.27 (±7.40) |
|  | Min; Max | 65.00; 111.00 | 65.00; 109.00 | 65.00; 111.00 | 65.00; 110.00 |
|  | Median | 76.00 | 80.00 | 76.00 | 74.00 |
|  | Q1; Q3 | 71.00; 82.00 | 74.00; 86.00 | 71.00; 82.00 | 69.00; 80.00 |
| Age at index date (large categories) | <75 years | 302,005 (44.71%) | 70,411 (26.41%) | 1,208,020 (44.71%) | 2,133,221 (52.89%) |
|  | 75-85 years | 265,528 (39.31%) | 125,985 (47.26%) | 1,062,112 (39.31%) | 1,437,635 (35.65%) |
|  | >85 years | 107,879 (15.97%) | 70,188 (26.33%) | 431,516 (15.97%) | 462,121 (11.46%) |
| Age at index date (small categories) | <=70 years | 161,140 (23.86%) | 31,033 (11.64%) | 658,895 (24.39%) | 1,261,763 (31.29%) |
|  | 71-75 years | 175,595 (26.00%) | 53,018 (19.89%) | 688,442 (25.48%) | 1,086,443 (26.94%) |
|  | 76-80 years | 130,552 (19.33%) | 57,257 (21.48%) | 526,741 (19.50%) | 739,599 (18.34%) |
|  | 81-85 years | 100,246 (14.84%) | 55,088 (20.66%) | 396,054 (14.66%) | 483,051 (11.98%) |
|  | 86-90 years | 71,777 (10.63%) | 43,274 (16.23%) | 284,581 (10.53%) | 311,911 (7.73%) |
|  | 91-95 years | 29,645 (4.39%) | 21,098 (7.91%) | 120,636 (4.47%) | 122,956 (3.05%) |
|  | 96-100 years | 5,897 (0.87%) | 5,270 (1.98%) | 24,176 (0.89%) | 24,919 (0.62%) |
|  | >100 years | 560 (0.08%) | 546 (0.20%) | 2,123 (0.08%) | 2,335 (0.06%) |
| Gender | Men | 303,862 (44.99%) | 117,966 (44.25%) | 1,215,448 (44.99%) | 1,828,320 (45.33%) |
|  | Women | 371,550 (55.01%) | 148,618 (55.75%) | 1,486,200 (55.01%) | 2,204,657 (54.67%) |
| Region of residence at index date | Ile-de-France | 98,509 (14.59%) | 40,608 (15.23%) | 394,036 (14.59%) | 509,072 (12.62%) |
| - Corrected | Centre-Val de Loire | 33,462 (4.95%) | 16,784 (6.30%) | 133,848 (4.95%) | 163,632 (4.06%) |
|  | BFC | 31,554 (4.67%) | 4,239 (1.59%) | 126,216 (4.67%) | 202,214 (5.01%) |
|  | Normandie | 31,681 (4.69%) | 39,318 (14.75%) | 126,724 (4.69%) | 252,286 (6.26%) |
|  | Hauts-de-France | 60,650 (8.98%) | 37,039 (13.89%) | 242,600 (8.98%) | 330,050 (8.18%) |
|  | Grand Est | 50,948 (7.54%) | 55,346 (20.76%) | 203,792 (7.54%) | 340,197 (8.44%) |
|  | Pays de la Loire | 42,052 (6.23%) | 6,985 (2.62%) | 168,208 (6.23%) | 275,107 (6.82%) |
|  | Bretagne | 39,124 (5.79%) | 7,105 (2.67%) | 156,496 (5.79%) | 270,019 (6.70%) |
|  | Nouvelle Aquitaine | 81,769 (12.11%) | 17,902 (6.72%) | 327,076 (12.11%) | 429,040 (10.64%) |
|  | Occitanie | 69,443 (10.28%) | 10,385 (3.90%) | 277,772 (10.28%) | 405,500 (10.05%) |
|  | AURA | 86,390 (12.79%) | 21,139 (7.93%) | 345,560 (12.79%) | 453,525 (11.25%) |
|  | PACA & Corse | 49,830 (7.38%) | 9,734 (3.65%) | 199,320 (7.38%) | 402,335 (9.98%) |
| Presence of C2S - Solidarity health | No | 652,694 (96.64%) | 258,718 (97.05%) | 2,612,621 (96.70%) | 3,907,786 (96.90%) |
| insurance free of charge  (formerly CMU-C) or with financial participation (formerly ACS) | Yes | 22,718 (3.36%) | 7,866 (2.95%) | 89,027 (3.30%) | 125,191 (3.10%) |
| Solidarity health insurance free of charge | No | 652,694 | 258,718 | 2,612,621 | 3,907,786 |
| (formerly CMU-C) or with financial | Free | 5,869 (25.83%) | 1,847 (23.48%) | 23,353 (26.23%) | 33,789 (26.99%) |
| participation (formerly ACS) | Unknown | 810 (3.57%) | 289 (3.67%) | 3,556 (3.99%) | 5,056 (4.04%) |
|  | Participatory | 16,039 (70.60%) | 5,730 (72.85%) | 62,118 (69.77%) | 86,346 (68.97%) |
| French social deprivation index (FDep) | 1st quintile | 134,033 (19.84%) | 53,646 (20.12%) | 534,373 (19.78%) | 743,777 (18.44%) |
| (quintile) | 2nd quintile | 133,609 (19.78%) | 49,932 (18.73%) | 535,348 (19.82%) | 786,945 (19.51%) |
|  | 3rd quintile | 138,963 (20.57%) | 53,489 (20.06%) | 560,556 (20.75%) | 827,562 (20.52%) |
|  | 4th quintile | 140,353 (20.78%) | 50,095 (18.79%) | 558,286 (20.66%) | 867,303 (21.51%) |
|  | 5th quintile | 128,454 (19.02%) | 59,422 (22.29%) | 513,085 (18.99%) | 807,390 (20.02%) |
| Local Potential Accessibility | Missing | 2,178 | 270 | 10,522 | 24,107 |
| to GP (LPA) - Corrected | 1st quintile | 136,258 (20.24%) | 60,337 (22.66%) | 532,761 (19.80%) | 730,020 (18.21%) |
|  | 2nd quintile | 129,386 (19.22%) | 43,733 (16.42%) | 565,874 (21.03%) | 850,423 (21.21%) |
|  | 3rd quintile | 140,781 (20.91%) | 66,599 (25.01%) | 540,679 (20.09%) | 844,683 (21.07%) |
|  | 4th quintile | 140,774 (20.91%) | 49,277 (18.50%) | 549,137 (20.41%) | 830,338 (20.71%) |
|  | 5th quintile | 126,035 (18.72%) | 46,368 (17.41%) | 502,675 (18.68%) | 753,406 (18.79%) |
| Patient's social security regimen | CANSSM | 3,233 (0.48%) | 2,957 (1.11%) | 12,516 (0.46%) | 17,774 (0.44%) |
|  | CAVIMAC | 669 (0.10%) | 336 (0.13%) | 2,329 (0.09%) | 2,884 (0.07%) |
|  | CNMSS | 1,229 (0.18%) | 396 (0.15%) | 4,825 (0.18%) | 7,243 (0.18%) |
|  | CRPCEN | 1,204 (0.18%) | 394 (0.15%) | 4,833 (0.18%) | 7,866 (0.20%) |
|  | ENIM | 1,320 (0.20%) | 439 (0.16%) | 6,253 (0.23%) | 9,024 (0.22%) |
|  | MSA | 42,238 (6.25%) | 15,152 (5.68%) | 178,765 (6.62%) | 280,958 (6.97%) |
|  | Other | 51 (0.01%) | 22 (0.01%) | 203 (0.01%) | 392 (0.01%) |
|  | RATP | 1,093 (0.16%) | 401 (0.15%) | 4,745 (0.18%) | 6,388 (0.16%) |
|  | RG | 617,579 (91.44%) | 243,743 (91.43%) | 2,459,084 (91.02%) | 3,659,193 (90.73%) |
|  | RSI | 134 (0.02%) | 25 (0.01%) | 463 (0.02%) | 820 (0.02%) |
|  | SNCF | 6,662 (0.99%) | 2,719 (1.02%) | 27,632 (1.02%) | 40,435 (1.00%) |
| Patients with an LTD status | No | 334,299 (49.50%) | 117,389 (44.03%) | 1,373,549 (50.84%) | 2,148,035 (53.26%) |
|  | Yes | 341,113 (50.50%) | 149,195 (55.97%) | 1,328,099 (49.16%) | 1,884,942 (46.74%) |
| Number of LTDs | N | 675,412 | 266,584 | 2,701,648 | 4,032,977 |
|  | Mean (±SD) | 0.70 (±0.83) | 0.80 (±0.88) | 0.68 (±0.82) | 0.63 (±0.80) |
|  | Min; Max | 0.00; 8.00 | 0.00; 7.00 | 0.00; 8.00 | 0.00; 8.00 |
|  | Median | 1.00 | 1.00 | 0.00 | 0.00 |
|  | Q1; Q3 | 0.00; 1.00 | 0.00; 1.00 | 0.00; 1.00 | 0.00; 1.00 |
| Number of LTDs among patients | Missing | 334,299 | 117,389 | 1,373,549 | 2,148,035 |
| having at least 1 LTD | One LTD | 238,315 (69.86%) | 100,072 (67.07%) | 934,240 (70.34%) | 1,357,601 (72.02%) |
|  | Two or more LTDs | 102,798 (30.14%) | 49,123 (32.93%) | 393,859 (29.66%) | 527,341 (27.98%) |
| Number of all-cause hospitalization | N | 675,412 | 266,584 | 2,701,648 | 4,032,977 |
| in the past 12 months | Mean (±SD) | 0.14 (±0.99) | 0.18 (±1.33) | 0.13 (±0.99) | 0.10 (±0.82) |
| (proxy for health status) | Min; Max | 0.00; 62.00 | 0.00; 53.00 | 0.00; 70.00 | 0.00; 64.00 |
|  | Median | 0.00 | 0.00 | 0.00 | 0.00 |
|  | Q1; Q3 | 0.00; 0.00 | 0.00; 0.00 | 0.00; 0.00 | 0.00; 0.00 |
| Number of all-cause hospitalization | None | 625,673 (92.64%) | 243,168 (91.22%) | 2,511,427 (92.96%) | 3,806,676 (94.39%) |
| in the past 12 months | 1 | 36,808 (5.45%) | 17,054 (6.40%) | 141,190 (5.23%) | 172,991 (4.29%) |
| (proxy for health status) - categorical | 2 | 7,445 (1.10%) | 3,611 (1.35%) | 27,952 (1.03%) | 31,524 (0.78%) |
|  | 3 | 2,184 (0.32%) | 1,009 (0.38%) | 8,405 (0.31%) | 8,746 (0.22%) |
|  | 4 | 1,046 (0.15%) | 463 (0.17%) | 3,702 (0.14%) | 3,848 (0.10%) |
|  | 5 | 584 (0.09%) | 257 (0.10%) | 2,232 (0.08%) | 2,052 (0.05%) |
|  | 6 | 332 (0.05%) | 168 (0.06%) | 1,262 (0.05%) | 1,222 (0.03%) |
|  | 7 | 179 (0.03%) | 93 (0.03%) | 654 (0.02%) | 708 (0.02%) |
|  | 8 | 127 (0.02%) | 79 (0.03%) | 558 (0.02%) | 506 (0.01%) |
|  | 9 | 89 (0.01%) | 45 (0.02%) | 307 (0.01%) | 333 (0.01%) |
|  | 10 or more | 945 (0.14%) | 637 (0.24%) | 3,959 (0.15%) | 4,371 (0.11%) |
| GP visits in the past 12 months | N | 675,412 | 266,584 | 2,701,648 | 4,032,977 |
| (proxy for health status) | Mean (±SD) | 5.80 (±4.42) | 6.50 (±4.98) | 5.73 (±4.38) | 5.49 (±4.24) |
|  | Min; Max | 0.00; 135.00 | 0.00; 197.00 | 0.00; 339.00 | 0.00; 316.00 |
|  | Median | 5.00 | 5.00 | 5.00 | 5.00 |
|  | Q1; Q3 | 3.00; 7.00 | 4.00; 8.00 | 3.00; 7.00 | 3.00; 7.00 |
| GP visits in the past 12 months | None | 41,220 (6.10%) | 14,918 (5.60%) | 159,287 (5.90%) | 253,324 (6.28%) |
| (proxy for health status) - categorical | 1 | 31,098 (4.60%) | 9,127 (3.42%) | 125,082 (4.63%) | 218,720 (5.42%) |
|  | 2 | 53,846 (7.97%) | 16,629 (6.24%) | 220,751 (8.17%) | 365,199 (9.06%) |
|  | 3 | 70,036 (10.37%) | 23,019 (8.63%) | 286,878 (10.62%) | 461,499 (11.44%) |
|  | 4 | 101,414 (15.02%) | 37,728 (14.15%) | 415,107 (15.36%) | 613,232 (15.21%) |
|  | 5 | 89,254 (13.21%) | 35,004 (13.13%) | 362,655 (13.42%) | 525,423 (13.03%) |
|  | 6 | 70,593 (10.45%) | 28,281 (10.61%) | 282,710 (10.46%) | 407,735 (10.11%) |
|  | 7 | 52,422 (7.76%) | 21,822 (8.19%) | 209,431 (7.75%) | 299,368 (7.42%) |
|  | 8 | 38,864 (5.75%) | 16,379 (6.14%) | 151,282 (5.60%) | 216,240 (5.36%) |
|  | 9 | 28,138 (4.17%) | 12,353 (4.63%) | 109,713 (4.06%) | 157,342 (3.90%) |
|  | 10 or more | 98,527 (14.59%) | 51,324 (19.25%) | 378,752 (14.02%) | 514,895 (12.77%) |
| Diabetes | No | 533,427 (78.98%) | 203,117 (76.19%) | 2,143,632 (79.35%) | 3,271,198 (81.11%) |
|  | Yes | 141,985 (21.02%) | 63,467 (23.81%) | 558,016 (20.65%) | 761,779 (18.89%) |
| Severe obesity | No | 619,582 (91.73%) | 238,828 (89.59%) | 2,485,132 (91.99%) | 3,725,561 (92.38%) |
|  | Yes | 55,830 (8.27%) | 27,756 (10.41%) | 216,516 (8.01%) | 307,416 (7.62%) |
| Severe malnutrition | No | 634,786 (93.99%) | 242,626 (91.01%) | 2,544,043 (94.17%) | 3,836,076 (95.12%) |
|  | Yes | 40,626 (6.01%) | 23,958 (8.99%) | 157,605 (5.83%) | 196,901 (4.88%) |
| COPD and asthma | No | 593,174 (87.82%) | 229,711 (86.17%) | 2,380,875 (88.13%) | 3,586,785 (88.94%) |
|  | Yes | 82,238 (12.18%) | 36,873 (13.83%) | 320,773 (11.87%) | 446,192 (11.06%) |
| Dementia - neurological or | No | 657,373 (97.33%) | 253,775 (95.20%) | 2,633,849 (97.49%) | 3,951,192 (97.97%) |
| degenerative disease | Yes | 18,039 (2.67%) | 12,809 (4.80%) | 67,799 (2.51%) | 81,785 (2.03%) |
| Cardiovascular disease | No | 486,957 (72.10%) | 179,118 (67.19%) | 1,968,696 (72.87%) | 3,021,809 (74.93%) |
|  | Yes | 188,455 (27.90%) | 87,466 (32.81%) | 732,952 (27.13%) | 1,011,168 (25.07%) |
| Myocardial infarction | No | 673,323 (99.69%) | 265,649 (99.65%) | 2,693,467 (99.70%) | 4,021,669 (99.72%) |
|  | Yes | 2,089 (0.31%) | 935 (0.35%) | 8,181 (0.30%) | 11,308 (0.28%) |
| Chronic coronary disease | No | 599,307 (88.73%) | 231,923 (87.00%) | 2,405,501 (89.04%) | 3,606,108 (89.42%) |
|  | Yes | 76,105 (11.27%) | 34,661 (13.00%) | 296,147 (10.96%) | 426,869 (10.58%) |
| Chronic heart failure | No | 653,396 (96.74%) | 254,606 (95.51%) | 2,617,329 (96.88%) | 3,925,912 (97.35%) |
|  | Yes | 22,016 (3.26%) | 11,978 (4.49%) | 84,319 (3.12%) | 107,065 (2.65%) |
| Occlusive arteriopathy of the lower limbs | No | 650,881 (96.37%) | 255,331 (95.78%) | 2,606,518 (96.48%) | 3,900,902 (96.73%) |
|  | Yes | 24,531 (3.63%) | 11,253 (4.22%) | 95,130 (3.52%) | 132,075 (3.27%) |
| Immunocompromised subjects including | No | 546,894 (80.97%) | 211,942 (79.50%) | 2,202,541 (81.53%) | 3,294,126 (81.68%) |
| cancer patients, organ transplant, HIV patients, and chronic autoimmune or inflammatory diseases treated with immunosuppressive or biologic drugs | Yes | 128,518 (19.03%) | 54,642 (20.50%) | 499,107 (18.47%) | 738,851 (18.32%) |
| Cancer | No | 559,704 (82.87%) | 216,913 (81.37%) | 2,252,021 (83.36%) | 3,367,529 (83.50%) |
|  | Yes | 115,708 (17.13%) | 49,671 (18.63%) | 449,627 (16.64%) | 665,448 (16.50%) |
| Hematological tumours | No | 663,478 (98.23%) | 261,418 (98.06%) | 2,655,331 (98.29%) | 3,967,112 (98.37%) |
|  | Yes | 11,934 (1.77%) | 5,166 (1.94%) | 46,317 (1.71%) | 65,865 (1.63%) |
| Solid tumours | No | 569,194 (84.27%) | 221,006 (82.90%) | 2,288,610 (84.71%) | 3,420,223 (84.81%) |
|  | Yes | 106,218 (15.73%) | 45,578 (17.10%) | 413,038 (15.29%) | 612,754 (15.19%) |
| Solid organ transplant | No | 673,949 (99.78%) | 265,957 (99.76%) | 2,695,956 (99.79%) | 4,025,723 (99.82%) |
|  | Yes | 1,463 (0.22%) | 627 (0.24%) | 5,692 (0.21%) | 7,254 (0.18%) |
| Stem cells transplant | No | 674,943 (99.93%) | 266,430 (99.94%) | 2,699,810 (99.93%) | 4,030,205 (99.93%) |
|  | Yes | 469 (0.07%) | 154 (0.06%) | 1,838 (0.07%) | 2,772 (0.07%) |
| HIV patients | No | 674,050 (99.80%) | 266,137 (99.83%) | 2,696,293 (99.80%) | 4,025,529 (99.82%) |
|  | Yes | 1,362 (0.20%) | 447 (0.17%) | 5,355 (0.20%) | 7,448 (0.18%) |
| Patients affected by chronic autoimmune | No | 662,521 (98.09%) | 261,399 (98.06%) | 2,651,864 (98.16%) | 3,958,669 (98.16%) |
| or inflammatory diseases treated by immunosuppressive or biologic drugs | Yes | 12,891 (1.91%) | 5,185 (1.94%) | 49,784 (1.84%) | 74,308 (1.84%) |
| Chronic liver disease - | No | 664,729 (98.42%) | 262,096 (98.32%) | 2,659,871 (98.45%) | 3,972,055 (98.49%) |
| diseases of the liver or pancreas | Yes | 10,683 (1.58%) | 4,488 (1.68%) | 41,777 (1.55%) | 60,922 (1.51%) |
| Severe renal disease | No | 672,170 (99.52%) | 265,079 (99.44%) | 2,689,095 (99.54%) | 4,016,337 (99.59%) |
|  | Yes | 3,242 (0.48%) | 1,505 (0.56%) | 12,553 (0.46%) | 16,640 (0.41%) |
| Number of comorbidities of interest | N | 675,412 | 266,584 | 2,701,648 | 4,032,977 |
| (major groups of comorbidities) | Mean (±SD) | 0.99 (±1.06) | 1.17 (±1.14) | 0.97 (±1.06) | 0.90 (±1.02) |
|  | Min; Max | 0.00; 8.00 | 0.00; 8.00 | 0.00; 8.00 | 0.00; 8.00 |
|  | Median | 1.00 | 1.00 | 1.00 | 1.00 |
|  | Q1; Q3 | 0.00; 2.00 | 0.00; 2.00 | 0.00; 2.00 | 0.00; 1.00 |
| Number of comorbidities of interest | None | 268,516 (39.76%) | 87,799 (32.93%) | 1,113,264 (41.21%) | 1,763,260 (43.72%) |
| (major groups of comorbidities) | 1 | 229,462 (33.97%) | 92,153 (34.57%) | 900,342 (33.33%) | 1,338,589 (33.19%) |
| - categorical | 2 or more | 177,434 (26.27%) | 86,632 (32.50%) | 688,042 (25.47%) | 931,128 (23.09%) |
| Number of comorbidities of interest | N | 675,412 | 266,584 | 2,701,648 | 4,032,977 |
| (detailed) | Mean (±SD) | 0.91 (±1.07) | 1.08 (±1.16) | 0.88 (±1.07) | 0.82 (±1.03) |
|  | Min; Max | 0.00; 9.00 | 0.00; 10.00 | 0.00; 11.00 | 0.00; 11.00 |
|  | Median | 1.00 | 1.00 | 1.00 | 1.00 |
|  | Q1; Q3 | 0.00; 1.00 | 0.00; 2.00 | 0.00; 1.00 | 0.00; 1.00 |
| Number of comorbidities of interest | None | 302,797 (44.83%) | 101,995 (38.26%) | 1,250,066 (46.27%) | 1,950,114 (48.35%) |
| (detailed) - categorical | 1 | 217,462 (32.20%) | 88,611 (33.24%) | 849,080 (31.43%) | 1,264,205 (31.35%) |
|  | 2 | 97,773 (14.48%) | 45,610 (17.11%) | 379,766 (14.06%) | 529,994 (13.14%) |
|  | 3 | 38,282 (5.67%) | 19,274 (7.23%) | 148,363 (5.49%) | 195,398 (4.85%) |
|  | 4 | 13,458 (1.99%) | 7,566 (2.84%) | 52,144 (1.93%) | 66,268 (1.64%) |
|  | 5 | 4,200 (0.62%) | 2,532 (0.95%) | 16,419 (0.61%) | 19,928 (0.49%) |
|  | 6 or more | 1,440 (0.21%) | 996 (0.37%) | 5,810 (0.22%) | 7,070 (0.18%) |
| Charlson comorbidity index | N | 675,412 | 266,584 | 2,701,648 | 4,032,977 |
| (unadjusted for age) at index date | Mean (±SD) | 0.51 (±0.78) | 0.58 (±0.84) | 0.50 (±0.76) | 0.46 (±0.73) |
|  | Min; Max | 0.00; 10.00 | 0.00; 8.00 | 0.00; 9.00 | 0.00; 9.00 |
|  | Median | 0.00 | 0.00 | 0.00 | 0.00 |
|  | Q1; Q3 | 0.00; 1.00 | 0.00; 1.00 | 0.00; 1.00 | 0.00; 1.00 |
| Follow-up duration (in days) | N | 675,412 | 266,584 | 2,701,648 | 4,032,977 |
|  | Mean (±SD) | 236.87 (±26.02) | 243.57 (±28.13) | 237.02 (±25.18) | 223.63 (±24.69) |
|  | Min; Max | 1.00; 277.00 | 1.00; 301.00 | 1.00; 278.00 | 1.00; 302.00 |
|  | Median | 241.00 | 252.00 | 241.00 | 225.00 |
|  | Q1; Q3 | 229.00; 253.00 | 241.00; 255.00 | 229.00; 253.00 | 213.00; 239.00 |
| Reason for the end of follow up | Admission into a medico-social housing establishment (nursing home excluded) | 96 (0.01%) | 75 (0.03%) | 394 (0.01%) | 460 (0.01%) |
|  | Death | 3,247 (0.48%) | 2,010 (0.75%) | 12,308 (0.46%) | 12,800 (0.32%) |
|  | End of follow-up | 12,354 (1.83%) | 7,527 (2.82%) | 45,393 (1.68%) | 52,867 (1.31%) |
|  | Exit from nursing home | 659,715 (97.68%) | 256,972 (96.39%) | 2,643,553 (97.85%) | 3,966,850 (98.36%) |
| Death during follow-up | No | 663,063 (98.17%) | 259,061 (97.18%) | 2,656,275 (98.32%) | 3,980,137 (98.69%) |
|  | Yes | 12,349 (1.83%) | 7,523 (2.82%) | 45,373 (1.68%) | 52,840 (1.31%) |
| In-hospital death during study period | No | 666,061 (98.62%) | 261,040 (97.92%) | 2,666,564 (98.70%) | 3,991,398 (98.97%) |
|  | Yes | 9,351 (1.38%) | 5,544 (2.08%) | 35,084 (1.30%) | 41,579 (1.03%) |
| Field in which the death occurred | HAD | 902 (0.13%) | 576 (0.22%) | 3,237 (0.12%) | 3,665 (0.09%) |
|  | MCO | 7,763 (1.15%) | 4,488 (1.68%) | 29,206 (1.08%) | 34,806 (0.86%) |
|  | No death | 666,061 (98.62%) | 261,040 (97.92%) | 2,666,564 (98.70%) | 3,991,398 (98.97%) |
|  | RIP | 3 (0.00%) | 1 (0.00%) | 24 (0.00%) | 33 (0.00%) |
|  | SSR | 683 (0.10%) | 479 (0.18%) | 2,617 (0.10%) | 3,075 (0.08%) |
| Last DP/DR of RUM is a P/I code | No | 675,392 (100.00%) | 266,558 (99.99%) | 2,701,552 (100.00%) | 4,032,858 (100.00%) |
|  | Yes | 20 (0.00%) | 26 (0.01%) | 96 (0.00%) | 119 (0.00%) |
| Last DP/DR of RUM is a cardiovascular code | No | 675,412 (100.00%) | 266,584 (100.00%) | 2,701,648 (100.00%) | 4,032,977 (100.00%) |
| Last DP/DR of RUM is a respiratory code | No | 675,412 (100.00%) | 266,584 (100.00%) | 2,701,647 (100.00%) | 4,032,975 (100.00%) |
|  | Yes | 0 (0.00%) | 0 (0.00%) | 1 (0.00%) | 2 (0.00%) |
| Last DP of RUM associated | I5009 | 161 (0.02%) | 113 (0.04%) | 648 (0.02%) | 591 (0.01%) |
| with a hospital death | I5019 | 91 (0.01%) | 62 (0.02%) | 371 (0.01%) | 414 (0.01%) |
|  | J181 | 65 (0.01%) | 54 (0.02%) | 301 (0.01%) | 313 (0.01%) |
|  | J189 | 82 (0.01%) | 53 (0.02%) | 341 (0.01%) | 386 (0.01%) |
|  | J690 | 179 (0.03%) | 130 (0.05%) | 652 (0.02%) | 751 (0.02%) |
|  | J9600 | 141 (0.02%) | 81 (0.03%) | 434 (0.02%) | 513 (0.01%) |
|  | Other | 673,141 (99.66%) | 265,251 (99.50%) | 2,692,815 (99.67%) | 4,022,554 (99.74%) |
|  | R53+0 | 92 (0.01%) | 69 (0.03%) | 378 (0.01%) | 483 (0.01%) |
|  | R572 | 125 (0.02%) | 73 (0.03%) | 459 (0.02%) | 546 (0.01%) |
|  | U0710 | 182 (0.03%) | 114 (0.04%) | 761 (0.03%) | 777 (0.02%) |
|  | Z515 | 1,153 (0.17%) | 584 (0.22%) | 4,488 (0.17%) | 5,649 (0.14%) |
| Last DR of RUM associated | C189 | 17 (0.00%) | 9 (0.00%) | 43 (0.00%) | 59 (0.00%) |
| with a hospital death | C20 | 22 (0.00%) | 8 (0.00%) | 71 (0.00%) | 114 (0.00%) |
|  | C220 | 34 (0.01%) | 15 (0.01%) | 158 (0.01%) | 142 (0.00%) |
|  |  | 15 (0.00%) | 4 (0.00%) | 44 (0.00%) | 90 (0.00%) |
|  | C250 | 22 (0.00%) | 12 (0.00%) | 116 (0.00%) | 184 (0.00%) |
|  | C259 | 18 (0.00%) | 8 (0.00%) | 82 (0.00%) | 99 (0.00%) |
|  | C341 | 38 (0.01%) | 12 (0.00%) | 159 (0.01%) | 200 (0.00%) |
|  | C343 | 12 (0.00%) | 8 (0.00%) | 77 (0.00%) | 100 (0.00%) |
|  | C349 | 81 (0.01%) | 19 (0.01%) | 263 (0.01%) | 363 (0.01%) |
|  | C509 | 35 (0.01%) | 18 (0.01%) | 130 (0.00%) | 181 (0.00%) |
|  | C56 | 32 (0.00%) | 8 (0.00%) | 86 (0.00%) | 131 (0.00%) |
|  | C61 | 54 (0.01%) | 29 (0.01%) | 183 (0.01%) | 240 (0.01%) |
|  | C64 | 10 (0.00%) | 12 (0.00%) | 75 (0.00%) | 102 (0.00%) |
|  | C679 | 21 (0.00%) | 19 (0.01%) | 103 (0.00%) | 136 (0.00%) |
|  | C920 | 16 (0.00%) | 8 (0.00%) | 61 (0.00%) | 91 (0.00%) |
|  | N185 | 16 (0.00%) | 7 (0.00%) | 85 (0.00%) | 78 (0.00%) |
|  | Other | 674,969 (99.93%) | 266,388 (99.93%) | 2,699,912 (99.94%) | 4,030,667 (99.94%) |
| Type of vaccine at index date | High-dose QIV | 675,412 (100.00%) | 266,584 (100.00%) | 0 (0.00%) | 0 (0.00%) |
|  | Standard-dose QIV | 0 (0.00%) | 0 (0.00%) | 2,701,648 (100.00%) | 4,032,977 (100.00%) |
| Vaccine brand at index date | Efluelda | 675,412 (100.00%) | 266,584 (100.00%) | 0 (0.00%) | 0 (0.00%) |
|  | Fluarix Tetra | 0 (0.00%) | 0 (0.00%) | 137,886 (5.10%) | 211,102 (5.23%) |
|  | Influvac Tetra | 0 (0.00%) | 0 (0.00%) | 1,460,784 (54.07%) | 2,063,143 (51.16%) |
|  | Vaxigriptetra | 0 (0.00%) | 0 (0.00%) | 1,102,978 (40.83%) | 1,758,732 (43.61%) |
| Vaccine administration (Pharmacist vs. Other) | other | 348,674 (51.62%) | 141,612 (53.12%) | 1,401,557 (51.88%) | 1,790,174 (44.39%) |
|  | pharmacist | 326,738 (48.38%) | 124,972 (46.88%) | 1,300,091 (48.12%) | 2,242,803 (55.61%) |
| Week of vaccination | Week 1: 35 |  | 1 (0.00%) |  | 6 (0.00%) |
|  | Week 2: 36 |  | 0 (0.00%) |  | 6 (0.00%) |
|  | Week 3: 37 |  | 5 (0.00%) |  | 8 (0.00%) |
|  | Week 4: 38 |  | 31 (0.01%) |  | 74 (0.00%) |
|  | Week 5: 39 | 40 (0.01%) | 41 (0.02%) | 160 (0.01%) | 756 (0.02%) |
|  | Week 6: 40 | 451 (0.07%) | 259 (0.10%) | 1,804 (0.07%) | 1,759 (0.04%) |
|  | Week 7: 41 | 882 (0.13%) | 659 (0.25%) | 3,528 (0.13%) | 3,671 (0.09%) |
|  | Week 8: 42 | 190,496 (28.20%) | 144,706 (54.28%) | 761,984 (28.20%) | 487,073 (12.08%) |
|  | Week 9: 43 | 134,839 (19.96%) | 55,750 (20.91%) | 539,356 (19.96%) | 279,361 (6.93%) |
|  | Week 10: 44 | 102,925 (15.24%) | 24,190 (9.07%) | 411,700 (15.24%) | 295,508 (7.33%) |
|  | Week 11: 45 | 87,823 (13.00%) | 22,275 (8.36%) | 351,292 (13.00%) | 488,240 (12.11%) |
|  | Week 12: 46 | 63,029 (9.33%) | 15,138 (5.68%) | 252,116 (9.33%) | 620,343 (15.38%) |
|  | Week 13: 47 | 40,360 (5.98%) | 2,761 (1.04%) | 161,440 (5.98%) | 584,997 (14.51%) |
|  | Week 14: 48 | 26,150 (3.87%) | 68 (0.03%) | 104,600 (3.87%) | 576,805 (14.30%) |
|  | Week 15: 49 | 11,465 (1.70%) | 57 (0.02%) | 45,860 (1.70%) | 337,168 (8.36%) |
|  | Week 16: 50 | 5,726 (0.85%) | 32 (0.01%) | 22,904 (0.85%) | 163,779 (4.06%) |
|  | Week 17: 51 | 3,479 (0.52%) | 119 (0.04%) | 13,916 (0.52%) | 70,799 (1.76%) |
|  | Week 18: 52 | 2,784 (0.41%) | 54 (0.02%) | 11,136 (0.41%) | 47,811 (1.19%) |
|  | Week 19: 1 | 2,307 (0.34%) | 62 (0.02%) | 9,228 (0.34%) | 34,932 (0.87%) |
|  | Week 20: 2 | 1,167 (0.17%) | 92 (0.03%) | 4,668 (0.17%) | 17,535 (0.43%) |
|  | Week 21: 3 | 667 (0.10%) | 60 (0.02%) | 2,668 (0.10%) | 9,379 (0.23%) |
|  | Week 22: 4 | 448 (0.07%) | 74 (0.03%) | 1,792 (0.07%) | 6,857 (0.17%) |
|  | Week 23: 5 | 260 (0.04%) | 28 (0.01%) | 1,040 (0.04%) | 3,334 (0.08%) |
|  | Week 24: 6 | 57 (0.01%) | 11 (0.00%) | 228 (0.01%) | 997 (0.02%) |
|  | Week 25: 7 | 31 (0.00%) | 25 (0.01%) | 124 (0.00%) | 607 (0.02%) |
|  | Week 26: 8 | 15 (0.00%) | 23 (0.01%) | 60 (0.00%) | 447 (0.01%) |
|  | Week 27: 9 | 6 (0.00%) | 19 (0.01%) | 24 (0.00%) | 394 (0.01%) |
|  | Week 28: 10 | 2 (0.00%) | 5 (0.00%) | 8 (0.00%) | 101 (0.00%) |
|  | Week 29: 11 | 1 (0.00%) | 10 (0.00%) | 4 (0.00%) | 76 (0.00%) |
|  | Week 30: 12 |  | 7 (0.00%) |  | 90 (0.00%) |
|  | Week 31: 13 | 2 (0.00%) | 22 (0.01%) | 8 (0.00%) | 64 (0.00%) |
| Second brand of influenza vaccine received in the season | Missing | 675,412 | 266,584 | 2,701,648 | 4,032,977 |
|  | Vaxigriptetra |  |  |  |  |
| Number of seasons with influenza vaccination | N | 675,412 | 266,584 | 2,701,648 | 4,032,977 |
| in the 5 preceding seasons | Mean (±SD) | 3.27 (±1.14) | 3.57 (±0.86) | 3.28 (±1.14) | 2.99 (±1.29) |
|  | Min; Max | 0.00; 5.00 | 0.00; 4.00 | 0.00; 5.00 | 0.00; 5.00 |
|  | Median | 4.00 | 4.00 | 4.00 | 4.00 |
|  | Q1; Q3 | 3.00; 4.00 | 4.00; 4.00 | 3.00; 4.00 | 2.00; 4.00 |
| Number of seasons with influenza vaccination | 0 | 26,275 (3.89%) | 2,863 (1.07%) | 113,344 (4.20%) | 276,305 (6.85%) |
| in the 5 preceding seasons - categorical | 1 | 40,717 (6.03%) | 8,486 (3.18%) | 157,132 (5.82%) | 354,638 (8.79%) |
|  | 2 | 88,046 (13.04%) | 24,305 (9.12%) | 344,336 (12.75%) | 674,324 (16.72%) |
|  | 3 | 86,649 (12.83%) | 29,009 (10.88%) | 340,054 (12.59%) | 562,132 (13.94%) |
|  | 4 | 433,724 (64.22%) | 201,921 (75.74%) | 1,746,780 (64.66%) | 2,165,577 (53.70%) |
|  | 5 | 1 (0.00%) | 0 (0.00%) | 2 (0.00%) | 1 (0.00%) |
| Type of influenza vaccination | HD | 13,076 (1.94%) | 140,858 (52.84%) | 52,355 (1.94%) | 163,270 (4.05%) |
| in the previous season | Not vaccinated | 54,430 (8.06%) | 9,201 (3.45%) | 223,091 (8.26%) | 508,001 (12.60%) |
|  | SD | 607,873 (90.00%) | 116,238 (43.60%) | 2,426,121 (89.80%) | 3,361,244 (83.34%) |
|  | both | 33 (0.00%) | 287 (0.11%) | 81 (0.00%) | 462 (0.01%) |
| Number of influenza vaccination during | N | 675,412 | 266,584 | 2,701,648 | 4,032,977 |
| the 5 previous seasons (dispensation) | Mean (±SD) | 3.28 (±1.14) | 3.58 (±0.87) | 3.29 (±1.15) | 3.00 (±1.30) |
|  | Min; Max | 0.00; 11.00 | 0.00; 11.00 | 0.00; 13.00 | 0.00; 16.00 |
|  | Median | 4.00 | 4.00 | 4.00 | 4.00 |
|  | Q1; Q3 | 3.00; 4.00 | 4.00; 4.00 | 3.00; 4.00 | 2.00; 4.00 |
| Number of influenza vaccinations during | 0 | 26,277 (3.89%) | 2,863 (1.07%) | 113,346 (4.20%) | 276,315 (6.85%) |
| the 5 previous seasons (dispensation) - | 1 | 40,576 (6.01%) | 8,458 (3.17%) | 156,614 (5.80%) | 353,448 (8.76%) |
| categorical | 2 | 87,573 (12.97%) | 24,139 (9.05%) | 342,382 (12.67%) | 670,572 (16.63%) |
|  | 3 | 86,367 (12.79%) | 28,830 (10.81%) | 338,925 (12.55%) | 561,009 (13.91%) |
|  | 4 | 429,619 (63.61%) | 199,694 (74.91%) | 1,730,962 (64.07%) | 2,145,770 (53.21%) |
|  | 5 | 4,854 (0.72%) | 2,524 (0.95%) | 18,759 (0.69%) | 25,056 (0.62%) |
|  | 6 or more | 146 (0.02%) | 76 (0.03%) | 660 (0.02%) | 807 (0.02%) |
| Vaccination status for COVID-19 at index date | On-going | 902 (0.13%) | 362 (0.14%) | 3,544 (0.13%) | 5,182 (0.13%) |
|  | Primo-vaccination | 657,801 (97.39%) | 260,161 (97.59%) | 2,632,920 (97.46%) | 3,924,865 (97.32%) |
|  | Unvaccinated | 16,709 (2.47%) | 6,061 (2.27%) | 65,184 (2.41%) | 102,930 (2.55%) |
| Pneumococcal vaccination status (presence | No | 592,623 (87.74%) | 229,699 (86.16%) | 2,377,816 (88.01%) | 3,588,203 (88.97%) |
| during the 5 years preceeding index date) | Yes | 82,789 (12.26%) | 36,885 (13.84%) | 323,832 (11.99%) | 444,774 (11.03%) |
| UTI-related hospitalizations | No | 670,246 (99.24%) | 263,811 (98.96%) | 2,681,734 (99.26%) | 4,009,104 (99.41%) |
|  | Yes | 5,166 (0.76%) | 2,773 (1.04%) | 19,914 (0.74%) | 23,873 (0.59%) |
| Number of UTI-related hospitalizations | N | 675,412 | 266,584 | 2,701,648 | 4,032,977 |
|  | Mean (±SD) | 0.01 (±0.10) | 0.01 (±0.12) | 0.01 (±0.10) | 0.01 (±0.09) |
|  | Min; Max | 0.00; 15.00 | 0.00; 6.00 | 0.00; 16.00 | 0.00; 8.00 |
|  | Median | 0.00 | 0.00 | 0.00 | 0.00 |
|  | Q1; Q3 | 0.00; 0.00 | 0.00; 0.00 | 0.00; 0.00 | 0.00; 0.00 |
| Number of UTI-related hospitalizations | Missing | 670,246 | **263,811** | 2,681,734 | **4,009,104** |
| in patients with at least 1 UTI hospitalization | N | 5,166 | 2,773 | 19,914 | 23,873 |
|  | Mean (±SD) | 1.12 (±0.44) | 1.12 (±0.38) | 1.12 (±0.41) | 1.12 (±0.41) |
|  | Min; Max | 1.00; 15.00 | 1.00; 6.00 | 1.00; 16.00 | 1.00; 8.00 |
|  | Median | 1.00 | 1.00 | 1.00 | 1.00 |
|  | Q1; Q3 | 1.00; 1.00 | 1.00; 1.00 | 1.00; 1.00 | 1.00; 1.00 |
| Erysipelas-related hospitalizations | No | 674,343 (99.84%) | 265,955 (99.76%) | 2,697,435 (99.84%) | 4,027,925 (99.87%) |
|  | Yes | 1,069 (0.16%) | 629 (0.24%) | 4,213 (0.16%) | 5,052 (0.13%) |
| Number of erysipelas-related hospitalizations | N | 675,412 | 266,584 | 2,701,648 | 4,032,977 |
|  | Mean (±SD) | 0.00 (±0.05) | 0.00 (±0.06) | 0.00 (±0.05) | 0.00 (±0.04) |
|  | Min; Max | 0.00; 4.00 | 0.00; 5.00 | 0.00; 8.00 | 0.00; 9.00 |
|  | Median | 0.00 | 0.00 | 0.00 | 0.00 |
|  | Q1; Q3 | 0.00; 0.00 | 0.00; 0.00 | 0.00; 0.00 | 0.00; 0.00 |
| Number of erysipelas-related hospitalizations | Missing | 674,343 | **265,955** | 2,697,435 | **4,027,925** |
| in patients with at least 1 erysipelas | N | 1,069 | 629 | 4,213 | 5,052 |
| hospitalization | Mean (±SD) | 1.13 (±0.38) | 1.14 (±0.45) | 1.15 (±0.44) | 1.14 (±0.42) |
|  | Min; Max | 1.00; 4.00 | 1.00; 5.00 | 1.00; 8.00 | 1.00; 9.00 |
|  | Median | 1.00 | 1.00 | 1.00 | 1.00 |
|  | Q1; Q3 | 1.00; 1.00 | 1.00; 1.00 | 1.00; 1.00 | 1.00; 1.00 |
| Cataract-related hospitalizations | No | 653,032 (96.69%) | 257,587 (96.63%) | 2,613,255 (96.73%) | 3,911,120 (96.98%) |
|  | Yes | 22,380 (3.31%) | 8,997 (3.37%) | 88,393 (3.27%) | 121,857 (3.02%) |
| Number of cataract-related hospitalizations | N | 675,412 | 266,584 | 2,701,648 | 4,032,977 |
|  | Mean (±SD) | 0.05 (±0.29) | 0.05 (±0.29) | 0.05 (±0.28) | 0.05 (±0.27) |
|  | Min; Max | 0.00; 3.00 | 0.00; 3.00 | 0.00; 3.00 | 0.00; 5.00 |
|  | Median | 0.00 | 0.00 | 0.00 | 0.00 |
|  | Q1; Q3 | 0.00; 0.00 | 0.00; 0.00 | 0.00; 0.00 | 0.00; 0.00 |
| Number of cataract-related hospitalizations | Missing | 653,032 | **257,587** | 2,613,255 | **3,911,120** |
| in patients with at least 1 cataract | N | 22,38 | 8,997 | 88,393 | 121,857 |
| hospitalization | Mean (±SD) | 1.52 (±0.50) | 1.51 (±0.50) | 1.52 (±0.50) | 1.51 (±0.50) |
|  | Min; Max | 1.00; 3.00 | 1.00; 3.00 | 1.00; 3.00 | 1.00; 5.00 |
|  | Median | 2.00 | 2.00 | 2.00 | 2.00 |
|  | Q1; Q3 | 1.00; 2.00 | 1.00; 2.00 | 1.00; 2.00 | 1.00; 2.00 |
| Presence of influenza hospitalizations | No | 674,593 (99.88%) | 266,073 (99.81%) | 2,697,564 (99.85%) | 4,028,176 (99.88%) |
|  | Yes | 819 (0.12%) | 511 (0.19%) | 4,084 (0.15%) | 4,801 (0.12%) |
| Number of influenza hospitalizations | N | 675,412 | 266,584 | 2,701,648 | 4,032,977 |
|  | Mean (±SD) | 0.00 (±0.04) | 0.00 (±0.05) | 0.00 (±0.04) | 0.00 (±0.04) |
|  | Min; Max | 0.00; 3.00 | 0.00; 4.00 | 0.00; 3.00 | 0.00; 3.00 |
|  | Median | 0.00 | 0.00 | 0.00 | 0.00 |
|  | Q1; Q3 | 0.00; 0.00 | 0.00; 0.00 | 0.00; 0.00 | 0.00; 0.00 |
| Number of influenza hospitalizations | Missing | 674,593 | 266,073 | 2,697,564 | 4,028,176 |
| in patients with at least one influenza | N | 819 | 511 | 4,084 | 4,801 |
| hospitalization | Mean (±SD) | 1.10 (±0.32) | 1.13 (±0.37) | 1.08 (±0.30) | 1.08 (±0.29) |
|  | Min; Max | 1.00; 3.00 | 1.00; 4.00 | 1.00; 3.00 | 1.00; 3.00 |
|  | Median | 1.00 | 1.00 | 1.00 | 1.00 |
|  | Q1; Q3 | 1.00; 1.00 | 1.00; 1.00 | 1.00; 1.00 | 1.00; 1.00 |
| Presence of pneumonia hospitalizations | No | 667,116 (98.77%) | 261,756 (98.19%) | 2,669,426 (98.81%) | 3,995,150 (99.06%) |
|  | Yes | 8,296 (1.23%) | 4,828 (1.81%) | 32,222 (1.19%) | 37,827 (0.94%) |
| Number of pneumonia hospitalizations | N | 675,412 | 266,584 | 2,701,648 | 4,032,977 |
|  | Mean (±SD) | 0.01 (±0.13) | 0.02 (±0.17) | 0.01 (±0.13) | 0.01 (±0.12) |
|  | Min; Max | 0.00; 6.00 | 0.00; 6.00 | 0.00; 9.00 | 0.00; 7.00 |
|  | Median | 0.00 | 0.00 | 0.00 | 0.00 |
|  | Q1; Q3 | 0.00; 0.00 | 0.00; 0.00 | 0.00; 0.00 | 0.00; 0.00 |
| Number of pneumonia hospitalizations | Missing | 667,116 | 261,756 | 2,669,426 | 3,995,150 |
| in patients with at least 1 pneumonia | N | 8,296 | 4,828 | 32,222 | 37,827 |
| hospitalization | Mean (±SD) | 1.14 (±0.40) | 1.16 (±0.44) | 1.14 (±0.41) | 1.14 (±0.40) |
|  | Min; Max | 1.00; 6.00 | 1.00; 6.00 | 1.00; 9.00 | 1.00; 7.00 |
|  | Median | 1.00 | 1.00 | 1.00 | 1.00 |
|  | Q1; Q3 | 1.00; 1.00 | 1.00; 1.00 | 1.00; 1.00 | 1.00; 1.00 |
| Presence of P/I hospitalizations | No | 666,503 (98.68%) | 261,382 (98.05%) | 2,666,280 (98.69%) | 3,991,491 (98.97%) |
|  | Yes | 8,909 (1.32%) | 5,202 (1.95%) | 35,368 (1.31%) | 41,486 (1.03%) |
| Number of P/I hospitalizations | N | 675,412 | 266,584 | 2,701,648 | 4,032,977 |
|  | Mean (±SD) | 0.02 (±0.14) | 0.02 (±0.18) | 0.02 (±0.14) | 0.01 (±0.13) |
|  | Min; Max | 0.00; 6.00 | 0.00; 6.00 | 0.00; 9.00 | 0.00; 7.00 |
|  | Median | 0.00 | 0.00 | 0.00 | 0.00 |
|  | Q1; Q3 | 0.00; 0.00 | 0.00; 0.00 | 0.00; 0.00 | 0.00; 0.00 |
| Number of P/I hospitalizations in patients | Missing | 666,503 | 261,382 | 2,666,280 | 3,991,491 |
| with at least 1 influenza or 1 pneumonia | N | 8,909 | 5,202 | 35,368 | 41,486 |
| hospitalization | Mean (±SD) | 1.16 (±0.44) | 1.19 (±0.48) | 1.16 (±0.44) | 1.16 (±0.44) |
|  | Min; Max | 1.00; 6.00 | 1.00; 6.00 | 1.00; 9.00 | 1.00; 7.00 |
|  | Median | 1.00 | 1.00 | 1.00 | 1.00 |
|  | Q1; Q3 | 1.00; 1.00 | 1.00; 1.00 | 1.00; 1.00 | 1.00; 1.00 |
| Presence of respiratory disease hospitalizations | No | 662,405 (98.07%) | 259,207 (97.23%) | 2,650,886 (98.12%) | 3,972,900 (98.51%) |
|  | Yes | 13,007 (1.93%) | 7,377 (2.77%) | 50,762 (1.88%) | 60,077 (1.49%) |
| Number of respiratory disease hospitalizations | N | 675,412 | 266,584 | 2,701,648 | 4,032,977 |
|  | Mean (±SD) | 0.02 (±0.18) | 0.03 (±0.22) | 0.02 (±0.18) | 0.02 (±0.16) |
|  | Min; Max | 0.00; 8.00 | 0.00; 7.00 | 0.00; 9.00 | 0.00; 9.00 |
|  | Median | 0.00 | 0.00 | 0.00 | 0.00 |
|  | Q1; Q3 | 0.00; 0.00 | 0.00; 0.00 | 0.00; 0.00 | 0.00; 0.00 |
| Number of respiratory hospitalizations | Missing | 662,405 | 259,207 | 2,650,886 | 3,972,900 |
| in patients with at least 1 respiratory | N | 13,007 | 7,377 | 50,762 | 60,077 |
| hospitalization | Mean (±SD) | 1.20 (±0.50) | 1.22 (±0.55) | 1.19 (±0.51) | 1.19 (±0.50) |
|  | Min; Max | 1.00; 8.00 | 1.00; 7.00 | 1.00; 9.00 | 1.00; 9.00 |
|  | Median | 1.00 | 1.00 | 1.00 | 1.00 |
|  | Q1; Q3 | 1.00; 1.00 | 1.00; 1.00 | 1.00; 1.00 | 1.00; 1.00 |
| Presence of cardiovascular disease | No | 643,744 (95.31%) | 249,745 (93.68%) | 2,579,315 (95.47%) | 3,883,943 (96.30%) |
| hospitalizations | Yes | 31,668 (4.69%) | 16,839 (6.32%) | 122,333 (4.53%) | 149,034 (3.70%) |
| Number of cardiovascular disease | N | 675,412 | 266,584 | 2,701,648 | 4,032,977 |
| hospitalizations | Mean (±SD) | 0.06 (±0.32) | 0.09 (±0.38) | 0.06 (±0.31) | 0.05 (±0.28) |
|  | Min; Max | 0.00; 12.00 | 0.00; 11.00 | 0.00; 14.00 | 0.00; 12.00 |
|  | Median | 0.00 | 0.00 | 0.00 | 0.00 |
|  | Q1; Q3 | 0.00; 0.00 | 0.00; 0.00 | 0.00; 0.00 | 0.00; 0.00 |
| Number of cardiovascular | Missing | 643,744 | 249,745 | 2,579,315 | 3,883,943 |
| hospitalizations in patients with at least 1 | N | 31,668 | 16,839 | 122,333 | 149,034 |
| cardiovascular hospitalization | Mean (±SD) | 1.32 (±0.69) | 1.35 (±0.73) | 1.32 (±0.69) | 1.30 (±0.67) |
|  | Min; Max | 1.00; 12.00 | 1.00; 11.00 | 1.00; 14.00 | 1.00; 12.00 |
|  | Median | 1.00 | 1.00 | 1.00 | 1.00 |
|  | Q1; Q3 | 1.00; 1.00 | 1.00; 1.00 | 1.00; 1.00 | 1.00; 1.00 |
| Presence of cardiorespiratory disease | No | 640,640 (94.85%) | 248,083 (93.06%) | 2,566,651 (95.00%) | 3,868,436 (95.92%) |
| hospitalizations | Yes | 34,772 (5.15%) | 18,501 (6.94%) | 134,997 (5.00%) | 164,541 (4.08%) |
| Number of cardiorespiratory disease | N | 675,412 | 266,584 | 2,701,648 | 4,032,977 |
| hospitalizations | Mean (±SD) | 0.08 (±0.43) | 0.12 (±0.53) | 0.08 (±0.43) | 0.07 (±0.38) |
|  | Min; Max | 0.00; 15.00 | 0.00; 18.00 | 0.00; 18.00 | 0.00; 18.00 |
|  | Median | 0.00 | 0.00 | 0.00 | 0.00 |
|  | Q1; Q3 | 0.00; 0.00 | 0.00; 0.00 | 0.00; 0.00 | 0.00; 0.00 |
| Number of cardiorespiratory hospitalizations | Missing | 640,64 | 248,083 | 2,566,651 | 3,868,436 |
| in patients with at least one cardiorespiratory | N | 34,772 | 18,501 | 134,997 | 164,541 |
| hospitalization | Mean (±SD) | 1.65 (±1.04) | 1.71 (±1.12) | 1.64 (±1.04) | 1.62 (±1.02) |
|  | Min; Max | 1.00; 15.00 | 1.00; 18.00 | 1.00; 18.00 | 1.00; 18.00 |
|  | Median | 1.00 | 1.00 | 1.00 | 1.00 |
|  | Q1; Q3 | 1.00; 2.00 | 1.00; 2.00 | 1.00; 2.00 | 1.00; 2.00 |
| Long Term Disease ICD-10 and name | Other | 159,181 (32.35%) | 68,647 (30.90%) | 608,520 (31.91%) | 856,160 (32.25%) |
|  | C18 |  | 3,367 (1.52%) |  | 38,095 (1.43%) |
|  | C50 | 9,265 (1.88%) | 8,320 (3.74%) | 36,575 (1.92%) | 116,271 (4.38%) |
|  | C61 | 6,709 (1.36%) | 9,686 (4.36%) | 26,642 (1.40%) | 123,774 (4.66%) |
|  | C67 |  | 2,623 (1.18%) |  | 31,301 (1.18%) |
|  | E10 |  | 2,775 (1.25%) |  | 33,884 (1.28%) |
|  | E11 | 104,173 (21.17%) | 45,689 (20.56%) | 409,195 (21.46%) | 566,541 (21.34%) |
|  | F00 (G30.-) |  | 5,237 (2.36%) |  | 35,197 (1.33%) |
|  | F01 | 12,142 (2.47%) |  | 47,301 (2.48%) |  |
|  | F03 | 43,750 (8.89%) |  | 170,642 (8.95%) |  |
|  | F29 | 5,055 (1.03%) |  | 19,271 (1.01%) |  |
|  | F31 | 5,594 (1.14%) |  | 21,926 (1.15%) |  |
|  | F32 | 8,012 (1.63%) |  | 32,134 (1.68%) |  |
|  | G20 | 9,827 (2.00%) | 2,978 (1.34%) | 37,326 (1.96%) | 35,555 (1.34%) |
|  | I10 | 6,962 (1.41%) | 3,332 (1.50%) | 25,299 (1.33%) | 24,812 (0.93%) |
|  | I21 |  | 3,741 (1.68%) |  | 54,383 (2.05%) |
|  | I25 | 21,568 (4.38%) | 19,864 (8.94%) | 84,526 (4.43%) | 245,344 (9.24%) |
|  | I35 |  | 2,758 (1.24%) |  | 29,888 (1.13%) |
|  | I48 | 35,422 (7.20%) | 17,163 (7.72%) | 138,950 (7.29%) | 182,429 (6.87%) |
|  | I49 | 6,958 (1.41%) | 3,337 (1.50%) | 27,162 (1.42%) | 34,683 (1.31%) |
|  | I50 | 9,610 (1.95%) | 5,029 (2.26%) | 37,811 (1.98%) | 48,561 (1.83%) |
|  | I64 | 19,826 (4.03%) | 5,709 (2.57%) | 76,717 (4.02%) | 63,503 (2.39%) |
|  | I69 | 7,771 (1.58%) |  | 29,378 (1.54%) |  |
|  | I70 | 7,636 (1.55%) | 4,065 (1.83%) | 29,128 (1.53%) | 53,696 (2.02%) |
|  | I702 | 5,709 (1.16%) | 3,532 (1.59%) | 22,345 (1.17%) | 40,253 (1.52%) |
|  | N18 | 6,905 (1.40%) | 4,338 (1.95%) | 26,255 (1.38%) | 40,601 (1.53%) |
