## Supplementary figures and images for "Relative effectiveness of high-dose vs standard-dose influenza vaccines in preventing hospitalizations: a national retrospective cohort study in France, 2022/23 season"

### Supplemental Figure 1

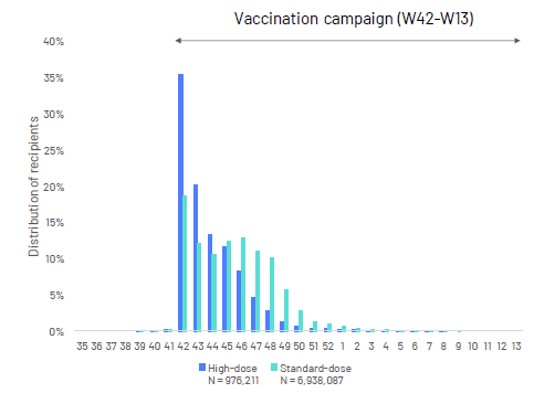
